# An ecological comparison of hospital-level antibiotic use and mortality in 36,124,372 acute/general medicine inpatients in England

**DOI:** 10.1101/2020.09.24.20199448

**Authors:** Eric P Budgell, Tim Davies, Tjibbe Donker, Susan Hopkins, David Wyllie, Tim E A Peto, Martin Gill, Martin Llewelyn, A. Sarah Walker

## Abstract

**Objectives:** To determine the extent to which variation in hospital antibiotic prescribing is associated with mortality risk in acute/general medicine inpatients.

**Design:** Ecological analysis, using electronic health records from Hospital Episode Statistics (HES) and antibiotic data from IQVIA.

**Setting:** 135 acute National Health Service (NHS) hospital Trusts in England.

**Participants:** 36,124,372 acute/general medicine inpatients (≥16 years old at admission) admitted between 01/April/2010-31/March/2017 (median age 66 years, 50.4% female, 83.8% white ethnicity).

**Main outcome measures:** Random-effects meta-regression was used to investigate whether heterogeneity in the adjusted probability of death within 30-days of admission was associated with hospital-level antibiotic use, measured in defined-daily-doses (DDD)/1,000 bed-days. Models also considered DDDs/1,000 admissions and DDDs for selected antibiotics, including narrow-spectrum/broad-spectrum, inpatient/outpatient, parenteral/oral, piperacillin-tazobactam and meropenem, and Public Health England interpretations of World Health Organization Access, Watch, and Reserve antibiotics. Secondary analyses examined 14-day mortality and non-elective re-admission to hospital within 30-days of discharge.

**Results:** There was a 15-fold variation in hospital-level DDDs/1,000 bed-days and comparable or greater variation in broad-spectrum, parenteral, and Reserve antibiotic use. After adjusting for a wide range of admission factors to reflect varying case-mix across hospitals, the adjusted probability of 30-day mortality changed by −0.010% (95% CI: −0.064 to +0.044) for each increase in hospital-level antibiotic use of 500 DDDs/1,000 bed-days. Analyses focusing on other metrics of antibiotic use, sub-populations, and 14-day mortality also showed no consistent association with the adjusted probability of death.

**Discussion:** We find no evidence that the wide variation in antibiotic use across NHS hospitals is associated with case-mix adjusted mortality risk in acute/general medicine inpatients. Our results indicate that hospital antibiotic use in the acute/general medicine population could be safely cut by up to one-third, greatly exceeding the 1% year-on-year reductions required of NHS hospitals.

**What is already known on this topic:**

- Previous studies have reported wide variation in both recommended antibiotic prescribing duration and total antibiotic consumption among acute hospitals.
- In hospitals with more acute patients, systematic under-treatment might reasonably be expected to harm patients, and though a growing body of evidence shows reducing hospital antibiotic overuse may be done safely, there is a lack of good data to indicate how much it may be possible to safely reduce use
- Examination of the possibility that substantially driving down antibiotic use could compromise clinical outcomes is needed to reassure practitioners and the public that substantially reducing antibiotic use is safe.

**What this study adds:**

- After adjusting for a wide range of admission factors to reflect varying case-mix across acute hospitals, we observed no consistent association between 24 metrics of hospital-level antibiotic use and the adjusted probability of death in a large national cohort of over 36 million acute/general medicine inpatients
- These findings indicate that at many hospitals patients are receiving considerably more antibiotics than necessary to treat their acute infections, and we estimate system-wide reductions of up to one-third of antibiotic defined-daily-doses (DDDs) could be achieved safely among medical admissions.
- The magnitude of the antibiotic reductions that could be safely achieved dwarf the 1% year-on-year reductions required of NHS hospitals.

## INTRODUCTION

Antibiotic overuse puts individual patients(1) and whole populations(2) at risk of antimicrobial resistance (AMR). AMR infections cause higher mortality, longer hospital admissions, and increased costs of care.(3) Reducing unnecessary antibiotic use is therefore essential to reduce the selective pressure for resistance(4,5) and avoid other harms including adverse drug events,(6) toxicity,(7–9) changes to the gut microbiome,(10,11) and increased susceptibility to infections such as *Clostridioides difficile*.(12–15) The urgency of this problem is reflected in many national plans to tackle AMR, with 26/30 countries in Europe, Canada, and the United States having established or planned targets to reduce antimicrobial use in humans,(16) including the UK which aims to reduce antimicrobial use by 15% between 2019-2024.(17)

In England’s National Health Service (NHS) primary care antibiotic stewardship initiatives focusing on decisions to start antibiotics have been successful in reducing antibiotic overuse(18), but in hospital practice, prescribers must balance the risks of harm posed by under-treating an infection and those of prolonged or excessively-broad spectrum antibiotic use.(19) For patients with life-threatening infections, even modest treatment delays can increase mortality risk,(20) and diagnostic uncertainty means it is often necessary to administer broad-spectrum antibiotics empirically while waiting for additional clinical, microbiological, or radiographic information.

For these reasons, controlling antibiotic overuse in hospitals depends on early antibiotic initiation followed by the review of prescriptions after 24-72 hours to re-evaluate whether continued therapy is appropriate.(21) Different healthcare systems operationalise this approach in different ways, including through “antibiotic timeouts” in the United States(22) and “Start Smart then Focus” in England.(21) Yet opportunities to stop early or “de-escalate” – i.e. switch from parenteral to oral antibiotics, or to agents with a narrower spectrum of activity – are often missed. Estimates suggest 20-30% of prescriptions may be safely stopped at review,(23) but stop and review dates are poorly documented(24) and fewer than 10% of antibiotic prescriptions are stopped early.(25,26) Similarly, audits in England have shown one in six patients who are eligible for de-escalation at 72 hours are not switched,(27) while data from US hospitals show only 9% of inpatients on empiric antibiotics have narrowed or discontinued therapy within three days of starting treatment.(28) In NHS hospitals antibiotic consumption has continued to increase year-on-year(18) despite the introduction of financial incentives to reduce overuse.(29)

The issues of clinical urgency and diagnostic uncertainty which make limiting antibiotic overuse in hospitals challenging for clinicians also make attempts to define inappropriate use inherently subjective.(30) Another way to approach this problem is through “benchmarking”, whereby low-prescribing organisations are used to drive improvements.(31–33) Previous studies have reported wide variation in both recommended antibiotic prescribing duration(34) and total antibiotic consumption(35,36) among acute hospitals. However, simple comparisons of hospital-level consumption data largely fail to account for case-mix, and in hospitals with more acute patients, systematic under-treatment might reasonably be expected to harm patients. Antibiotic stewardship leads at acute hospitals in England have also expressed widespread concern about the safety of a target-driven antibiotic reduction strategy.(26) As a result, many hospitals are benchmarked against their own historical performance rather than externally, and only relatively small reduction targets are sought, such as the 1% year-on-year reduction target used in the NHS Standard Contract with hospital Trusts.(37) Examination of the possibility that substantially driving down antibiotic use could compromise clinical outcomes is needed to reassure practitioners and the public that substantially reducing antibiotic use is safe.

This study therefore aimed to determine the extent to which variation in antibiotic prescribing was associated with mortality risk in acute/general medicine inpatients in England.

## METHODS

### Data sources

Patient data was obtained through NHS Digital’s Data Access Request Service, which provided an extract from the Health Episode Statistics (HES) Admitted Patient Care data warehouse. This database captures all emergency, planned, and day-case admissions requiring an NHS hospital bed in England, but excludes outpatient visits and A&E attendances.(38) General medical patients may be treated under several different adult specialties, with coding practices varying by hospital Trust. The study population was therefore defined using multiple consultant specialty codes (Figure 1). This definition was selected to align with the most commonly used HES specialty codes used to admit adult general medicine inpatients. All inpatient spells and episodes were requested for patients with any eligible admission, in order to calculate hospital exposure.

**Figure 1:**
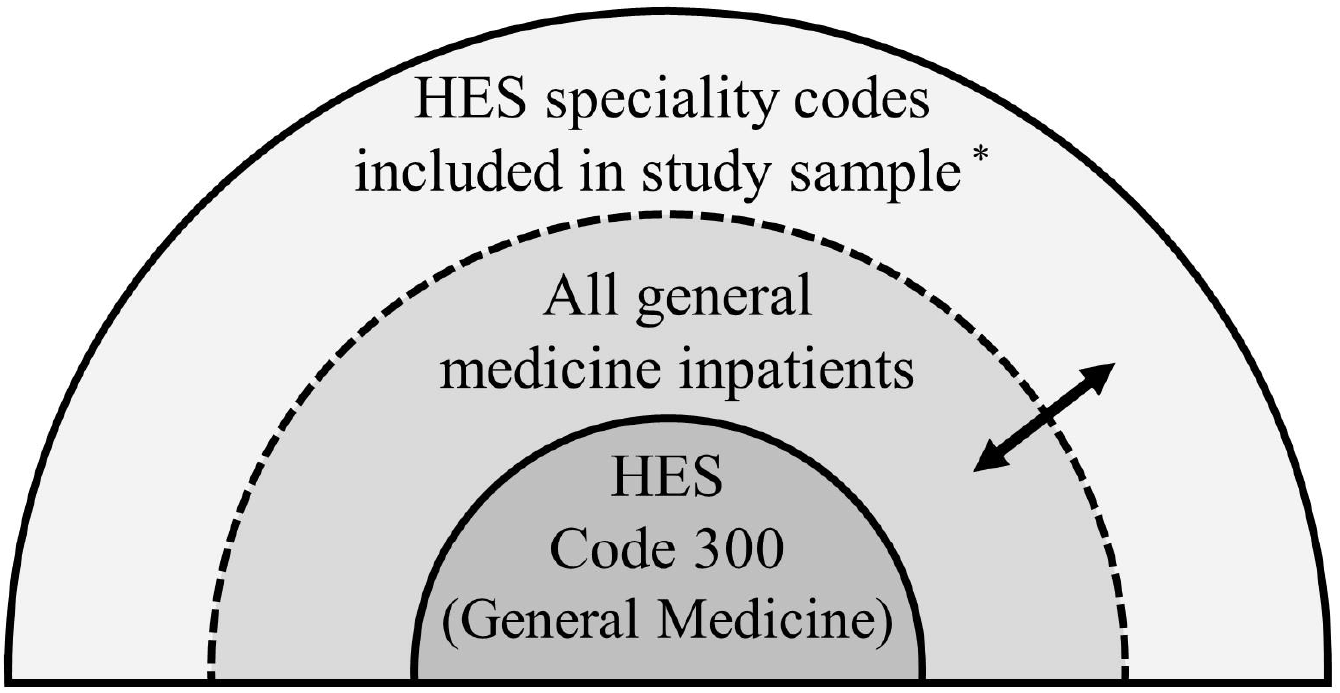
Study sample. ^*^Admissions were included if the specialty under which attending consultants were contracted (HES field: mainspef) or worked (HES field: tretspef) included any of the following HES codes in the first or second episode: acute/general medicine (300), gastroenterology (301), endocrinology (302), haematology (303), diabetic medicine (307), cardiology (320), acute internal medicine (326), respiratory/thoracic medicine (340), infectious diseases (350), neurology (400), rheumatology (410), and geriatric medicine (430). Given the variation in coding practices among hospitals, admission codes in general medicine (300) are likely only a subset of general medicine inpatients (perfect specificity, but less than perfect sensitivity). By contrast, the wider set of admission codes used to define this study sample is likely to have very high sensitivity but at the cost of reduced specificity. The extent of the reduction in specificity will vary by hospital depending on local coding practices.

The data contained a pseudonymised patient ID that is unique across hospital Trusts and over time,(39) patient information (sex, age at admission, ethnicity), clinical information (diagnoses), geographic information (admitting hospital, Index of Multiple Deprivation (IMD)), and administrative information such as admission and discharge dates, method and source of admission, method and destination of discharge, patient classification (day-case or ordinary admission), the specialties under which attending consultants were contracted or worked, and the start and end dates of consultant care episodes. A binary measure of death within 14 and 30 days of admission (all cause, in/out of hospital) was obtained through linkage with data from the Office of National Statistics (ONS), performed in advance by NHS Digital.

Information on NHS hospital-level antibiotic consumption was obtained from pharmacy dispensing records provided by IQVIA (formerly Quintiles and IMS Health, Inc)(40) through Public Health England (PHE) and measured in defined-daily-doses (DDDs), which enable standardised comparisons of antibiotic use and are defined by the World Health Organisation (WHO) as the average maintenance dose per day for a drug used for its main indication in adults.(41) The DDDs were calculated using the WHO Anatomical Therapeutic Chemical (ATC) DDD Index 2020 and disaggregated by drug, route of administration, and quarter/year (April 2014 to March 2017) for the included 135 NHS Trusts. Data was only available at the hospital Trust-level (not by specialty) and only from April 2014. Bed-days by hospital Trust were obtained from PHE(42) and admissions by hospital Trust were obtained from NHS Digital.(43)

### Patient and Public Involvement (PPI)

Our lead PPI representative had input into the conception and design of this study as part of the original programme grant. This study is based on routinely collected de-identified electronic medical records in HES and therefore members of the public were not involved in the data analysis; they were also not involved in the interpretation or reporting of the results. There are no plans to disseminate the results to the study participants as the data were de-identified.

### Data Cleaning

The HES data extract included 88,718,419 hospital admissions (“spells”) from 15,708,476 patients admitted between 1 April 2009 and 31 March 2017. Data cleaning steps are outlined in Appendix Figure A1. The merging and splitting of NHS hospital Trusts over time was accounted for by updating provider codes so they were current at March 2017 (Appendix Table A1).(44) This left admissions from 188 NHS hospital Trusts, defined as spells with a provider code of treatment beginning with “R” (thereby excluding primary care Trusts, independent providers and NHS treatment centres).(45) To improve model stability, 49 hospital Trusts with fewer than 50,000 admissions between April/2010 and March/2017 were excluded, as were four specialist hospital Trusts which lacked admissions in general medicine, producing an analytic cohort with 135 acute hospital Trusts in England for which antibiotic data was merged (subsequently denoted “hospitals”).

All data processing was carried out in Stata/MP 16 with data held on NHS servers located at the Oxford University Hospitals NHS Trust. We followed the RECORD statement(46)—an extension of the STROBE reporting guideline(47) for routinely collected observational health data (Appendix Table A2).

### Primary statistical analysis

The primary ecological analysis employed a meta-regression(48) on hospital-level summary data to compare outcomes in acute/general medicine inpatients with hospital-level antibiotic use. This involved deriving confounding-adjusted relative risks of death (all-cause, in/out of hospital) within 30 days of admission using Poisson regression with a robust variance adjustment by patient.(49) Time-to-event models could not be used as they required knowledge of date of death, which is available from ONS but considered identifiable by NHS Digital; only a binary indicator of death was therefore available for analysis. Models included admissions between April 2010 to March 2017, with prior data used only to calculate previous hospital exposure. A separate model was fit to each hospital (135 models) to allow each potential confounder to have a different impact on the outcome in each hospital, including the same factors in each model regardless of statistical significance. These multivariate models included an *a priori* list of 16 admission factors used to adjust for case-mix in previous analyses (Table 1) and nine interaction terms that improved model fit by lowering the Bayesian information criterion (BIC) of a single multivariate model applied to all hospitals (with hospital as a main effect).(50) Continuous factors were truncated at 2.5^th^ and 95^th^ or 99^th^ percentiles to reduce the influence of outliers, and natural cubic splines were used to account for non-linear effects of continuous covariates if they lowered the BIC of a model containing hospital as a factor; the number of knots was chosen based on BIC (Appendix Figure A2). Marginal effects (conditional adjusted predictions) were then derived, along with their standard errors, at the reference level of all model covariates in each hospital.(51)

**Table 1:**
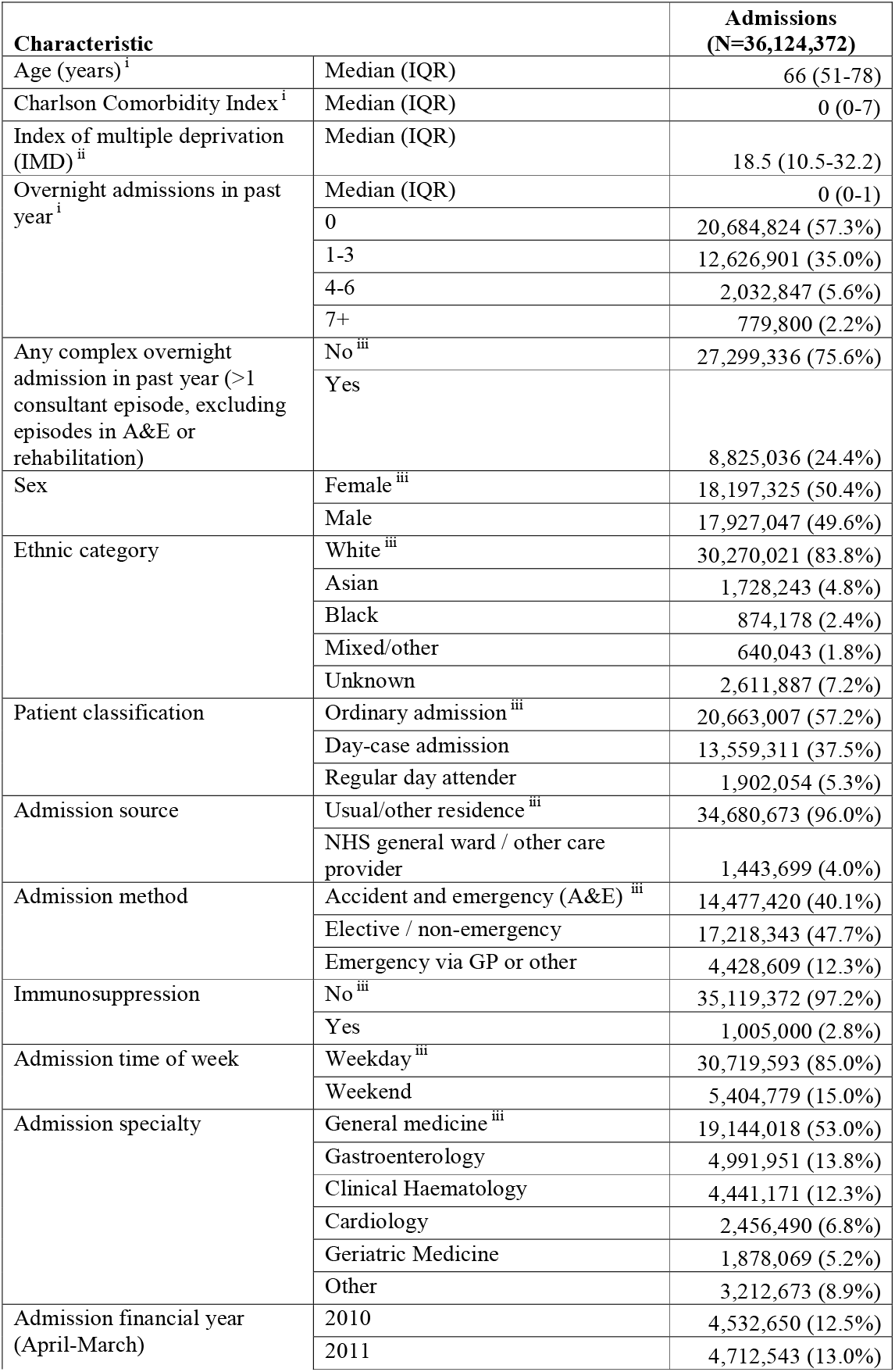
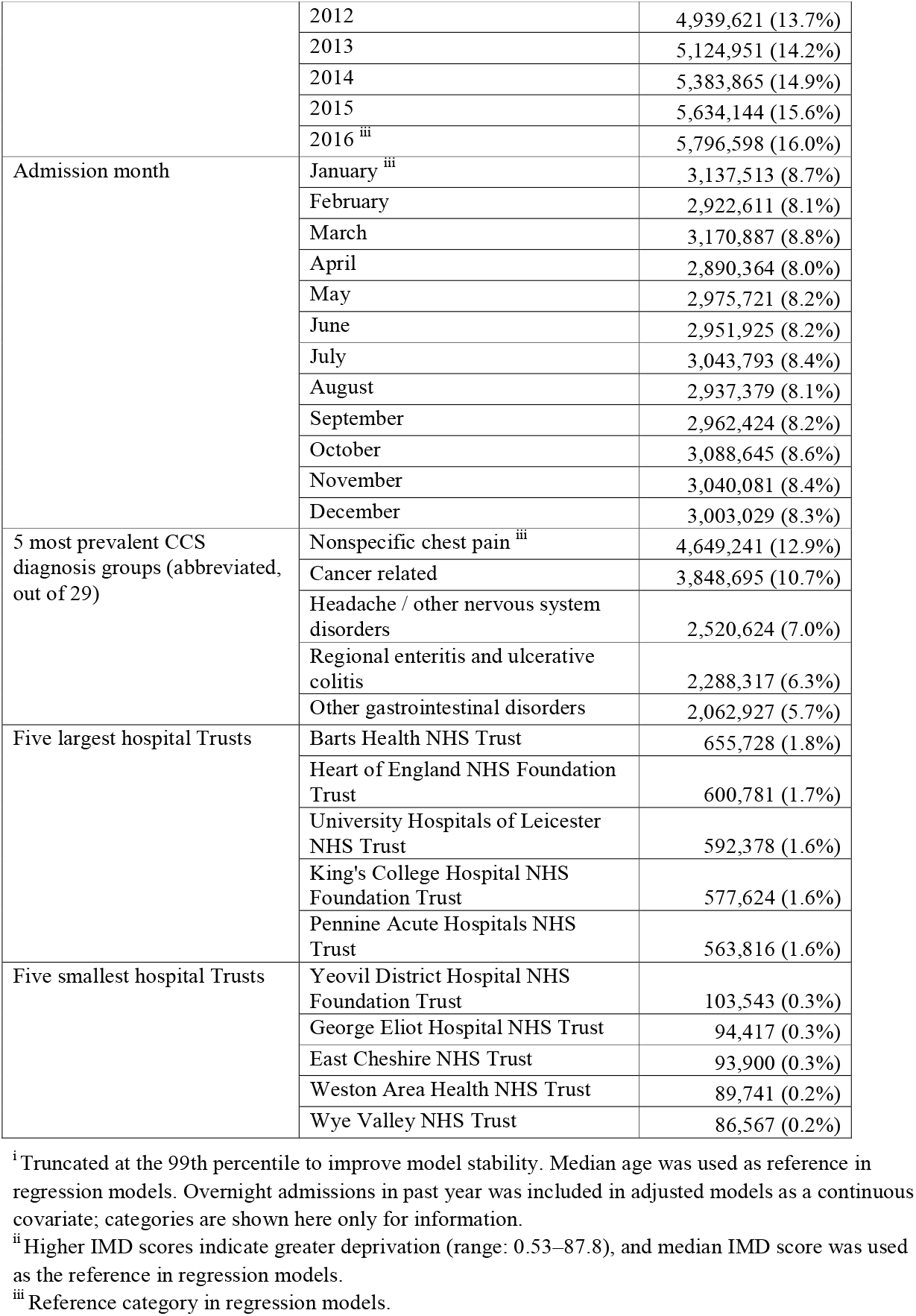
Admission Characteristics of 36,124,372 Acute/General Medical Inpatient Admissions in 135 Acute Care NHS Hospital Trusts, England, April 2010 to March 2017.

Finally, random effects meta-regression was used to investigate whether heterogeneity in the adjusted probability of death (one estimate per hospital) was associated with hospital-level antibiotic use, measured in total DDDs/1,000 bed-days. Hospital-level antibiotic data was used as a proxy for consumption in acute/general medicine as specialty-level data was not available during the study period. The DDD estimates were calculated as the annual mean of antibiotic data from April 2014 to March 2017. Multiple other metrics of antibiotic use were also considered, including inpatient and outpatient DDDs, narrow-spectrum and broad-spectrum DDDs, parenteral and oral DDDs, DDDs for piperacillin/tazobactam and meropenem specifically, and DDDs for PHE-interpretations of WHO Access, Watch, and Reserve (AWaRe) antibiotics,(52) which were introduced in WHO’s 2017 Essential Medicines List to improve access and monitoring of important or ‘last resort’ antibiotics (Appendix Table A3). The minor PHE amendments to AWaRe classifications reflected which antibiotics NHS hospitals should be prioritizing for human use. Meta-regression models adjusted for hospital size measured in terciles of either bed-days(42) or admissions,(43) depending on whether DDDs was measured per bed-days or per admissions. An interaction between DDDs and hospital size was assessed and retained where the heterogeneity p<0.01. Meta-regressions were fit on both the log probability and the probability scale; the latter are presented here to aid interpretation of observed associations.

In sensitivity analyses, a more narrow definition of general medicine was used, where all spells had a HES main specialty code or treatment specialty code of 300 in the first or second consultant episode. As outlined in Figure 1, this definition is likely to have perfect specificity but varying sensitivity among hospitals due to local variations in coding practices. Since antibiotic data was only available from April 2014, a separate sensitivity test restricted the study sample to admissions between 01/April/2014 and 31/March/2017.

### Secondary outcomes

Secondary outcomes included death within 14 days of admission (all cause, in/out of hospital) and non-elective re-admission to hospital within 30 days of discharge (regardless of re-admission speciality). Analyses of re-admission were restricted to patients discharged alive more than 30 days before 31 March 2017. Without death date, deaths outside hospital could not be treated as competing events for re-admission. Poisson models for each binary outcome adjusted for the same admission characteristics and interaction terms and this was followed by random effects meta-regression as previously described.

## RESULTS

### Primary statistical analysis

The final analytic cohort contained 36,124,372 acute/general medicine admissions from 12,320,069 patients between April 2010 and March 2017 inclusive, with median of two admissions (IQR: 1-3) per patient. Admissions increased year-on-year admissions (Table 1). The largest and smallest hospitals were Barts Health NHS Trust (655,728 admissions) and Wye Valley NHS Trust (86,567 admissions) respectively (complete list in Appendix Table A4). Patients had a median age of 66 years (IQR: 51-78), the most prevalent admission characteristics were female sex (50.4%), ethnically white (83.8%), admission on a weekday (85.0%), a low median Charlson Comorbidity Index (0, IQR: 0-7), low median IMD score (18.5, IQR: 10.5-32.2), and a Clinical Classifications Software (CCS) diagnosis group indicating non-specific chest pain (12.9%) (see Appendix Table A5 for other CCS groups).

In primary analyses, which adjusted for all the factors in Table 1, the adjusted probability of death within 30 days of admission at the reference category (as in Table 1) varied three-fold by hospital (0.58-1.75%; median: 1.12%, IQR: 0.96-1.28%). Antibiotic consumption measured as mean total DDDs/1,000 bed-days (2014-2016) varied 15-fold across hospitals (median: 1,814; IQR: 1,624-2,080; range: 266-4,006), with a 13-fold difference when expressed as DDDs/1,000 admissions (median: 4,132; IQR: 3,604-4,766; range: 584-7,494). Wide variation was also observed for other antibiotic metrics (Figure 2), including broad-spectrum DDDs/1,000 bed-days (median: 520; IQR: 389-650; range: 88-1,269) and per 1,000 admissions (median: 1,185; IQR: 925-1,443; range: 193-2,387), parenteral DDDs/1,000 bed-days (median: 437; IQR: 380-496; range: 51-729) and per 1,000 admissions (median: 971; IQR: 845-1124; range: 112-1893), and ‘Reserve’ DDDs/1,000 bed-days (median: 40; IQR: 29-57; range: 5-172) and per 1,000 admissions (median: 88; IQR: 63-129; range: 12-350). Ten antibiotics accounted for the majority prescribed (63.36-85.10%) at every acute hospital, but there was also considerable variation in the use of individual agents. For example, in descending order, co-amoxiclav accounted for between 1.7-29.4% of total use among hospitals in 2016, followed by doxycycline (5.0-35.8%), flucloxacillin (4.2-18.2%), clarithromycin (1.4-25.5%), amoxicillin (1.8-20.3%), metronidazole (1.8-7.7%), ciprofloxacin (1.0-14.4%), trimethoprim (0.6-8.4%), piperacillin/tazobactam (0.5-8.7%), and azithromycin (0.3-17.0%) (Appendix Figure A3).

**Figure 2:**
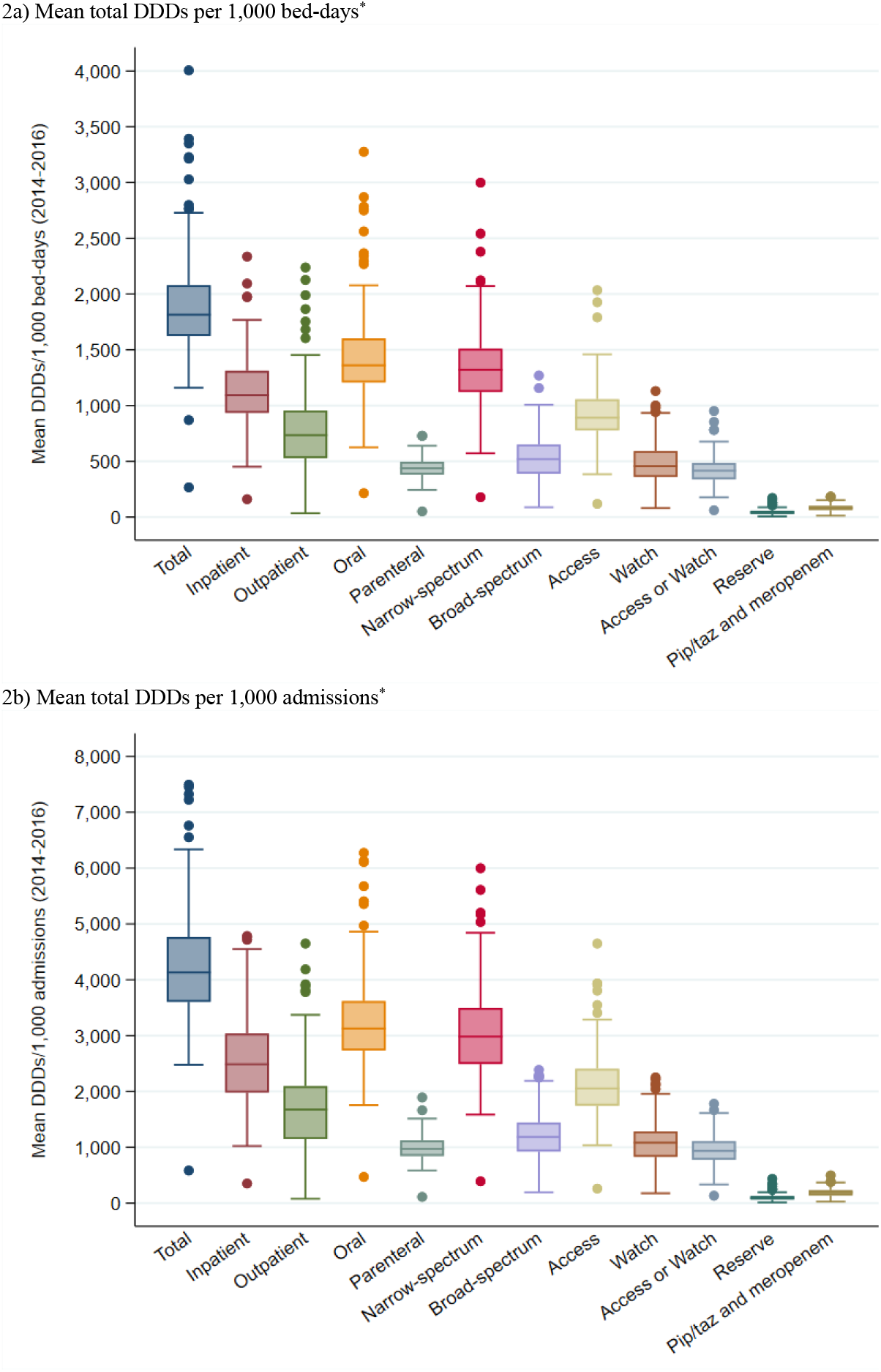
Mean Antibiotic Defined-Daily-Doses Consumed Among 135 NHS Acute Care Hospital Trusts (April 2014 – March 2017) * Each box represents the interquartile range of the distribution and is subdivided by a horizontal line representing the median. The ends of the whiskers display the most extreme DDD estimate within 1.5 IQR of the nearest quartile, while even more extreme outliers are displayed as isolated points. “Pip/taz” refers to piperacillin/tazobactam. Some antibiotics may be considered Access or Watch depending on indication, so they have been included as their own separate category.

Meta-regression estimates are displayed in Figure 3 and Appendix Table A6, with associations between the four main measures of antibiotic consumption (total, parenteral, broad-spectrum, ‘Reserve’ DDDs) per 1000 bed-days and 30-day mortality in Figure 4 (other meta-regression plots for 30-day mortality in Appendix Figure A4). In 22/24 meta-regression models we found evidence of no association between the adjusted probability of death and hospital-level antibiotic use. The two models with some evidence of identified effects in opposite directions; for each increase of 500 oral DDDs per 1000 admissions we observed a decrease in adjusted probability of death of −0.028% (95% CI: −0.053,-0.003; p=0.028), while for each increase of 500 parenteral DDDs per 1000 bed-days we observed an increase in the adjusted probability of death of +0.284% (95% CI: +0.031,+0.538; p=0.028).

**Figure 3:**
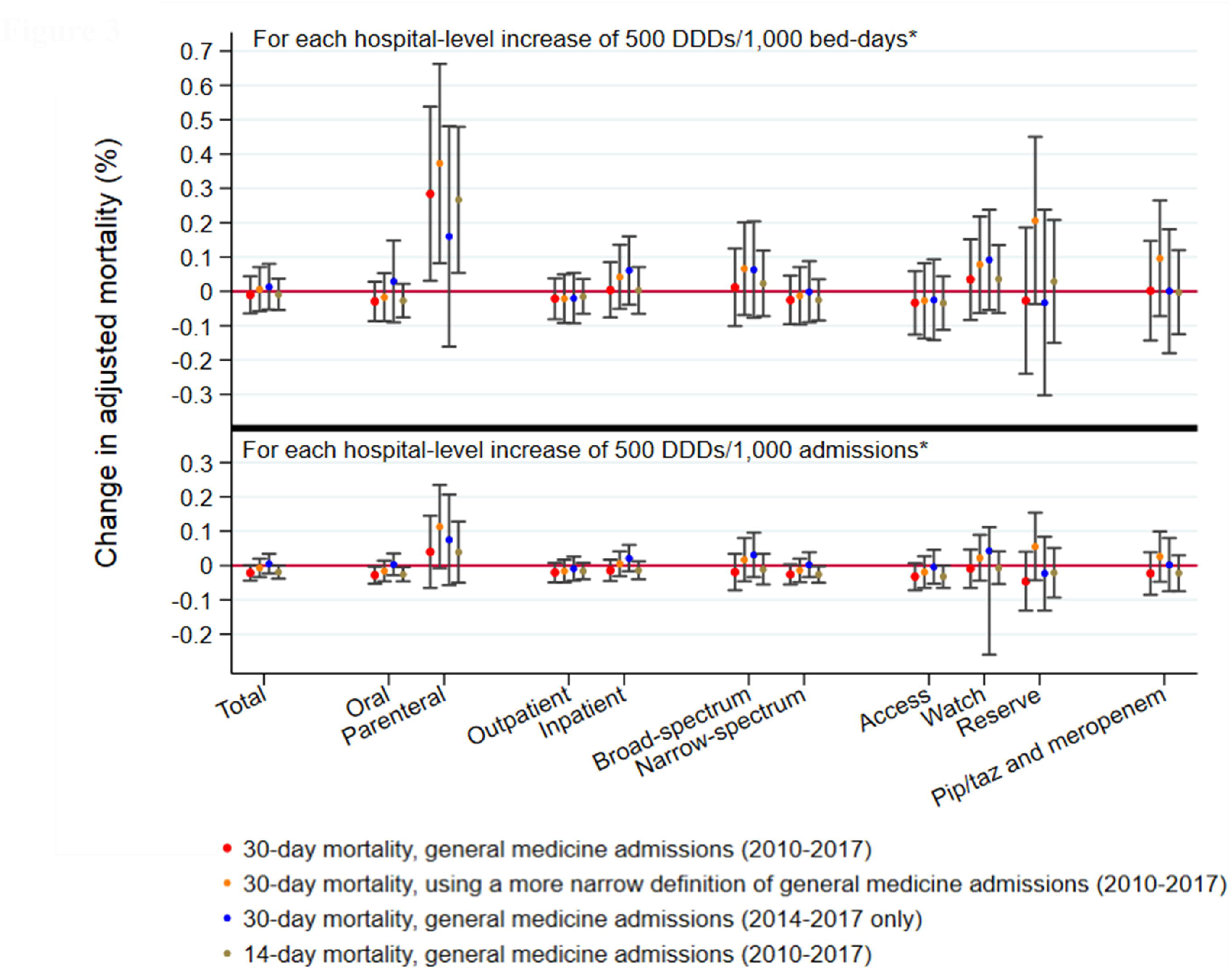
Random Effects Meta-Regression of the Association Between the Adjusted Probability of Death (In/Out of Hospital) and Different Metrics of Hospital-level Antibiotic Use. * Point estimates above the null (red line) suggest increasing hospital-level antibiotic use is associated with harm (increased mortality risk), or conversely that decreasing hospital-level antibiotic use is associated with clinical benefit (reduced mortality risk). Estimates below the null suggest increasing antibiotic use is associated with clinical benefit, or conversely that decreasing antibiotic use is associated with clinical harm. The associations displayed for Reserve antibiotics and piperacillin/tazobactam (“pip/taz”) and meropenem are for a unit increase of 100 DDDs rather than 500 DDDs. Some antibiotics (not shown) may be considered either Access or Watch (depending on indication) and have been analysed as their own separate category in Appendix Tables A6–A10 alongside the categories shown here.

**Figure 4:**
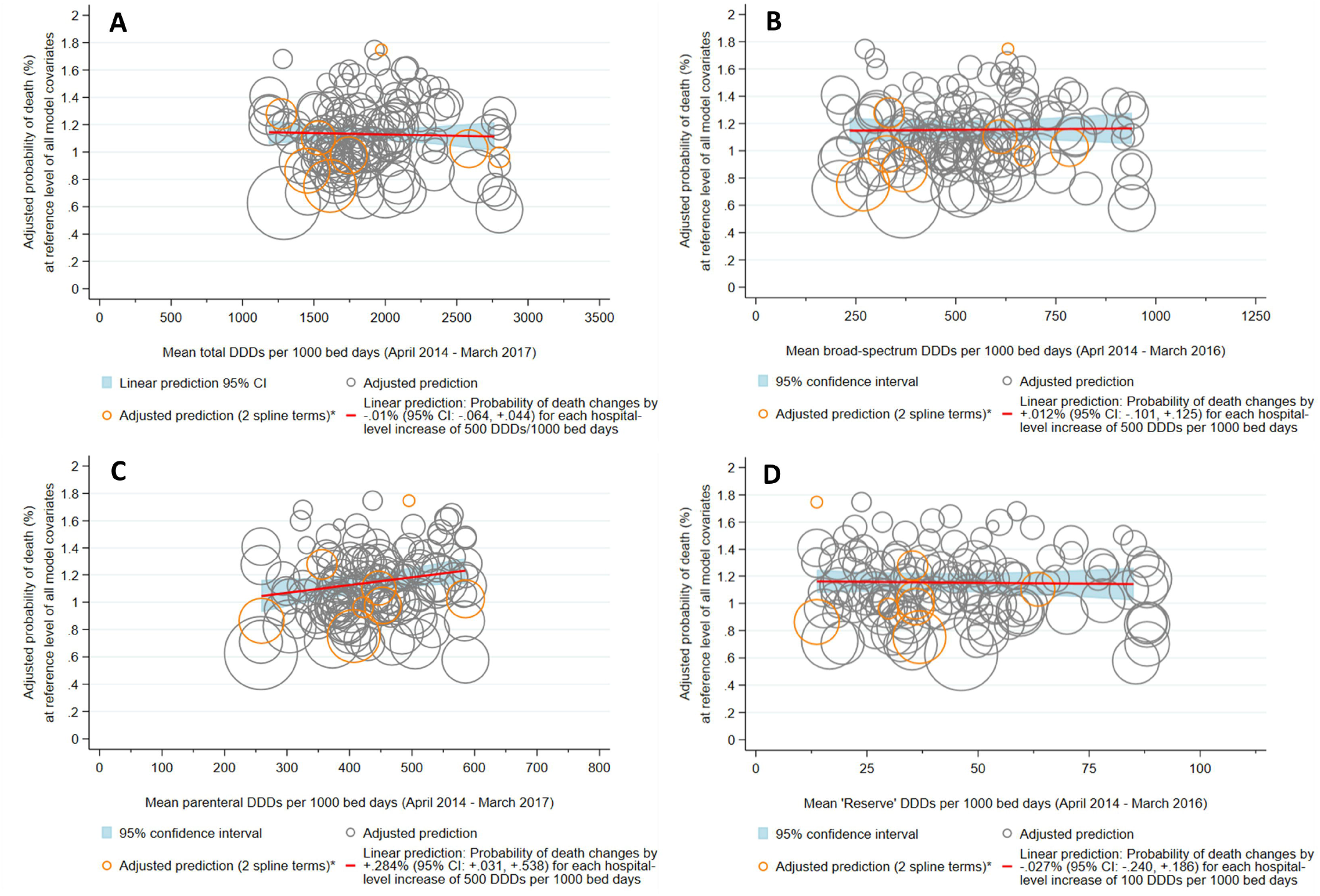
Random Effects Meta-Regression of the Association Between the Adjusted Probability of Death Within 30 Days of Admission (In/Out of Hospital) and Hospital-level Antibiotic Use. A) Mean total DDDs/1,000 bed-days B) Mean broad-spectrum DDDs/1,000 bed-days C) Mean parenteral DDDs/1,000 bed-days D) Mean “Reserve” DDDs/1,000 bed-days * Marginal effects were derived from 135 separate (hospital-specific) models, represented here as circles sized according to the precision of the estimate (inverse of the within-hospital variance). Most (127/135) hospital-specific multivariate models included four spline terms (for age, Charlson Comorbidity Index, IMD score, and overnight admissions in the past year), however models for eight hospitals would only converge with two spline terms (age and overnight admissions in the past year). Antibiotic use was truncated below the 2.5^th^ percentile and above the 95^th^ percentile. Probability estimates were truncated above the 99^th^ percentile. Associations displayed here can be found in Table 2.

### Scope for achieving antibiotic prescribing reductions

If hospitals with antibiotic use above the 25^th^ percentile (1,624 DDDs/1,000 bed-days; 101 hospitals), 10^th^ percentile (1,454 DDDs/1,000 bed-days; 121 hospitals), or 5^th^ percentile (1,282 DDDs/1,000 bed-days; 128 hospitals) reduced their consumption to this level, total DDD use would decline by 21.6% (from 51,732,671 DDDs to 40,558,491 DDDs), 27.0% (from 58,838,197 DDDs to 42,953,040 DDDs), or 34.4% (from 61,250,673 DDDs to 40,153,594), respectively. With antibiotic use measured in DDDs/1,000 admissions, reducing consumption to the 25^th^ percentile (3,604 DDDs/1,000 admissions), 10^th^ percentile (3,198 DDDs/1,000 admissions), or 5^th^ percentile (2,992 DDDs/1,000 admissions) would drop total DDD use by 23.2%, 28.4%, and 31.8%, respectively.

### Sensitivity analyses

Using a narrower definition of general medicine reduced the cohort from 36,124,372 to 19,144,018 admissions. To improve model stability, five hospitals with fewer than 50,000 admissions were then excluded (as in the primary analysis; Appendix Figure A1), leaving an analytic cohort with 19,023,144 admissions from 130 hospitals. The adjusted probability of death within 30 days of admission varied between 0.59-2.10% across hospitals (median: 1.11%, IQR: 0.96-1.33%) at the reference level of all model factors. In 23/24 meta-regression models there was no evidence of association between the adjusted probability of death and hospital-level antibiotic use (Figure 3 and Appendix Table A7). Again we observed that for each increase of 500 parenteral DDDs per 1000 bed-days there was a small increase in the adjusted probability of death of +0.373% (95% CI: +0.082,+0.663; p=0.012).

Restricting the study sample to admissions between April 2014-March 2017 inclusive (the period with overlapping antibiotic data) excluded 19,309,765 spells. Four hospitals with fewer than 50,000 admissions were then excluded, as were two hospitals with too few admissions in the reference group of admission specialty for multivariate models to converge. The resulting analytic cohort contained 16,492,990 spells from 129 hospitals. There was no evidence of association between the adjusted probability of death and hospital-level antibiotic use, regardless of how antibiotic use was measured (Figure 3 and Appendix Table A8).

### Secondary outcomes

The adjusted probability of death within 14 days of admission (in/out of hospital) varied from 0.38-1.40% across hospitals (median: 0.79%, IQR: 0.65-0.93%) at the reference level of all model factors. Most (20/24) metrics of antibiotic use showed no evidence of association with 14-day mortality, while the remaining estimates showed both positive and negative associations (Figure 3 and Appendix Table A9). The adjusted probability of non-elective re-admission to hospital within 30 days of discharge varied between 7.07-13.59% across hospitals (median: 10.16%, IQR: 9.34-10.86%) at the reference level of all model factors. Some (14/24) metrics of antibiotic use suggested re-admission risk increased with greater hospital-level antibiotic use, while the remainder (10/24) showed no evidence of association with re-admission risk (Appendix Table A10).

## DISCUSSION

We found very wide variation in the quantity of antibiotics being consumed across NHS acute hospitals, which is consistent with previous reports,(36) including a systematic review of antibiotic consumption in 3130 (primarily European) hospitals which found a 40-fold difference among studies.(35) By calculating the confounding-adjusted probability of death among general medicine inpatients in each hospital we have been able to control extensively for case-mix and found no evidence that variation in antibiotic use is associated with 14 or 30-day mortality. This finding is consistent across different measures of antibiotic consumption and in multiple sensitivity analyses focusing on sub-populations. These results indicate that at many hospitals patients are receiving considerably more antibiotics than necessary to treat their acute infections. Accordingly, substantial reductions in antibiotic use should be achievable without harming patients from under-treatment. Rather than set an arbitrary threshold for what reduction in use could be achieved, we have considered the impact of reducing hospital antibiotic use to the 25^th^, 10^th^ and 5^th^ centiles among hospitals. Depending on the threshold, our models indicate that system-wide reductions of up to one-third of DDDs could be achieved safely if high using hospitals could replicate prescribing practices of lower using hospitals.

Safe control of antibiotic overuse in hospital depends on balancing the need to initiate prompt effective therapy when bacterial infection is present with early review and revision of antibiotic prescriptions in the light of clinical and diagnostic data.(21,22) This is challenging in clinical practice. We have reported previously that hospitals which do this well have lower rates of *C. difficile* infection.(34) It is to be expected that at hospitals where antibiotic prescription reviews are done well, fewer antibiotics would be used without compromising patient outcomes. Data from medical inpatients in Oxford showed substantial reductions in antibiotic use (of around 30%) can be achieved without adverse clinical outcomes when patients are admitted under an infectious diseases specialist versus other clinical teams.(23) The most likely explanation for our observation therefore is that some hospitals are much more effective than others at implementing antibiotic review and revise in line with best practice. This is in keeping with a growing body of evidence that reducing antibiotic treatment duration across a wide-range of clinical scenarios is a safe and effective way of controlling antibiotic overuse.(53–57) Our data indicate the magnitude of reductions that could be safely achieved, dwarfing the 1% year-on-year reductions required of NHS hospitals.(37)

Our study has important limitations. Of particular concern is unmeasured confounding. Patient-level factors significantly associated with mortality risk such as baseline haematology and biochemistry test results and admission time of day are not available in HES.(50) Any residual bias arising because hospitals with more acutely unwell patients are likely to use more antibiotics and to have patients at greater risk of death could mean we fail to detect harm associated with low antibiotic use. Some of our analyses indicated greater use of parenteral antibiotics (per 1,000 bed-days) was associated with increased risk of death, which is consistent with residual confounding after adjustment. Nevertheless, the magnitude of differences in antibiotic use we have observed and the marginal impact on mortality means residual confounders would have to be exerting a very large effect to meaningfully change our inferences.

Although we find no evidence that reduced antibiotic use is associated with increased risk of readmission to hospital, this result should be interpreted with caution. More than half of our analyses using different measures of antibiotic consumption indicated increased antibiotic use to be associated with greater risk of non-elective readmission. This is difficult to explain biologically and is mostly likely because our model factors, selected *a priori* based on previous analyses to adjust for case-mix in mortality models,(50) are not sufficient to control for confounding of the relationship between antibiotic use and readmission. It may be that some hospitals which use more antibiotics also discharge more quickly, and there is evidence from the United States indicating patients discharged against medical advice have higher readmission rates.(58) Nevertheless, it does not undermine our fundamental observation that there is no evidence lower antibiotic use is associated with increased risk of death.

Another important limitation of this study is that marginal effects were derived from outcomes in acute/general medicine admissions between 2010-2017, yet antibiotic data was not available at the specialty-level during this period or at all before April 2014. As a result, the share of DDDs attributable to general medical inpatients likely varies by hospital. The drawbacks of the available antibiotic data are off-set by our broad definition of general medicine (Figure 1) and the fact that general medical inpatients are the largest consumers of non-prophylactic antibiotics in hospitals.(59) Also, sensitivity analyses restricting to April 2014-March 2017 similarly found no evidence of association between 30-day mortality and hospital-level antibiotic use. The antibiotic data for 2014-2017 was only received in January 2020; most hospitals use was fairly stable over this period, supporting our comparison of average effects.

While the use of DDDs as a metric of antibiotic consumption has the advantage of enabling standardised comparisons of hospital antibiotic use without the need for patient-level data, it also has drawbacks.(60) For example, to reduce the selective pressure for resistance, antibiotic stewardship initiatives may encourage switching from the use of one broad-spectrum agent to combinations of narrow-spectrum agents. This would increase DDDs for treatment of the same indication, producing an apparent increase in antibiotic use. Differences may also exist between prescribed doses and WHO’s DDD reference values, leading to overestimates or underestimates of antibiotic use, depending on local treatment guidelines. Unfortunately, linkage of clinical data with electronic prescribing data is not yet widely available in England.

Other limitations stem from the underlying HES datasets. For example, the diagnosis (ICD-10) codes used to derive Charlson Comorbidity Index, CCS groups, and immunosuppression status are captured in HES by clinical coding departments using discharge summaries. This process is both standardised and audited but primarily serves administrative and reimbursement purposes and describes the main condition managed in an episode, which may be ruled out at a later date or be unrelated to the clinical indication for antibiotics. Adjusting for these factors could therefore use the future to predict the past in patients with long single-consultant admissions.

Despite these limitations, electronic health record studies such as ours allow the impact of antibiotic therapy on patient outcomes to be assessed on a scale that cannot be achieved with other approaches. Where richer patient-level antibiotic data are available, investigations could analyse inappropriate prescribing, additional outcomes, and the use of alternate measures of antibiotic use that mitigate the limitations of DDDs, such as length of therapy (LOT) (days between first and last administered antibiotic inclusive) or days of therapy (DOT) (the sum of days between first and last administered antibiotic inclusive, with each antibiotic counted separately).(60)

Future observational work could also expand outcome measures to include other potential markers of harm, including length of stay or admission to intensive care. By using HES data from NHS Digital we have not been able to assess potential benefits of lower antibiotic prescribing, such as declines in AMR or *C. difficile* infection. Such data would need to either be obtained from individual hospitals, or only analysed at a hospital level (rather than within general medical inpatients) by using mandatory reported surveillance data.

## Conclusion

We found no evidence that the very wide variation in hospital antibiotic prescribing is associated with mortality risk in medical inpatients. Accordingly, risk-adjusted benchmarking of antibiotic use in hospitals could be used to drive safe and substantial reductions in antibiotic consumption. Further investigations should consider how some hospitals achieve low levels of antibiotic use. Understanding what explains these differences will facilitate the design of interventions that can be evaluated in randomised trials for unbiased inference. Although trials are costly and difficult to implement, the results of this study provide evidence to justify this further work.

## Data Availability

A copy of the Health Episode Statistics (HES) Admitted Patient Care dataset used in this study can be requested from NHS Digital via their Data Access Request Service (DARS) by using the same inclusion criteria and admission dates described in this study. NHS Digital can also be asked (via DARS) to link the requested HES data with death information from the Office of National Statistics (ONS). Antibiotic consumption data by NHS Trust can be requested from IQVIA Solutions UK Limited and its affiliates. Copyright: IQVIA Solutions UK Limited and its affiliates. All rights reserved. Use for sales, marketing or any other commercial purposes is not permitted without express prior written consent from IQVIA Solutions UK Limited.

## ACKNOWLEDGEMENTS

This work was supported by the National Institute for Health Research (NIHR) under its Programme Grants for Applied Research Programme (Reference Number RP-PG-0514-20015), and by Antibiotic Research UK (ANTRUK) grant number ANTSRG 02/2018. ASW is supported by the NIHR Biomedical Research Centre, Oxford. ASW and TEAP are NIHR Senior Investigators. The views expressed in this publication are those of the author(s) and not necessarily those of ANTRUK, the NHS, the NIHR, or the Department of Health.

## DATA SHARING

A copy of the Health Episode Statistics (HES) Admitted Patient Care dataset used in this study can be requested from NHS Digital’s Data Access Request Service (DARS) by using the same inclusion criteria and admission dates described in this study. NHS Digital can also be asked (via DARS) to link the requested HES data with death information from the Office of National Statistics (ONS).

Antibiotic consumption data by NHS Trust can be requested from IQVIA Solutions UK Limited and its affiliates. Copyright: IQVIA Solutions UK Limited and its affiliates. All rights reserved. Use for sales, marketing or any other commercial purposes is not permitted without IQVIA Solutions UK Limited’s express prior written consent.

## COPYRIGHT / LICENCE FOR PUBLICATION

The Corresponding Author has the right to grant on behalf of all authors and does grant on behalf of all authors, a world-wide licence to the Publishers and its licencees in perpetuity, in all forms, formats and media (whether known now or created in the future), to i) publish, reproduce, distribute, display and store the Contribution, ii) translate the Contribution into other languages, create adaptations, reprints, include within collections and create summaries, extracts and/or, abstracts of the Contribution, iii) create any other derivative work(s) based on the Contribution, iv) to exploit all subsidiary rights in the Contribution, v) the inclusion of electronic links from the Contribution to third party material where—ever it may be located; and, vi) licence any third party to do any or all of the above.

## CONTRIBUTOR AND GUARANTOR INFORMATION

EPB supported the study conceptualisation and funding acquisition, revised the statistical analysis plan, led the formal analysis of the data, wrote the first draft of the manuscript, and revised the paper. He is the guarantor. EPB attests that all listed authors meet authorship criteria and that no others meeting the criteria have been omitted.

TJD provided advice during the analysis and interpretation of results, and revised the paper.

TD provided advice during the analysis and interpretation of results, and revised the paper.

SH obtained data, provided advice during the analysis and interpretation of results, and revised the paper.

DW provided advice during the interpretation of results and revised the paper.

TP conceived of the study, provided advice during the analysis and interpretation of results, and revised the paper.

MG provided advice during the interpretation of results and revised the paper.

MJL conceived of the study, provided advice during the analysis and interpretation of results, and revised the paper.

ASW conceived of the study, obtained data, wrote the first draft of the statistical analysis plan, provided advice during the analysis and interpretation of results, and revised the paper.

## TRANSPARANCY STATEMENT

The manuscript’s guarantor (EPB) affirms that this manuscript is an honest, accurate, and transparent account of the study being reported; that no important aspects of the study have been omitted; and that any discrepancies from the study as planned (and, if relevant, registered) have been explained

## ETHICS APPROVAL

This study is an analysis of de-identified routine electronic health record data obtained through NHS Digital’s Data Access Request Service (DARS). The original study protocol was approved by the Health and Social Care Information Centre (HSCIC; now NHS Digital) on 8 June 2016. At that time, guidance provided by NHS Digital, the Medical Research Council (MRC), and the NHS Health Research Authority (HRA) advised that the use of non-identifiable data from NHS Digital did not require HRA Research Ethics Committee (REC) review. In line with this guidance, REC review and university ethics review were not obtained. HES data for the project was finally received in January 2018. The hospital-level antibiotic data used in this study was obtained from IQVIA (via Public Health England) in January 2020 and contains no patient-level information.

## REFERENCES

1. Costelloe C, Metcalfe C, Lovering A, Mant D, Hay AD. Effect of antibiotic prescribing in primary care on antimicrobial resistance in individual patients: systematic review and metaanalysis. BMJ. 2010 May 18;340:c2096. Available from: http://www.ncbi.nlm.nih.gov/pubmed/20483949

2. Goossens H, Ferech M, Vander Stichele R, Elseviers M, ESAC Project Group. Outpatient antibiotic use in Europe and association with resistance: a cross-national database study. Lancet. 2005 Feb;365(9459):579–87. Available from: http://www.ncbi.nlm.nih.gov/pubmed/15708101

3. de Kraker MEA, Davey PG, Grundmann H, BURDEN study group. Mortality and hospital stay associated with resistant Staphylococcus aureus and Escherichia coli bacteremia: estimating the burden of antibiotic resistance in Europe. Opal SM, editor. PLoS Med. 2011 Oct 11;8(10):e1001104. Available from: http://dx.plos.org/10.1371/journal.pmed.1001104

4. Bell BG, Schellevis F, Stobberingh E, Goossens H, Pringle M. A systematic review and metaanalysis of the effects of antibiotic consumption on antibiotic resistance. BMC Infect Dis. 2014 Dec 9;14(1):13. Available from: http://www.ncbi.nlm.nih.gov/pubmed/24405683

5. Chatterjee A, Modarai M, Naylor NR, et al. Quantifying drivers of antibiotic resistance in humans: a systematic review. Lancet Infect Dis. 2018 Dec;18(12):e368–78. Available from: https://linkinghub.elsevier.com/retrieve/pii/S1473309918302962

6. Tamma PD, Avdic E, Li DX, Dzintars K, Cosgrove SE. Association of Adverse Events With Antibiotic Use in Hospitalized Patients. JAMA Intern Med. 2017 Sep 1;177(9):1308. Available from: http://www.ncbi.nlm.nih.gov/pubmed/28604925

7. Tennyson L, Averch T. An update on fluoroquinolones: the emergence of a multisystem toxicity syndrome. Urol Pract. 2017;4(5):386–7.

8. Gao Z, Chen Y, Guan M-X. Mitochondrial DNA mutations associated with aminoglycoside induced ototoxicity. J Otol. 2017 Mar;12(1):1–8. Available from: http://www.ncbi.nlm.nih.gov/pubmed/29937831

9. Kalghatgi S, Spina CS, Costello JC, et al. Bactericidal Antibiotics Induce Mitochondrial Dysfunction and Oxidative Damage in Mammalian Cells. Sci Transl Med. 2013 Jul 3;5(192):192ra85–192ra85. Available from: http://www.ncbi.nlm.nih.gov/pubmed/23825301

10. Blaser MJ. Antibiotic use and its consequences for the normal microbiome. Science. 2016 Apr 29;352(6285):544–5. Available from: http://www.ncbi.nlm.nih.gov/pubmed/27126037

11. Bhalodi AA, van Engelen TSR, Virk HS, Wiersinga WJ. Impact of antimicrobial therapy on the gut microbiome. J Antimicrob Chemother. 2019 Jan 1;74(Suppl 1):i6–15. Available from: https://academic.oup.com/jac/article/74/Supplement_1/i6/5300216

12. Malik U, Armstrong D, Ashworth M, et al. Association between prior antibiotic therapy and subsequent risk of community-acquired infections: a systematic review. J Antimicrob Chemother. 2018 Feb 1;73(2):287–96. Available from: http://www.ncbi.nlm.nih.gov/pubmed/29149266

13. Brown KA, Khanafer N, Daneman N, Fisman DN. Meta-Analysis of Antibiotics and the Risk of Community-Associated Clostridium difficile Infection. Antimicrob Agents Chemother. 2013 May;57(5):2326–32. Available from: http://aac.asm.org/lookup/doi/10.1128/AAC.02176-12

14. Deshpande A, Pasupuleti V, Thota P, et al. Community-associated Clostridium difficile infection and antibiotics: a meta-analysis. J Antimicrob Chemother. 2013 Sep;68(9):1951–61. Available from: https://academic.oup.com/jac/article-lookup/doi/10.1093/jac/dkt129

15. Slimings C, Riley T V. Antibiotics and hospital-acquired Clostridium difficile infection: update of systematic review and meta-analysis. J Antimicrob Chemother. 2014 Apr;69(4):881–91. Available from: http://www.ncbi.nlm.nih.gov/pubmed/24324224

16. D’Atri F, Arthur J, Blix HS, Hicks LA, Plachouras D, Monnet DL. Targets for the reduction of antibiotic use in humans in the Transatlantic Taskforce on Antimicrobial Resistance (TATFAR) partner countries. Euro Surveill. 2019 Jul 1;24(28). Available from: /pmc/articles/PMC6636213/?report=abstract

17. HM Government. Tackling antimicrobial resistance 2019–2024: the UK’s five-year national action plan. 2019. Available from: https://www.gov.uk/government/publications/uk-5-year-action-plan-for-antimicrobial-resistance-2019-to-2024

18. Public Health England. English surveillance programme for antimicrobial utilisation and resistance (ESPAUR) Report 2018-2019. 2018. Available from: https://www.gov.uk/government/publications/english-surveillance-programme-antimicrobial-utilisation-and-resistance-espaur-report

19. Rhee C, Kadri SS, Dekker JP, et al. Prevalence of Antibiotic-Resistant Pathogens in Culture-Proven Sepsis and Outcomes Associated With Inadequate and Broad-Spectrum Empiric Antibiotic Use. JAMA Netw open. 2020 Apr 1;3(4):e202899. Available from: /pmc/articles/PMC7163409/?report=abstract

20. Levy MM, Dellinger RP, Townsend SR, et al. The Surviving Sepsis Campaign: Results of an international guideline-based performance improvement program targeting severe sepsis. Crit Care Med. 2010 Feb;38(2):367–74. Available from: http://www.ncbi.nlm.nih.gov/pubmed/20035219

21. Ashiru-Oredope D, Sharland M, Charani E, McNulty C, Cooke J, ARHAI Antimicrobial Stewardship Group. Improving the quality of antibiotic prescribing in the NHS by developing a new Antimicrobial Stewardship Programme: Start Smart--Then Focus. J Antimicrob Chemother. 2012 Jul 1;67(suppl 1):i51–63. Available from: http://www.ncbi.nlm.nih.gov/pubmed/22855879

22. Centers for Disease Control and Prevention. Core Elements of Hospital Antibiotic Stewardship Programs. Atlanta, GA: US Department of Health and Human Services, CDC. 2019. Available from: https://www.cdc.gov/antibiotic-use/core-elements/hospital.html#_ENREF_4

23. Fawcett NJ, Jones N, Quan TP, et al. Antibiotic use and clinical outcomes in the acute setting under management by an infectious diseases acute physician versus other clinical teams: a cohort study. BMJ Open. 2016 Aug 23;6(8):e010969. Available from: http://bmjopen.bmj.com/lookup/doi/10.1136/bmjopen-2015-010969

24. Versporten A, Zarb P, Caniaux I, et al. Antimicrobial consumption and resistance in adult hospital inpatients in 53 countries: results of an internet-based global point prevalence survey. Lancet Glob Heal. 2018;6(6):e619–29. Available from: https://www.ncbi.nlm.nih.gov/pubmed/29681513

25. Cross ELA, Sivyer K, Islam J, et al. Adaptation and implementation of the ARK (Antibiotic Review Kit) intervention to safely and substantially reduce antibiotic use in hospitals: a feasibility study. J Hosp Infect. 2019 Nov;103(3):268–75. Available from: http://www.ncbi.nlm.nih.gov/pubmed/31394146

26. Islam J, Ashiru-Oredope D, Budd E, et al. A national quality incentive scheme to reduce antibiotic overuse in hospitals: Evaluation of perceptions and impact. J Antimicrob Chemother. 2018 Jun 1;73(6):1708–13.

27. Powell N, McGraw-Allen K, Menzies A, Peet B, Simmonds C, Wild A. Identifying antibiotic stewardship interventions to meet the NHS England CQUIN: An evaluation of antibiotic prescribing against published evidence-based antibiotic audit tools. Clin Med (Lond). 2018 Aug 1;18(4):276–81. Available from: /pmc/articles/PMC6334038/?report=abstract

28. Braykov NP, Morgan DJ, Schweizer ML, et al. Assessment of empirical antibiotic therapy optimisation in six hospitals: An observational cohort study. Lancet Infect Dis. 2014;14(12):1220–7. Available from: https://pubmed.ncbi.nlm.nih.gov/25455989/

29. NHS England. Commissioning for Quality and Innovation (CQUIN) Guidance for 2016/17. 2016. Available from: https://www.england.nhs.uk/nhs-standard-contract/cquin/cquin-16-17/

30. Hood G, Hand KS, Cramp E, Howard P, Hopkins S, Ashiru-Oredope D. Measuring appropriate antibiotic prescribing in acute hospitals: Development of a national audit tool through a Delphi consensus. Antibiotics. 2019 Jun 1;8(2). Available from: https://pubmed.ncbi.nlm.nih.gov/31035663/

31. Fridkin SK, Lawton R, Edwards JR, Tenover FC, McGowan JE, Gaynes RP. Monitoring antimicrobial use and resistance: Comparison with a national benchmark on reducing vancomycin use and vancomycin-resistant enterococci. Emerg Infect Dis. 2002;8(7):702–7. Available from: https://pubmed.ncbi.nlm.nih.gov/12095438/

32. Ibrahim OM, Polk RE. Benchmarking antimicrobial drug use in hospitals. Expert Rev Anti Infect Ther. 2012 Apr;10(4):445–57. Available from: https://pubmed.ncbi.nlm.nih.gov/22512754/

33. Kuster SP, Ruef C, Bollinger AK, et al. Correlation between case mix index and antibiotic use in hospitals. J Antimicrob Chemother. 2008;62(4):837–42. Available from: https://pubmed.ncbi.nlm.nih.gov/18617509/

34. Llewelyn MJ, Hand K, Hopkins S, Sarah Walker A. Antibiotic policies in acute English NHS trusts: Implementation of “Start Smart-Then Focus” and relationship with Clostridium difficile infection rates. J Antimicrob Chemother. 2014 Sep 16;70(4):1230–5.

35. Bitterman R, Hussein K, Leibovici L, Carmeli Y, Paul M. Systematic review of antibiotic consumption in acute care hospitals. Clin Microbiol Infect. 2016 Jun 1;22(6):561.e7–561.e19.

36. Zanichelli V, Monnier A, Gyssens I, et al. Variation in Antibiotic Use Among and Within Different Settings: A Systematic Review. J Antimicrob Chemother. 2018;73(suppl_6):vi17–29. Available from: https://pubmed.ncbi.nlm.nih.gov/29878219/

37. NHS England. NHS Standard Contract 2019/20 Service Conditions. 2019. Available from: https://www.england.nhs.uk/publication/nhs-standard-contract-2019-20-service-conditions-full-length/

38. Herbert A, Wijlaars L, Zylbersztejn A, Cromwell D, Hardelid P. Data Resource Profile: Hospital Episode Statistics Admitted Patient Care (HES APC). Int J Epidemiol. 2017;46(4):1093–1093i. Available from: http://www.ncbi.nlm.nih.gov/pubmed/28338941

39. Health and Social Care Information Centre. Methodology for Creation of the HES Patient ID (HESID). 2014. Available from: http://content.digital.nhs.uk/media/1370/HES-Hospital-Episode-Statistics-Replacement-of-the-HES-patient-ID/pdf/HESID_Methodology.pdf

40. IQVIA. Welcome to IQVIA. Available from: https://www.iqvia.com/

41. WHO Collaborating Centre for Drug Statistics Methodology. International language for drug utilization research. 2019. Available from: https://www.whocc.no/

42. Public Health England. MRSA bacteraemia: annual data. Annual counts and rates of meticillin resistant Staphylococcus aureus (MRSA) bacteraemia by acute trust and clinical commissioning group (CCG). 2018. Available from: https://www.gov.uk/government/statistics/mrsa-bacteraemia-annual-data

43. NHS Digital. Hospital admitted patient care activity: provider level analysis. 2017. Available from: https://digital.nhs.uk/data-and-information/publications/statistical/hospital-admitted-patient-care-activity

44. NHS Digital. Provider spells methodology: provider mapping files October 2019. 2019. Available from: https://digital.nhs.uk/data-and-information/publications/ci-hub/summary-hospital-level-mortality-indicator-shmi

45. NHS Digital. HES Data Dictionary Admitted Patient Care. 2018. Available from: https://digital.nhs.uk/data-and-information/data-tools-and-services/data-services/hospital-episode-statistics/hospital-episode-statistics-data-dictionary

46. Benchimol EI, Smeeth L, Guttmann A, et al. The REporting of studies Conducted using Observational Routinely-collected health Data (RECORD) Statement. PLOS Med. 2015;12(10):e1001885. Available from: http://www.ncbi.nlm.nih.gov/pubmed/26440803

47. von Elm E, Altman DG, Egger M, Pocock SJ, Gøtzsche PC, Vandenbroucke JP. The Strengthening the Reporting of Observational Studies in Epidemiology (STROBE) statement: guidelines for reporting observational studies. Lancet. 2007;370(9596):1453–7. Available from: https://pubmed.ncbi.nlm.nih.gov/18064739/

48. Thompson S, Higgins J. How should meta-regression analyses be undertaken and interpreted? Stat Med. 2002;21(11):1559–73.

49. Zou G. A modified poisson regression approach to prospective studies with binary data. Am J Epidemiol. 2004 Apr 1;159(7):702–6. Available from: http://www.ncbi.nlm.nih.gov/pubmed/15033648

50. Walker AS, Mason A, Quan TP, et al. Mortality risks associated with emergency admissions during weekends and public holidays: an analysis of electronic health records. Lancet. 2017 Jul 1;390(10089):62–72. Available from: http://linkinghub.elsevier.com/retrieve/pii/S0140673617307821

51. Onukwugha E, Bergtold J, Jain R. A Primer on Marginal Effects—Part I: Theory and Formulae. Pharmacoeconomics. 2015 Jan 5;33(1):25–30. Available from: http://www.ncbi.nlm.nih.gov/pubmed/25189459

52. Budd E, Cramp E, Sharland M, et al. Adaptation of the WHO Essential Medicines List for national antibiotic stewardship policy in England: being AWaRe. J Antimicrob Chemother. 2019;74(11):3384–9. Available from: https://www.ncbi.nlm.nih.gov/pubmed/31361000

53. Llewelyn MJ, Fitzpatrick JM, Darwin E, et al. The antibiotic course has had its day. BMJ. 2017 Jul 26;358:j3418. Available from: http://www.ncbi.nlm.nih.gov/pubmed/28747365

54. Onakpoya I, Walker A, Tan P, et al. Overview of systematic reviews assessing the evidence for shorter versus longer duration antibiotic treatment for bacterial infections in secondary care. PLoS One. 2018;13(3):e0194858. Available from: https://www.ncbi.nlm.nih.gov/pubmed/29590188

55. Royer S, DeMerle KM, Dickson RP, Prescott HC. Shorter Versus Longer Courses of Antibiotics for Infection in Hospitalized Patients: A Systematic Review and Meta-Analysis. J Hosp Med. 2018 Jan 25;13(5):336–42. Available from: https://www.journalofhospitalmedicine.com/jhospmed/article/156407/hospital-medicine/shorter-versus-longer-courses-antibiotics-infection

56. Sawyer RG, Claridge JA, Nathens AB, et al. Trial of Short-Course Antimicrobial Therapy for Intraabdominal Infection. N Engl J Med. 2015 May 21;372(21):1996–2005. Available from: http://www.ncbi.nlm.nih.gov/pubmed/25992746

57. Uranga A, España PP, Bilbao A, et al. Duration of Antibiotic Treatment in Community-Acquired Pneumonia. JAMA Intern Med. 2016 Sep 1;176(9):1257. Available from: http://www.ncbi.nlm.nih.gov/pubmed/27455166

58. Safi W, Elnegouly M, Schellnegger R, et al. Infection and Predictors of Outcome of Cirrhotic Patients after Emergency Care Hospital Admission. Ann Hepatol. 2018 Oct;17(6):948–58.

59. Health Protection Agency. English national point prevalence survey on healthcare-associated infections and antimicrobial use, 2011. 2012. Available from: https://webarchive.nationalarchives.gov.uk/20140714085429/ http://www.hpa.org.uk/Publications/InfectiousDiseases/AntimicrobialAndHealthcareAssociatedInfections/1205HCAIEnglishPPSforhcaiandamu2011prelim/

60. Ibrahim OM, Polk RE. Antimicrobial use metrics and benchmarking to improve stewardship outcomes: methodology, opportunities, and challenges. Infect Dis Cinics North Am. 2014 Jun;28(2):195–214. Available from: http://www.ncbi.nlm.nih.gov/pubmed/24857388

