## Appendix for "An ecological comparison of hospital-level antibiotic use and mortality in 36,124,372 acute/general medicine inpatients in England"

### Appendix - Table of Contents

Table 5: Clinical Classifications Software (CCS) Diagnosis Group Identified among Acute/General Medical Admissions in 135 Acute Care NHS Hospital Trusts, England, April 2010 to March 201721

#### METHODS

##### Terminology

A “Trust” is an organisational unit within the NHS that may provide hospital services, community services, or may act as a commissioner when sub-contracting care to other health services providers. In this study, the term “Trust” is synonymous with acute hospital.

A provider “spell” reflects a period of care in a single hospital, starting at the time of admission and ending at the time of discharge, death, or transfer out. Within a spell an “episode” reflects a defined period of care under a consultant. Thus, a spell comprises one or more episodes.(1) Episodes which share a patient identification number (HESID), admission date, provider code, and hospital provider spell number together compose a provider spell.(2)

Bed days reported by PHE(3) are collected by consultant main specialties and include either beds that are occupied at midnight on wards that are open overnight or beds occupied by at least one day-case on day-only wards. The reporting excludes beds that have been commissioned from non-NHS providers, beds designated solely for use by well infants, critical care beds, and residential care beds.(4)

##### Identifying spells

Study authors Eric P. Budgell and A. Sarah Walker had access to the raw HES data extracts, which included 123,079,071 consultant episodes. Episodes were dropped if identified as perfect duplicates (n=14,672,193), duplicates across all fields except episode identifier number (epikey) (n=22,196), if admitted before 01 April 2009 (n=139,179), missing an admission date (n=5,804), missing a HESID (n=70,619), or missing a hospital provider spell number (n=340). Among the remaining 108,168,740 episodes, 88,718,419 unique spells were identified in 15,708,476 patients between 1 April 2009 and 31 March 2017. The remaining cleaning steps are outlined in Appendix Figure 1.

##### Imputing missing data

In the final analytic cohort (n=36,124,372 spells), the nearest preceding or subsequent spells were used to impute missing IMD score (n=83,642) (patients missing IMD score in all spells had already been dropped). Missing sex (n=1,547) was imputed to the mode (female), as was missing admission method (n=3,832) (mode: elective and other non-emergency), and missing admission source (n=36,848) (mode: usual/other place of residence).

##### Preparation of antibiotic data

Only systemic antibiotics were included in DDD estimates; thus eye drops and other topical agents (gels, creams) were excluded, as were pessaries, beads, bone cement, and non-absorbed antibiotics such as neomycin (ATC code: J01GB05). A small number of other agents were excluded for miscellaneous reasons, including methenamine (this is metabolised to formalin to suppress urinary tract infections, but is not an antibiotic), demeclocycline (this is an antibiotic but is used primarily to treat hyponatremia), and colistin (ATC code: A07AA10) which was very rarely used. Rifabutin was excluded due to its probable use in tuberculosis treatment; other antibiotics used primarily in the treatment of tuberculosis were excluded in advance by PHE.

Broad-spectrum antibiotics were defined as co-amoxiclav, piperacillin/tazobactam, second (e.g. cefuroxime), third (e.g. ceftriaxone, ceftazidime), fourth (e.g. cefepime) or fifth (e.g. ceftobiprole) generation cephalosporins, cephalosporin-beta-lactamase inhibitor combinations (e.g. ceftazidime-avibactam, ceftolozane-tazobactam), carbapenems, quinolones, azithromycin, tigecycline, aztreonam, telithromycin, and polymyxins (Appendix Table 3). Antibiotics not meeting this definition were classified as narrow-spectrum.

**Preparation of HES data and model selection**

Following HES guidance,(5) the primary diagnosis code was used to derive 142 CCS diagnosis groups, while secondary diagnosis fields were used to derive a Charlson Comorbidity Index. Following an approach reported in previous analyses,(6) the CCS groups were then aggregated into 43 subgroups based on clinical feedback, and categories with low (<0.5%) observed mortality were collapsed into a single 'low risk' group. To improve model stability, the smallest remaining categories (cumulatively accounting for 6.11% of all spells) were then collapsed into two 'other' categories with 30-day mortality risk above and below the overall mortality risk, respectively. This process yielded a total of 29 CCS categories shown in Appendix Table 5. A binary indicator of immunosuppression was defined from secondary diagnosis code indicating metastatic cancer, severe liver disease, or HIV.

The IMD score was provided by NHS Digital based on the 2010 English index of deprivation associated with the Super Output Area Level of the patient's residence (postal codes were not collected); a more recent 2015 deprivation index is available, but HES continues to use the 2010 index.(5)

The categories used to define primary admission specialty were based on the descending prevalence of consultant specialties. First, spells with a main specialty code (HES: mainspef) or treatment specialty code (HES: tretspef) of 300 in the first or second consultant episode were defined as acute/general medicine. If this code was not present then the next most common admission specialty code was used. This logic was then repeated for each of the specialties included in the sample population (Figure 1, main text). To improve model stability, individual admission specialties comprising <1 million spells were grouped as "other", including endocrinology, diabetic medicine, acute internal medicine, respiratory medicine, infectious diseases, neurology, and rheumatology. In the primary analysis (n=36,124,372 spells), Royal Surrey NHS Foundation Trust had very few admissions (n=182) in the reference group of admission specialty (general medicine) and yielded the highest adjusted probability estimate for 30-day mortality (2.96%, 95% CI: 0.84,5.07). In meta-regression models this point estimate was therefore truncated down to the 99$^{th}$ percentile.

Ethnic categories (HES: ethnos) were defined as white (ethnos="British (White)", "Irish (White)", "Any other White background"), Asian (ethnos="Indian (Asian or Asian British)", "Pakistani (Asian or Asian British)", "Bangladeshi (Asian or Asian British)", "Any other Asian background", "Chinese (other ethnic group)"), black (ethnos="Caribbean (Black or Black British)", "African (Black or Black British)", "Any other Black background"), other (ethnos="White and Black Caribbean (Mixed)", "White and Black African (Mixed)", "White and Asian (Mixed)", "Any other Mixed background", "Any other ethnic group"), and unknown (ethnos="Not stated", "Not known"). Admission method categories (HES: admimeth) were defined as A&E (admimeth="2A", "21"), elective and other non-emergency (admimeth="11", "12", "13", "25", "31", "32", "81", "82", "83"), or emergency via GP or other source ("22", "23", "24", "28", "2B", "2D"). Admission source categories (HES admisorc) were defined as usual/other place of residence (admisorc="19", "29", "39", "66", "79") or NHS general ward/other care provider (admisorc="49", "51", "52", "53", "54", "65", "85", "87", "88"). The definition of each admission method and source category can be found in the HES Admitted Patient Care data dictionary.(7) Intended management was collected from HES but not included in regression models as it was unknown in 53.8% (n=19,417,403) of spells.

Patient classification categories were collapsed, with "Regular night attender" (n=1,113) being combined with "Regular day attender", "Mothers and babies using only delivery facilities" (n=168) being combined with “Ordinary admission”, and “Day-case admission” being combined with “Ordinary admission” if length of stay was >1 day (n=92).

If no outcomes were observed in one of the categories of a model factor (for example, no deaths among “regular day attenders”), the category was combined with the mode for that hospital Trust only. One exception was the “low risk” category of CCS group which was combined with the “Other” CCS group where mortality risk was below the overall risk (Appendix Table 5). In the primary analysis of 30-day mortality, such changes were made to 13/135 hospital Trusts and affected only one or two variables where categories concerned had very few admissions (often <0.1% of total admissions within the hospital).

Substantial collinearity was observed between alternate measures of previous hospital exposure, including the number of overnight admissions in the past year versus ever (Spearman's ρ: 0.73, p<0.0001) and the number of complex overnight admissions in the past year versus ever (Spearman's ρ: 0.75, p<0.0001). Each measure was assessed in a single model adjusting for all hospital Trusts, as was a binary indicator for ever having had a complex admission in the past year. Factors were selected for inclusion in multivariate models based on minimizing the BIC in these single models, and included overnight admissions in the past year and any complex admission in the past year.

To improve model stability, age at admission (years), Charlson Comorbidity Index, and overnight admissions in the past year were truncated at the 99$^{th}$ percentile. To account for non-linear effects of continuous variables, age, Charlson score, IMD score, and overnight admissions in the past year were fit as natural cubic splines (Stata command: mkspline, cubic), as they lowered BIC of models adjusting for all hospital Trusts. The number of knots (up to six) was chosen based on BIC, with five Harrell knots(8) chosen for age, six Harrell knots chosen for IMD score, six knots chosen for Charlson score (with knots placed at the at the 10th, 35th, 50th, 65th, 80th, and 90th percentile of the non-zero distribution), and three knots chosen for overnight admissions in the past year (with knots placed at the 10th, 70th, and 90th percentile of the non-zero distribution) (Appendix Figure 2). In the latter two covariates, placement of knots at fixed intervals was not possible due to limited variation in the truncated distributions. Adjusted models retained the same number of knots, which remained significant (p<0.001) and were visualized to assess over-fitting.

Nine interaction terms included in previous analyses(6) were added individually to a single multivariate model including all hospital Trusts and found to significantly improve the model fit (lowered the BIC); these interactions were between age and Charlson score, admission method and calendar year, admission method and overnight admissions in the past year, admission specialty and overnight admissions in the past year, admission specialty and Charlson score, overnight admissions in the past year and Charlson score, ever had a complex overnight admission in the past year and Charlson score, overnight admissions in the past year and ever had a complex overnight admission in the last year, age and overnight admissions in the last year.

The 16 admission factors in Table 1 were adjusted for irrespective of statistical significance to ensure the best possible adjustment for case-mix when deriving marginal effects, rather than to assess the relative effect of factors within models, and these admission factors have been identified as predictors of mortality in previous large studies.(6,9) For the same reason, each model was fitted to data from each hospital Trust individually. To account for patients with repeated hospital admissions (observations which are not independent), a clustered version of the sandwich estimator was used, which affects the variance–covariance matrix of the estimators (including standard errors) but has no impact on estimated coefficients, thus yielding wider confidence bounds around point estimates than would be observed if all observations were assumed to be independent.

All marginal effects were derived in Stata using “nlcom” for nonlinear combination of estimators; results were compared with output from Stata's “margins” command, which was computationally slow but provided a reference check on all estimates and their standard errors. Random effects meta-regression was then performed in Stata using the "metareg" command.(10) This method is an extension of standard random effects meta-analysis(11–14) and is often performed on study-level summary data, allowing the underlying sources of statistical heterogeneity in effect estimates across hospital Trusts to be explored. The use of random-effects models accounts for the variance of hospital-level effects and residual (unexplained) sources of between-hospital heterogeneity, assuming hospital-level effects follow a normal distribution around the linear predictor.

For the primary mortality analysis, marginal effects were also derived using logistic regression.(15) These were very highly correlated (Spearman's $\rho$: 0.9992, $p<0.0001$) and similar in magnitude to estimates derived from Poisson models, so were not considered further.

#### APPENDIX TABLES

Table 1: Changes to HES Provider Codes Due to Splitting and Mergers Between NHS Hospital Trusts, April 2010 to March 2017

| **Old provider code** [i] | **Updated provider code** | **Financial year when change occurred** |
|---|---|---|
| Worthing and Southlands Hospitals NHS Trust (RPL), and Royal West Sussex NHS Trust (RPR) | Western Sussex Hospitals NHS Foundation Trust (RYR) | 2010/11 |
| Nuffield Orthopaedic Centre NHS Trust (RBF) | Oxford University Hospitals NHS Foundation Trust (RTH) | 2012/13 |
| Winchester and Eastleigh Healthcare NHS Trust (RN1) | Hampshire Hospitals NHS Foundation Trust (RN5) | 2012/13 |
| Trafford Healthcare NHS Trust (RM4) | Central Manchester University Hospitals NHS Foundation Trust (RW3) | 2012/13 |
| Scarborough and North East Yorkshire Health Care NHS Trust (RCC) | York Teaching Hospital NHS Foundation Trust (RCB) | 2012/13 |
| Newham University Hospital NHS Trust (RNH), and Whipps Cross University Hospital NHS Trust (RGC), and Barts and The London NHS Trust (RNJ) | Barts Health NHS Trust (R1H) | 2013/14 |
| South London Healthcare NHS Trust (RYQ) | King's College Hospital NHS Foundation Trust (RJZ) [ii] | 2014/15 |
| Mid Staffordshire NHS Foundation Trust (RJD) | University Hospitals of North Midlands NHS Trust (RJE) [ii] | 2014/15 |
| Ealing Hospital NHS Trust (RC3), and North West London Hospitals NHS Trust (RV8) | London North West Healthcare NHS Trust (R1K) | 2015/16 |
| Barnet and Chase Farm Hospitals NHS Trust (RVL) | Royal Free London NHS Foundation Trust (RAL) | 2015/16 |
| Royal National Hospital for Rheumatic Diseases NHS Foundation Trust (RBB) | Royal United Hospitals Bath NHS Foundation Trust (RD1) | 2015/16 |
| Heatherwood and Wexham Park Hospitals NHS Foundation Trust (RD7) | Frimley Health NHS Foundation Trust (RDU) | 2015/16 |
| West Middlesex University Hospital NHS Trust (RFW) | Chelsea and Westminster Hospital NHS Foundation Trust (RQM) | 2015/16 |
| Torbay and Southern Devon Health and Care NHS Trust (R1G) | Torbay and South Devon Health Care NHS Foundation Trust (RA9) | 2015/16 |

[i] A mapping from old to new provider codes is available from NHS Digital(16). Note other Trust mergers and splitting occurred during the study timeframe (for example, RG3, RGZ, and RG2 merged to form RYQ in 2010/11), but are excluded from this table because these provider codes were either already updated in our raw data files or not included. Note some provider codes have changed since the close of the HES data in March 2017 (for example, between April 2017 and October 2019 all of the following provider codes had changed due to Trust mergers: RE9, RGQ, RJF, RLN, RNL, RQ6, RQQ, RR1, RW3).

[ii] RYQ split into RJZ, RJ2, and RPG. RJD split into RJE and RL4. Since RJZ and RJE had the most admissions per year(17), these provider codes were assumed.

Table 2: The RECORD Statement: A checklist of items that should be reported in observational studies using routinely collected health data, and the page on which each checklist item appears

| | STROBE items | RECORD items | Page where checklist item appears |
|---|---|---|---|
| **Title and Abstract** | | | |
| | (a) Indicate the study's design with a commonly used term in the title or the abstract. | The type of data used should be specified in the title or abstract. When possible, the name of the databases used should be included. | p.3 |
| | (b) Provide in the abstract an informative and balanced summary of what was done and what was found. | If applicable, the geographic region and time frame within which the study took place should be reported in the title or abstract. | p.3 |
| | | If linkage between databases was conducted for the study, this should be clearly stated in the title or abstract. | N/ A – linkage of HES data with death information from ONS was performed by NHS Digital before data were shared with the study team, as described on p.6. |
| **Introduction** | | | |
| Background and rationale | Explain the scientific background and rationale for the investigation being reported. | | p.5 |
| Objectives | State specific objectives, including any prespecified hypotheses. | | p.6 |
| **Methods** | | | |
| Study Design | Present key elements of study design early in the paper. | | p.6-7 |
| Setting | Describe the setting, locations, and relevant dates, including periods of recruitment, exposure, follow-up, and data collection. | | p.6-7 |
| Participants | (a) Cohort study: Give the eligibility criteria and the sources and methods of selection of participants. Describe methods of follow-up. Case-control study: Give the eligibility criteria | The methods of study population selection (such as codes or algorithms used to identify subjects) should be listed in detail. If this is not possible, an | p.6 and Figure 1 |

| | | | |
|---|---|---|---|
| | and the sources and methods of case ascertainment and control selection. Give the rationale for the choice of cases and controls. Cross-sectional study: Give the eligibility criteria and the sources and methods of selection of participants. | explanation should be provided. RECORD of individuals with linked data at each stage. | |
| | (b) Cohort study: For matched studies, give matching criteria and number of exposed and unexposed. Case-control study: For matched studies, give matching criteria and the number of controls per case. | Any validation studies of the codes or algorithms used to select the population should be referenced. If validation was conducted for this study and not published elsewhere, detailed methods and results should be provided. | N/A |
| | | If the study involved linkage of databases, consider use of a flow diagram or other graphical display to demonstrate the data linkage process, including the number of individuals with linked data at each stage. | Hospital-level antibiotic data was merged with HES data using hospital name, as discussed on p.6-7. Hospital names were updated in advance of the merge to account for hospital mergers/splitting as outlined in Appendix Table 1. |
| Variables | Clearly define all outcomes, exposures, predictors, potential confounders, and effect modifiers. Give diagnostic criteria, if applicable. | A complete list of codes and algorithms used to classify exposures, outcomes, confounders, and effect modifiers should be provided. If these cannot be reported, an explanation should be provided. | Main text: p.7 and p.10, Table 1. Supplementary appendix: p.2-4, Tables 3-5. |
| Data sources / measurement | For each variable of interest, give sources of data and details of methods of assessment (measurement). Describe comparability of assessment methods if there is more than one group. | | Main text: p.6-7, and p.10, Table 1 Supplementary appendix: p.2-4, Tables 1 and 3-5. |
| Bias | Describe any efforts to address potential sources of bias. | | Main text: p.6, p.13 Supplementary Appendix: p.4 |
| Study size | Explain how the study size was arrived at. | | Supplementary Appendix: p.2 and Figure 1 |

| | | | |
|---|---|---|---|
| Quantitative variables | Explain how quantitative variables were handled in the analyses. If applicable, describe which groupings were chosen and why. | | Main text: p.7 and p.10, Table 1<br>Supplementary Appendix: p.2-5 |
| Statistical methods | (a) Describe all statistical methods, including those used to control for confounding. | | Main text: p.7 and p.10<br>Supplementary Appendix: p.3-5 |
| | (b) Describe any methods used to examine subgroups and interactions. | | Main text: p.7 and p.10<br>Supplementary Appendix: p.4 |
| | (c) Explain how missing data were addressed. | | Supplementary Appendix: p.2 |
| | (d) Cohort study: If applicable, explain how loss to follow-up was addressed. Case-control study: If applicable, explain how matching of cases and controls was addressed. Cross-sectional study: If applicable, describe analytical methods taking account of sampling strategy. | | N/A |
| | (e) Describe any sensitivity analyses. | | p.10-12 |
| Data access and cleaning methods | N/A | Authors should describe the extent to which the investigators had access to the database population used to create the study population. | p.6 |
| | | Authors should provide information on the data cleaning methods used in the study. | Main text: p.6-7<br>Supplementary Appendix: p.2-4 and Figure 1 |
| Linkage | N/A | State whether the study included person-level, institutional-level, or other data linkage across two or more databases. The methods of linkage and methods of linkage quality evaluation should be provided. | Hospital-level antibiotic data was merged with HES data using hospital name, as discussed on p.6-7. Hospital names were updated in advance of the merge to account for hospital mergers/splitting as outlined in Appendix Table 1. |
| **Results** | | | |

| | | | |
|---|---|---|---|
| Participants | (a) Report the numbers of individuals at each stage of the study (e.g., numbers potentially eligible, examined for eligibility, confirmed eligible, included in the study, completing follow-up, and analysed). | Describe in detail the selection of the persons included in the study (i.e., study population selection), including filtering based on data quality, data availability, and linkage. The selection of included persons can be described in the text and/or by means of the study flow diagram. | Supplementary Appendix: p.2 and Figure 1 |
| | (b) Give reasons for nonparticipation at each stage. | | Supplementary Appendix: p.2 and Figure 1 |
| | (c) Consider use of a flow diagram. | | Supplementary Appendix: Figure 1 |
| Descriptive data | (a) Give characteristics of study participants (e.g., demographic, clinical, and social) and information on exposures and potential confounders. | | p.7 and 10, Table 1 |
| | (b) Indicate the number of participants with missing data for each variable of interest. | | Supplementary Appendix, p.2 |
| | (c) Cohort study: summarise follow-up time (e.g., average and total amount). | | p.7 |
| Outcome data | Cohort study: Report numbers of outcome events or summary measures over time. Case-control study: Report numbers in each exposure category or summary measures of exposure. Cross-sectional study: Report numbers of outcome events or summary measures. | | Main text: p.11-12, Figure 3-4 |
| Main results | (a) Give unadjusted estimates and, if applicable, confounder-adjusted estimates and their precision (e.g., 95% confidence interval). Make clear which confounders were adjusted for and why they were included. (b) Report category boundaries when continuous variables were categorized. (c) If relevant, consider translating estimates of relative risk into absolute risk for a meaningful time period. | | Table 1 displays factors that were adjusted for in models of mortality and readmission. Figure 3-4 present results of meta-regression models, as do Supplementary Appendix Tables 6-10.<br><br>As outlined in Supplementary Appendix |

| | | | |
|---|---|---|---|
| | | | p.4, marginal effects were derived from models containing 16 admission factors, irrespective of statistical significance, to ensure the best possible adjustment for case-mix, rather than to assess the relative effect of factors within models. Up to 135 hospital-specific models were fit for each outcome analysed. For these reasons, univariate effects are not presented. |
| Other analyses | Report other analyses done—e.g., analyses of subgroups and interactions and sensitivity analyses | | p.10-12, Figure 3 |
| **Discussion** | | | |
| Key results | Summarise key results with reference to study objectives. | | Main text: p.10-12, 14, Figure 3-4<br>Supplementary Appendix: Tables 6-10 |
| Limitations | Discuss limitations of the study, taking into account sources of potential bias or imprecision. Discuss both direction and magnitude of any potential bias. | Discuss the implications of using data that were not created or collected to answer the specific research question(s). Include discussion of misclassification bias, unmeasured confounding, missing data, and changing eligibility over time, as they pertain to the study being reported. | p.13-14 |
| Interpretation | Give a cautious overall interpretation of results considering objectives, limitations, multiplicity of analyses, results from similar studies, and other relevant evidence. | | p.3-4 and 12-14, Figure 3 |
| Generalisability | Discuss the generalisability (external validity) of the study results. | | p.12 and 14 |

| **Other information** | | | |
|---|---|---|---|
| Funding | Give the source of funding and the role of the funders for the present study and, if applicable, for the original study on which the present article is based. | | p.16 |
| Accesibility of protocol, raw data, and programming code | N/A | Authors should provide information on how to access any supplemental information such as the study protocol, raw data, or programming code. | p.15 |

N/A = Not applicable

Table 3: Classification of Antibiotics into Broad-spectrum, Access, Watch, and Reserve Categories, Adapted by Public Health England from the World Health Organization 2017 Essential Medicines List

| Antibiotics | ATC index code 2020 | Classified as broad-spectrum | AWaRe England category |
|---|---|---|---|
| Amikacin | J01GB06 | No | Watch |
| Amoxicillin | J01CA04 | No | Access |
| Amoxicillin/Clavulanic Acid | J01CR02 | Yes | Watch |
| Ampicillin | J01CA01 | No | Access |
| Ampicillin/Flucloxacillin | J01CA51 | No | Access |
| Ampicillin/Flucloxacillin | J01CR50 | No | Access |
| Avibactam/Ceftazidime | J01DD52 | Yes | Reserve |
| Azithromycin | J01FA10 | No | Access or Watch [1] (depending on indication) |
| Aztreonam | J01DF01 | Yes | Reserve |
| Cefaclor[2] | J01DC04 | No | Watch |
| Cefadroxil | J01DB05 | No | Watch |
| Cefalexin | J01DB01 | No | Watch |
| Cefixime | J01DD08 | Yes | Access or Watch (depending on indication) |
| Cefotaxime | J01DD01 | Yes | Access or Watch (depending on indication) |
| Cefpodoxime Proxetil | J01DD13 | Yes | Watch |
| Cefradine | J01DB09 | No | Watch |
| Ceftaroline Fosamil | J01DI02 | Yes | Reserve |
| Ceftazidime | J01DD02 | Yes | Watch |
| Ceftobiprole Medocaril | J01DI01 | Yes | Reserve |
| Ceftolozane/Tazobactam | J01DI54 | Yes | Reserve |
| Ceftriaxone | J01DD04 | Yes | Access or Watch (depending on indication) |
| Cefuroxime | J01DC02 | Yes | Watch |
| Cefuroxime Axetil | J01DC02 | Yes | Watch |
| Chloramphenicol | J01BA01 | No | Watch |
| Cilastatin/Imipenem | J01DH51 | Yes | Reserve |
| Ciprofloxacin | J01MA02 | Yes | Access or Watch (depending on indication) |
| Clarithromycin | J01FA09 | No | Access or Watch (depending on indication) |
| Clindamycin | J01FF01 | No | Watch |
| Colistin | J01XB01 | Yes | Reserve |
| Dalfopristin/Quinupristin | J01FG02 | No | Watch |
| Daptomycin | J01XX09 | No | Reserve |
| Doripenem | J01DH04 | Yes | Reserve |
| Doxycycline | J01AA02 | No | Access |

| Antibiotics | ATC index code 2020 | Classified as broad-spectrum | AWaRe England category |
|---|---|---|---|
| Ertapenem | J01DH03 | Yes | Reserve |
| Erythromycin | J01FA01 | No | Watch |
| Fidaxomicin | A07AA12[3] | No | Watch |
| Flucloxacillin | J01CF05 | No | Access |
| Fosfomycin | J01XX01 | No | Reserve |
| Fusidic Acid | J01XC01 | No | Access |
| Gentamicin | J01GB03 | No | Access |
| Levofloxacin | J01MA12 | Yes | Watch |
| Linezolid | J01XX08 | No | Reserve |
| Lymecycline | J01AA04 | No | Watch |
| Meropenem | J01DH02 | Yes | Reserve |
| Metronidazole | J01XD01 | No | Access |
| Minocycline | J01AA08 | No | Watch |
| Moxifloxacin | J01MA14 | Yes | Watch |
| Nitrofurantoin | J01XE01 | No | Access |
| Norfloxacin | J01MA06 | Yes | Watch |
| Ofloxacin | J01MA01 | Yes | Watch |
| Oxytetracycline | J01AA06 | No | Watch |
| Penicillin G | J01CE01 | No | Access |
| Penicillin V | J01CE02 | No | Access |
| Piperacillin | J01CA12 | Yes | Watch |
| Piperacillin/Tazobactam | J01CR05 | Yes | Access or Watch (depending on indication) |
| Pivmecillinam | J01CA08 | No | Access |
| Sulfamethoxazole/Trimethoprim | J01EE01 | No | Access |
| Tedizolid | J01XX11 | No | Reserve |
| Teicoplanin | J01XA02 | No | Watch |
| Telavancin | J01XA03 | No | Watch |
| Telithromycin | J01FA15 | No | Watch |
| Temocillin | J01CA17 | No | Watch |
| Tetracycline | J01AA07 | No | Access |
| Tigecycline | J01AA12 | Yes | Reserve |
| Tinidazole | J01XD02 | No | Access |
| Tobramycin | J01GB01 | No | Watch |
| Trimethoprim | J01EA01 | No | Access |
| Vancomycin | J01XA01 | No | Access or Watch (depending on indication) |
| Vancomycin | A07AA09[3] | No | Access or Watch (depending on indication) |

[1] Included in meta-regression models as its own category, separate from Access and separate from Watch antibiotics

[2] Although second-generation cephalosporins were classified as broad-spectrum, Cefaclor was not as it is administered orally and is not well absorbed.

[3]Though not absorbed, A07AA12 and A07AA09 are important (alternate) treatment options for *Clostridium difficile* and were retained as they accounted for 0.8% of total DDDs.

Table 4: Number of Acute/General Medical Admissions in 135 Acute Care NHS Hospital Trusts, England, April 2010 to March 2017

| **NHS Acute Care Hospital Trust** | **Admissions (N=36,124,372)** |
|---|---|
| Barts Health NHS Trust | 655,728 (1.82%) |
| Heart of England NHS Foundation Trust | 600,781 (1.66%) |
| University Hospitals of Leicester NHS Trust | 592,378 (1.64%) |
| King's College Hospital NHS Foundation Trust | 577,624 (1.60%) |
| Pennine Acute Hospitals NHS Trust | 563,816 (1.56%) |
| Sheffield Teaching Hospitals NHS Foundation Trust | 563,268 (1.56%) |
| Leeds Teaching Hospitals NHS Trust | 535,954 (1.48%) |
| University Hospital of North Midlands NHS Trust | 515,228 (1.43%) |
| East Kent Hospitals University NHS Foundation Trust | 500,870 (1.39%) |
| The Royal Wolverhampton NHS Trust | 497,593 (1.38%) |
| Shrewsbury and Telford Hospital NHS Trust | 478,043 (1.32%) |
| Nottingham University Hospitals NHS Trust | 475,821 (1.32%) |
| Imperial College Healthcare NHS Trust | 455,868 (1.26%) |
| Royal Devon and Exeter NHS Foundation Trust | 442,324 (1.22%) |
| Norfolk and Norwich University Hospitals NHS Foundation Trust | 439,213 (1.22%) |
| University College London NHS Foundation Trust | 431,899 (1.20%) |
| Oxford University Hospitals NHS Foundation Trust | 431,286 (1.19%) |
| Royal Free London NHS Foundation Trust | 424,790 (1.18%) |
| Frimley Health NHS Foundation Trust | 423,701 (1.17%) |
| London North West Healthcare NHS Trust | 422,383 (1.17%) |
| The Newcastle Upon Tyne Hospitals NHS Foundation Trust | 415,388 (1.15%) |
| South Tees Hospitals NHS Foundation Trust | 397,418 (1.10%) |
| Mid Yorkshire Hospitals NHS Trust | 387,113 (1.07%) |
| York Teaching Hospital NHS Foundation Trust | 374,825 (1.04%) |
| The Dudley Group NHS Foundation Trust | 372,312 (1.03%) |
| Northumbria Healthcare NHS Foundation Trust | 371,996 (1.03%) |
| Barking, Havering and Redbridge University Hospitals NHS Trust | 366,606 (1.01%) |
| East Lancashire Hospitals NHS Trust | 364,950 (1.01%) |
| County Durham and Darlington NHS Foundation Trust | 355,572 (0.98%) |
| Royal Cornwall Hospitals NHS Trust | 355,337 (0.98%) |
| Hull and East Yorkshire Hospitals NHS Trust | 352,449 (0.98%) |
| Central Manchester University Hospitals NHS Foundation Trust | 346,302 (0.96%) |
| United Lincolnshire Hospitals NHS Trust | 344,500 (0.95%) |
| The Royal Bournemouth and Christchurch Hospitals NHS Foundation Trust | 341,723 (0.95%) |
| University Hospital Birmingham NHS Foundation Trust | 337,519 (0.93%) |
| University Hospitals Coventry and Warwickshire NHS Trust | 335,214 (0.93%) |
| Portsmouth Hospitals NHS Trust | 330,267 (0.91%) |
| University Hospital Southampton NHS Foundation Trust | 329,881 (0.91%) |
| Gloucestershire Hospitals NHS Foundation Trust | 326,409 (0.90%) |
| Derby Teaching Hospitals NHS Foundation Trust | 325,309 (0.90%) |

| NHS Acute Care Hospital Trust | Admissions (N=36,124,372) |
|---|---|
| Western Sussex Hospitals NHS Foundation Trust | 323,149 (0.89%) |
| Cambridge University Hospitals NHS Foundation Trust | 317,725 (0.88%) |
| St George's University Hospitals NHS Foundation Trust | 310,309 (0.86%) |
| Aintree University Hospital NHS Foundation Trust | 304,050 (0.84%) |
| Worcestershire Acute Hospitals NHS Trust | 302,599 (0.84%) |
| Royal Liverpool and Broadgreen University Hospitals NHS Trust | 295,000 (0.82%) |
| Doncaster and Bassetlaw Teaching Hospitals NHS Foundation Trust | 293,277 (0.81%) |
| Sandwell and West Birmingham Hospitals NHS Trust | 290,692 (0.80%) |
| Bradford Teaching Hospitals NHS Foundation Trust | 290,347 (0.80%) |
| Guy's and St Thomas' NHS Foundation Trust | 288,611 (0.80%) |
| Wirral University Teaching Hospital NHS Foundation Trust | 279,952 (0.77%) |
| Lancashire Teaching Hospitals NHS Foundation Trust | 276,100 (0.76%) |
| Blackpool Teaching Hospitals NHS Foundation Trust | 272,910 (0.76%) |
| Brighton and Sussex University Hospitals NHS Trust | 272,827 (0.76%) |
| East Sussex Healthcare NHS Trust | 265,799 (0.74%) |
| Hampshire Hospitals NHS Foundation Trust | 262,829 (0.73%) |
| North Tees and Hartlepool NHS Foundation Trust | 262,443 (0.73%) |
| University Hospitals Bristol NHS Foundation Trust | 255,811 (0.71%) |
| Maidstone and Tunbridge Wells NHS Trust | 255,481 (0.71%) |
| Salford Royal NHS Foundation Trust | 255,241 (0.71%) |
| West Hertfordshire Hospitals NHS Trust | 252,202 (0.70%) |
| Chelsea and Westminster Hospital NHS Foundation Trust | 250,648 (0.69%) |
| Royal United Hospitals Bath NHS Foundation Trust | 244,224 (0.68%) |
| North Bristol NHS Trust | 243,843 (0.68%) |
| Basildon and Thurrock University Hospitals NHS Foundation Trust | 243,711 (0.67%) |
| Northern Lincolnshire and Goole NHS Foundation Trust | 243,420 (0.67%) |
| Calderdale and Huddersfield NHS Foundation Trust | 238,032 (0.66%) |
| Plymouth Hospitals NHS Trust | 236,544 (0.65%) |
| St Helens and Knowsley Teaching Hospitals NHS Trust | 235,402 (0.65%) |
| Southend University Hospital NHS Foundation Trust | 233,632 (0.65%) |
| Ipswich Hospital NHS Trust | 229,172 (0.63%) |
| Royal Berkshire NHS Foundation Trust | 226,441 (0.63%) |
| University Hospitals of Morecambe Bay NHS Foundation Trust | 225,491 (0.62%) |
| Colchester Hospital University NHS Foundation Trust | 225,100 (0.62%) |
| City Hospitals Sunderland NHS Foundation Trust | 224,731 (0.62%) |
| University Hospital of South Manchester NHS Foundation Trust | 224,210 (0.62%) |
| Great Western Hospitals NHS Foundation Trust | 216,419 (0.60%) |
| Wrightington, Wigan and Leigh NHS Foundation Trust | 216,327 (0.60%) |
| Epsom and St Helier University Hospitals NHS Trust | 215,461 (0.60%) |
| Sherwood Forest Hospitals NHS Foundation Trust | 213,972 (0.59%) |
| Kettering General Hospital NHS Foundation Trust | 213,856 (0.59%) |
| North Cumbria University Hospitals NHS Trust | 207,188 (0.57%) |

| NHS Acute Care Hospital Trust | Admissions (N=36,124,372) |
|---|---|
| Stockport NHS Foundation Trust | 204,489 (0.57%) |
| The Lewisham and Greenwich NHS Trust | 204,330 (0.57%) |
| Torbay and South Devon Health Care NHS Foundation Trust | 204,304 (0.57%) |
| Taunton and Somerset NHS Foundation Trust | 203,272 (0.56%) |
| Buckinghamshire Healthcare NHS Trust | 199,735 (0.55%) |
| Northampton General Hospital NHS Trust | 198,621 (0.55%) |
| Mid Essex Hospital Services NHS Trust | 196,750 (0.54%) |
| Surrey and Sussex Healthcare NHS Trust | 195,349 (0.54%) |
| Luton and Dunstable University Hospital NHS Foundation Trust | 195,087 (0.54%) |
| Gateshead Health NHS Foundation Trust | 192,158 (0.53%) |
| East and North Hertfordshire NHS Trust | 191,475 (0.53%) |
| Chesterfield Royal Hospital NHS Foundation Trust | 190,801 (0.53%) |
| The Queen Elizabeth Hospital King's Lynn NHS Foundation Trust | 190,063 (0.53%) |
| Bolton NHS Foundation Trust | 188,364 (0.52%) |
| Barnsley Hospital NHS Foundation Trust | 181,844 (0.50%) |
| Poole Hospital NHS Foundation Trust | 179,480 (0.50%) |
| Walsall Healthcare NHS Trust | 178,890 (0.50%) |
| Ashford and St. Peter's Hospitals NHS Foundation Trust | 176,576 (0.49%) |
| Peterborough and Stamford Hospitals NHS Foundation Trust | 173,905 (0.48%) |
| Medway NHS Foundation Trust | 169,650 (0.47%) |
| The Princess Alexandra Hospital NHS Trust | 167,726 (0.46%) |
| The Rotherham NHS Foundation Trust | 165,268 (0.46%) |
| North Middlesex University Hospital NHS Trust | 160,818 (0.45%) |
| Warrington and Halton Hospitals NHS Foundation Trust | 159,990 (0.44%) |
| James Paget University Hospitals NHS Foundation Trust | 157,301 (0.44%) |
| Croydon Health Services NHS Trust | 156,603 (0.43%) |
| Southport and Ormskirk Hospital NHS Trust | 155,726 (0.43%) |
| Milton Keynes Hospital NHS Foundation Trust | 152,350 (0.42%) |
| The Mid Cheshire Hospitals NHS Foundation Trust | 151,949 (0.42%) |
| Tameside Hospital NHS Foundation Trust | 150,714 (0.42%) |
| Dartford and Gravesham NHS Trust | 150,517 (0.42%) |
| Bedford Hospital NHS Trust | 148,377 (0.41%) |
| Kingston Hospital NHS Foundation Trust | 147,291 (0.41%) |
| Burton Hospitals NHS Foundation Trust | 144,039 (0.40%) |
| Airedale NHS Foundation Trust | 142,587 (0.39%) |
| Salisbury NHS Foundation Trust | 139,310 (0.39%) |
| Homerton University Hospital NHS Foundation Trust | 138,945 (0.38%) |
| The Hillingdon Hospitals NHS Foundation Trust | 138,454 (0.38%) |
| Dorset County Hospital NHS Foundation Trust | 131,000 (0.36%) |
| South Warwickshire NHS Foundation Trust | 130,656 (0.36%) |
| West Suffolk NHS Foundation Trust | 129,822 (0.36%) |
| Northern Devon Healthcare NHS Trust | 127,192 (0.35%) |

| NHS Acute Care Hospital Trust | Admissions (N=36,124,372) |
|---|---|
| Countess of Chester Hospital NHS Foundation Trust | 124,431 (0.34%) |
| Royal Surrey County Hospital NHS Foundation Trust | 120,376 (0.33%) |
| South Tyneside NHS Foundation Trust | 120,055 (0.33%) |
| The Whittington Hospital NHS Trust | 117,822 (0.33%) |
| Harrogate and District NHS Foundation Trust | 113,270 (0.31%) |
| Hinchingbrooke Health Care NHS Trust | 103,636 (0.29%) |
| Yeovil District Hospital NHS Foundation Trust | 103,543 (0.29%) |
| George Eliot Hospital NHS Trust | 94,417 (0.26%) |
| East Cheshire NHS Trust | 93,900 (0.26%) |
| Weston Area Health NHS Trust | 89,741 (0.25%) |
| Wye Valley NHS Trust | 86,567 (0.24%) |

Table 5: Clinical Classifications Software (CCS) Diagnosis Group Identified among Acute/General Medical Admissions in 135 Acute Care NHS Hospital Trusts, England, April 2010 to March 2017

| **CCS diagnosis groups collapsed into 29 categories** | **Admissions (N=36,124,372)** |
|---|---|
| Nonspecific chest pain; Cardiac dysrhythmias; Coronary atherosclerosis and other heart disease; Pulmonary heart disease; Essential hypertension, Hypertension with complications and secondary hypertension; Peri-, endo-, and myocarditis; cardiomyopathy (except that caused by tuberculosis or sexually transmitted disease); Conduction disorders; Heart valve disorders; Cardiac arrest and ventricular fibrillation; Other and ill-defined heart disease; Other circulatory disease | 4,649,241 (12.87%) |
| Cancer of bladder; Cancer of bone and connective tissue, Cancer of thyroid, Malignant neoplasm without specification of site; Cancer of brain and nervous system; Cancer of breast; Cancer of bronchus, lung; Cancer of cervix, Cancer of other female genital organs; Cancer of colon; Cancer of head and neck; Cancer of kidney and renal pelvis, Cancer of other urinary organs; Cancer of liver and intrahepatic bile duct; Cancer of oesophagus; Cancer of other GI organs, peritoneum; Cancer of ovary; Cancer of pancreas; Cancer of prostate, Cancer of testis, Cancer of other male genital organs; Cancer of rectum and anus; Cancer of stomach; Cancer of uterus; Cancer, other and unspecified primary, Maintenance chemotherapy; radiotherapy; Cancer, other respiratory and intrathoracic; Hodgkin's disease, Non-Hodgkin's lymphoma; Leukemias; Melanomas of skin, Other non-epithelial cancer of skin; Multiple myeloma; Neoplasms of unspecified nature or uncertain behavior, Nonmalignant breast conditions; Pathological fracture; Secondary malignancies | 3,848,695 (10.65%) |
| Headache; including migraine, Cataract, Retinal detachments; defects; vascular occlusion; and retinopathy, Glaucoma, Blindness and vision defects, Inflammation; infection of eye (except that caused by tuberculosis or sexually transmitted disease), Other eye disorders, Otitis media and related conditions, Conditions associated with dizziness or vertigo, Other ear and sense organ disorders; Syncope; Epilepsy; convulsions; Other nervous system disorders; Malaise and fatigue; Multiple sclerosis, Other hereditary and degenerative nervous system conditions; Parkinson's disease; Paralysis, Late effects of cerebrovascular disease | 2,520,624 (6.98%) |
| Regional enteritis and ulcerative colitis; Diverticulosis & diverticulitis, Anal and rectal conditions; Noninfectious gastroenteritis; Abdominal pain; Abdominal hernia; Intestinal obstruction without hernia; Appendicitis and other appendiceal conditions, Peritonitis and intestinal abscess | 2,288,317 (6.33%) |
| Other gastrointestinal disorders; Gastritis & duodenitis, Other disorders of stomach and duodenum; Gastroduodenal ulcer (except hemorrhage) | 2,062,927 (5.71%) |
| Deficiency and other anemia, Acute posthemorrhagic anemia; Nutritional deficiencies, Disorders of lipid metabolism, Other nutritional; endocrine; and metabolic disorders; Diseases of white blood cells | 1,779,260 (4.93%) |
| Allergic reactions, Rehabilitation care; fitting of prostheses; and adjustment of devices, Administrative/social admission, Medical examination/evaluation, Other aftercare, Other screening for suspected conditions (not mental disorders or infectious disease), Residual codes; unclassified, E Codes: All (external causes of injury and poisoning); Poisoning by psychotropic agents, Poisoning by other medications and drugs, Poisoning by nonmedicinal substances | 1,755,860 (4.86%) |
| Esophageal disorders; Gastrointestinal hemorrhage | 1,546,058 (4.28%) |

| CCS diagnosis groups collapsed into 29 categories | Admissions (N=36,124,372) |
|---|---|
| Other connective tissue disease; Gout and other crystal arthropathies, Rheumatoid arthritis and related disease, Osteoarthritis, Acquired foot deformities, Other acquired deformities, Systemic lupus erythematosus and connective tissue disorders, Other bone disease and musculoskeletal deformities; Other non-traumatic joint disorders | 1,530,254 (4.24%) |
| Pneumonia (except that caused by tuberculosis or sexually transmitted disease); Aspiration pneumonitis; food/vomitus | 1,392,396 (3.85%) |
| Chronic obstructive pulmonary disease and bronchiectasis; Cystic fibrosis, Other lower respiratory disease; Lung disease due to external agents | 1,228,385 (3.40%) |
| Other, outcome risk above overall risk | 1,148,971 (3.18%) |
| Other, outcome risk is below overall risk | 1,058,630 (2.93%) |
| Urinary tract infections; Genitourinary symptoms and ill-defined conditions | 838,968 (2.32%) |
| Acute cerebrovascular disease; Occlusion or stenosis of precerebral arteries, Other and ill-defined cerebrovascular disease, Transient cerebral ischemia; Coma; stupor; and brain damage | 805,434 (2.23%) |
| Acute bronchitis; Asthma; | 756,552 (2.09%) |
| Nephritis; nephrosis; renal sclerosis, Chronic renal failure; Calculus of urinary tract, Other diseases of kidneys and ureters, Other diseases of bladder and urethra | 754,331 (2.09%) |
| Other liver diseases; Biliary tract disease; Liver disease; alcohol-related; Pancreatic disorders (not diabetes) | 646,577 (1.79%) |
| Mental retardation, Senility and organic mental disorders; Alcohol-related mental disorders, Substance-related mental disorders, Affective disorders, Anxiety; somatoform; dissociative; and personality disorders; Other psychoses; Schizophrenia and related disorders, Preadult disorders, Other mental conditions, Personal history of mental disorder | 641,475 (1.78%) |
| Superficial injury; contusion; Open wounds of head; neck; and trunk; Crushing injury or internal injury; Fracture of upper limb; Intracranial injury; Other injuries & conditions due to external causes; Open wounds of extremities; Fracture of lower limb; Fracture of neck of femur (hip); Burns | 637,873 (1.77%) |
| Skin and subcutaneous tissue infections; Other inflammatory condition of skin, Chronic ulcer of skin, Other skin disorders | 559,582 (1.55%) |
| Acute myocardial infarction | 498,736 (1.38%) |
| Spondylosis; intervertebral disc disorders; other back problems, Osteoporosis; Joint disorders and dislocations; trauma-related, Spinal cord injury, Skull and face fractures, Other fractures, Sprains and strains | 481,424 (1.33%) |
| Low risk (probability of outcome <0.5%) | 478,387 (1.32%) |
| Immunity disorders, Sickle cell anemia, Coagulation and hemorrhagic disorders, Other hematologic conditions | 458,254 (1.27%) |
| Influenza, Acute and chronic tonsillitis, Other upper respiratory infections, Other upper respiratory disease, Disorders of teeth and jaw, Diseases of mouth; excluding dental | 456,453 (1.26%) |
| Congestive heart failure; nonhypertensive | 454,455 (1.26%) |
| Intestinal infection | 442,769 (1.23%) |
| Phlebitis; thrombophlebitis and thromboembolism, Varicose veins of lower extremity, Hemorrhoids, Other disease of veins and lymphatics; Lymphadenitis, Gangrene; Peripheral and visceral atherosclerosis; Aortic and peripheral arterial embolism or thrombosis; Aortic; peripheral; and visceral artery aneurysms | 403,484 (1.12%) |

Table 6: Random effects meta-regression of the association between the adjusted probability of death within 30 days of admission (in/out of hospital) and different metrics of hospital-level antibiotic use, among 36,124,372 acute/general medicine admissions to 135 NHS acute care hospital Trusts, April 2010 – March 2017.

| **Metric of antibiotic use**<br>**For each hospital-level increase of:** | **Change in the adjusted probability of 30-day mortality (95% CI)** [1] | **P-value** |
|---|---|---|
| 500 Total DDDs per 1000 bed-days | -0.010% (-0.064,+0.044) | 0.718 |
| 500 Total DDDs per 1000 admissions | -0.021% (-0.044,+0.001) | 0.065 |
| 500 Inpatient DDDs per 1000 bed-days | +0.004% (-0.076,+0.085) | 0.917 |
| 500 Inpatient DDDs per 1000 admissions | -0.014% (-0.045,+0.017) | 0.362 |
| 500 Outpatient DDD per 1000 bed-days | -0.021% (-0.081,+0.038) | 0.483 |
| 500 Outpatient DDD per 1000 admissions | -0.020% (-0.049,+0.008) | 0.161 |
| 500 Oral DDDs per 1000 bed-days | -0.029% (-0.087,+0.028) | 0.316 |
| 500 Oral DDDs per 1000 admissions | **-0.028% (-0.053,-0.003)** | **0.028** |
| 500 Parenteral DDDs per 1000 bed-days | **+0.284% (+0.031,+0.538)** | **0.028** |
| 500 Parenteral DDDs per 1000 admissions | +0.040% (-0.065,+0.145) | 0.454 |
| 500 Narrow-spectrum DDDs per 1000 bed-days | -0.025% (-0.096,+0.046) | 0.483 |
| 500 Narrow-spectrum DDDs per 1000 admissions | -0.026% (-0.055,+0.003) | 0.074 |
| 500 Broad-spectrum DDDs per 1000 bed-days | +0.012% (-0.101,+0.125) | 0.836 |
| 500 Broad-spectrum DDDs per 1000 admissions | -0.019% (-0.072,+0.034) | 0.475 |
| 500 Access DDDs per 1000 bed-days | -0.033% (-0.126,+0.059) | 0.476 |
| 500 Access DDDs per 1000 admissions | -0.032% (-0.072,+0.007) | 0.102 |
| 500 Watch DDDs per 1000 bed-days | +0.035% (-0.083,+0.152) | 0.561 |
| 500 Watch DDDs per 1000 admissions | -0.009% (-0.065,+0.047) | 0.751 |
| 500 Access or Watch (depending on indication) DDDs per 1000 bed-days | -0.091% (-0.272,+0.090) | 0.323 |
| 500 Access or Watch (depending on indication) DDDs per 1000 admissions | -0.078% (-0.157,+0.000) | 0.050 |
| 100 Reserve DDDs per 1000 bed-days | -0.027% (-0.240,+0.186) | 0.800 |
| 100 Reserve DDDs per 100 per 1000 admissions | -0.046% (-0.131,+0.040) | 0.293 |
| 100 Piperacillin/tazobactam and meropenem DDDs per 1000 bed-days | +0.002% (-0.143,+0.147) | 0.978 |
| 100 Piperacillin/tazobactam and meropenem DDDs per 1000 admissions | -0.023% (-0.085,+0.039) | 0.464 |

[1] Probability of death derived from 135 separate (hospital-specific) Poisson models, with each model adjusting for all the factors in Table 1 (main text). Plots for each model are displayed in Appendix Figure 3.

Table 7: Random effects meta-regression of the association between the adjusted probability of death within 30 days of admission (in/out of hospital) and different metrics of hospital-level antibiotic use, among a more narrowly defined cohort of 19,023,144 acute/general medicine admissions, April 2010 – March 2017.

| **Metric of antibiotic use**<br>**For each hospital-level increase of:** | **Change in the adjusted probability of 30-day mortality (95% CI)** [1] | **P-value** |
|---|---|---|
| 500 Total DDDs per 1000 bed-days | +0.006% (-0.058,+0.071) | 0.844 |
| 500 Total DDDs per 1000 admissions | -0.007% (-0.034,+0.020) | 0.626 |
| 500 Inpatient DDDs per 1000 bed-days | +0.042% (-0.051,+0.136) | 0.374 |
| 500 Inpatient DDDs per 1000 admissions | +0.005% (-0.031,+0.041) | 0.779 |
| 500 Outpatient DDD per 1000 bed-days | -0.021% (-0.092,+0.050) | 0.557 |
| 500 Outpatient DDD per 1000 admissions | -0.016% (-0.049,+0.017) | 0.343 |
| 500 Oral DDDs per 1000 bed-days | -0.017% (-0.087,+0.053) | 0.630 |
| 500 Oral DDDs per 1000 admissions | -0.016% (-0.046,+0.014) | 0.299 |
| 500 Parenteral DDDs per 1000 bed-days | **+0.373% (+0.082,+0.663)** | **0.012** |
| 500 Parenteral DDDs per 1000 admissions | +0.113% (-0.008,+0.235) | 0.068 |
| 500 Narrow-spectrum DDDs per 1000 bed-days | -0.013% (-0.097,+0.071) | 0.761 |
| 500 Narrow-spectrum DDDs per 1000 admissions | -0.014% (-0.048,+0.020) | 0.431 |
| 500 Broad-spectrum DDDs per 1000 bed-days | +0.066% (-0.069,+0.201) | 0.337 |
| 500 Broad-spectrum DDDs per 1000 admissions | +0.017% (-0.046,+0.080) | 0.592 |
| 500 Access DDDs per 1000 bed-days | -0.027% (-0.137,+0.082) | 0.624 |
| 500 Access DDDs per 1000 admissions | -0.019% (-0.065,+0.027) | 0.407 |
| 500 Watch DDDs per 1000 bed-days | +0.078% (-0.063,+0.218) | 0.276 |
| 500 Watch DDDs per 1000 admissions | +0.022% (-0.044,+0.089) | 0.508 |
| 500 Access or Watch (depending on indication) DDDs per 1000 bed-days | -0.042% (-0.258,+0.173) | 0.698 |
| 500 Access or Watch (depending on indication) DDDs per 1000 admissions | -0.038% (-0.133,+0.056) | 0.426 |
| 100 Reserve DDDs per 1000 bed-days | +0.206% (-0.037,+0.450) | 0.096 |
| 100 Reserve DDDs per 100 per 1000 admissions | +0.055% (-0.043,+0.154) | 0.270 |
| 100 Piperacillin/tazobactam and meropenem DDDs per 1000 bed-days | +0.096% (-0.072,+0.265) | 0.260 |
| 100 Piperacillin/tazobactam and meropenem DDDs per 1000 admissions | +0.026% (-0.047,+0.099) | 0.486 |

[1] To improve model stability, five hospital Trusts with fewer than 50,000 admissions were excluded, including City Hospitals Sunderland NHS Foundation Trust, Harrogate and District NHS Foundation Trust, Royal Surrey County Hospital NHS Foundation Trust, The Hillingdon Hospitals NHS Foundation Trust, and The Whittington Hospital NHS Trust. Thus, the probability of death was derived from 130 separate (hospital-specific) Poisson models, with each model adjusting for all the factors in Table 1 (main text) excluding admission specialty. Two out of 130 models required the removal of an interaction term between admission method and admission year in order for the model to converge.

Table 8: Random effects meta-regression of the association between the adjusted probability of death within 30 days of admission (in/out of hospital) and different metrics of hospital-level antibiotic use, among 16,492,990 acute/general medicine admissions, 01/April/2014 – 31/March/2017.

| **Metric of antibiotic use**<br>**For each hospital-level increase of:** | **Change in the adjusted probability of re-admissions (95% CI)** [1] | **P-value** |
|---|---|---|
| 500 Total DDDs per 1000 bed-days | +0.013 (-0.053,+0.080) | 0.691 |
| 500 Total DDDs per 1000 admissions | +0.005 (-0.023,+0.034) | 0.713 |
| 500 Inpatient DDDs per 1000 bed-days | +0.061 (-0.039,+0.160) | 0.231 |
| 500 Inpatient DDDs per 1000 admissions | +0.021 (-0.017,+0.060) | 0.281 |
| 500 Outpatient DDD per 1000 bed-days | -0.020 (-0.093,+0.054) | 0.592 |
| 500 Outpatient DDD per 1000 admissions | -0.009 (-0.044,+0.026) | 0.612 |
| 500 Oral DDDs per 1000 bed-days | +0.029 (-0.090,+0.148) | 0.634 |
| 500 Oral DDDs per 1000 admissions | +0.003 (-0.028,+0.035) | 0.848 |
| 500 Parenteral DDDs per 1000 bed-days | +0.160 (-0.161,+0.481) | 0.326 |
| 500 Parenteral DDDs per 1000 admissions | +0.075 (-0.057,+0.207) | 0.261 |
| 500 Narrow-spectrum DDDs per 1000 bed-days | -0.001 (-0.090,+0.088) | 0.979 |
| 500 Narrow-spectrum DDDs per 1000 admissions | +0.002 (-0.034,+0.039) | 0.908 |
| 500 Broad-spectrum DDDs per 1000 bed-days | +0.063 (-0.077,+0.204) | 0.373 |
| 500 Broad-spectrum DDDs per 1000 admissions | +0.031 (-0.034,+0.096) | 0.353 |
| 500 Access DDDs per 1000 bed-days | -0.025 (-0.142,+0.093) | 0.678 |
| 500 Access DDDs per 1000 admissions | -0.004 (-0.053,+0.046) | 0.886 |
| 500 Watch DDDs per 1000 bed-days | +0.092 (-0.054,+0.238) | 0.214 |
| 500 Watch DDDs per 1000 admissions | +0.043 (-.026,+0.112) | 0.215 |
| 500 Access or Watch (depending on indication) DDDs per 1000 bed-days | +0.026 (-0.198,+0.250) | 0.818 |
| 500 Access or Watch (depending on indication) DDDs per 1000 admissions | +0.012 (-0.087,+0.111) | 0.811 |
| 100 Reserve DDDs per 1000 bed-days | -0.033 (-0.303,+0.238) | 0.812 |
| 100 Reserve DDDs per 100 per 1000 admissions | -0.023 (-0.131,+0.084) | 0.666 |
| 100 Piperacillin/tazobactam and meropenem DDDs per 1000 bed-days | +0.001 (-0.180,+0.181) | 0.994 |
| 100 Piperacillin/tazobactam and meropenem DDDs per 1000 admissions | +0.002 (-0.075,+0.080) | 0.952 |

[1] To improve model stability, four hospital Trusts with fewer than 50,000 admissions were excluded, including East Cheshire NHS Trust, George Eliot Hospital NHS Trust, Weston Area Health NHS Trust, and Wye Valley NHS Trust. Royal Surrey County Hospital was also excluded as it had just nine admissions in the reference group of admission specialty. City Hospitals Sunderland was excluded as it had no remaining admissions in the reference group of admission specialty. The probability of death was therefore derived from 129 separate (hospital-specific) Poisson models, with each model adjusting for all the factors in Table 1 (main text). Charlson comorbidity index and IMD score were modelled as linear predictors, rather than using natural cubic splines, as the use of splines led to poorly fitting models or the failure of models to converge.

Table 9: Random effects meta-regression of the association between the adjusted probability of death within 14 days of admission (in/out of hospital) and different metrics of hospital-level antibiotic use, among 36,124,372 acute/general medicine admissions, 01/April/2010 – 31/March/2017.

| **Metric of antibiotic use**<br>**For each hospital-level increase of:** | **Change in the adjusted probability of re-admissions (95% CI)** [1] | **P-value** |
|---|---|---|
| 500 Total DDDs per 1000 bed-days | -0.009% (-0.054,+0.037) | 0.699 |
| 500 Total DDDs per 1000 admissions | **-0.019% (-0.038,-0.000)** | **0.047** |
| 500 Inpatient DDDs per 1000 bed-days | +0.003% (-0.065,+0.071) | 0.928 |
| 500 Inpatient DDDs per 1000 admissions | -0.014% (-0.040,+0.012) | 0.290 |
| 500 Outpatient DDD per 1000 bed-days | -0.015% (-0.065,+0.036) | 0.564 |
| 500 Outpatient DDD per 1000 admissions | -0.016% (-0.040,+0.008) | 0.186 |
| 500 Oral DDDs per 1000 bed-days | -0.027% (-0.076,+0.022) | 0.273 |
| 500 Oral DDDs per 1000 admissions | **-0.026% (-0.046,-0.005)** | **0.016** |
| 500 Parenteral DDDs per 1000 bed-days | **+0.267% (+0.054,+0.479)** | **0.014** |
| 500 Parenteral DDDs per 1000 admissions | +0.039% (-0.050,+0.128) | 0.384 |
| 500 Narrow-spectrum DDDs per 1000 bed-days | -0.025% (-0.085,+0.035) | 0.412 |
| 500 Narrow-spectrum DDDs per 1000 admissions | **-0.026% (-0.050,-0.002)** | **0.036** |
| 500 Broad-spectrum DDDs per 1000 bed-days | +0.023% (-0.072,+0.119) | 0.630 |
| 500 Broad-spectrum DDDs per 1000 admissions | -0.011% (-0.055,+0.034) | 0.639 |
| 500 Access DDDs per 1000 bed-days | -0.034% (-0.112,+0.044) | 0.392 |
| 500 Access DDDs per 1000 admissions | -0.032% (-0.065,+0.000) | 0.052 |
| 500 Watch DDDs per 1000 bed-days | +0.036% (-0.063,+0.135) | 0.475 |
| 500 Watch DDDs per 1000 admissions | -0.007% (-0.054,+0.041) | 0.784 |
| 500 Access or Watch (depending on indication) DDDs per 1000 bed-days | -0.069% (-0.222,+0.083) | 0.370 |
| 500 Access or Watch (depending on indication) DDDs per 1000 admissions | -0.064% (-0.130,+0.002) | 0.056 |
| 100 Reserve DDDs per 1000 bed-days | +0.029% (-0.150,+0.208) | 0.750 |
| 100 Reserve DDDs per 100 per 1000 admissions | -0.021% (-0.093,+0.051) | 0.563 |
| 100 Piperacillin/tazobactam and meropenem DDDs per 1000 bed-days | -0.003% (-0.125,+0.120) | 0.966 |
| 100 Piperacillin/tazobactam and meropenem DDDs per 1000 admissions | -0.022% (-0.075,+0.030) | 0.399 |

[1] Each model adjusting for all the factors in Table 1 (main text). However, 14 models would only converge with Charlson comorbidity index and IMD score modelled as linear predictors, rather than as natural cubic splines.

Table 10: Random effects meta-regression of the association between the adjusted probability of non-elective re-admission within 30 days of discharge and different metrics of hospital-level antibiotic use, among 34,427,698 acute/general medicine admissions discharged alive, 01/April/2010 – 28/February/2017.

| **Metric of antibiotic use**<br>**For each hospital-level increase of:** | **Change in the adjusted probability of re-admissions (95% CI)** [1] | **P-value** |
|---|---|---|
| 500 Total DDDs per 1000 bed-days | **+0.404 (+0.157, +0.652)** | **0.002** |
| 500 Total DDDs per 1000 admissions | **+0.208 (+0.106,+0.311)** | **<0.001** |
| 500 Inpatient DDDs per 1000 bed-days | +0.149 (-0.234,+0.531) | 0.443 |
| 500 Inpatient DDDs per 1000 admissions | +0.117 (-0.028,+0.261) | 0.114 |
| 500 Outpatient DDD per 1000 bed-days | **+0.380 (+0.103,+0.657)** | **0.008** |
| 500 Outpatient DDD per 1000 admissions | **+0.213 (+0.082,+0.344)** | **0.002** |
| 500 Oral DDDs per 1000 bed-days | **+0.448 (+0.183,+0.713)** | **0.001** |
| 500 Oral DDDs per 1000 admissions | **+0.233 (+0.120,+0.345)** | **<0.001** |
| 500 Parenteral DDDs per 1000 bed-days | +0.431 (-0.793,+1.655) | 0.488 |
| 500 Parenteral DDDs per 1000 admissions | +0.463 (-0.031,+0.956) | 0.066 |
| 500 Narrow-spectrum DDDs per 1000 bed-days | **+0.521 (+0.194,+0.848)** | **0.002** |
| 500 Narrow-spectrum DDDs per 1000 admissions | **+0.254 (+0.124,+0.384)** | **<0.001** |
| 500 Broad-spectrum DDDs per 1000 bed-days | +0.514 (-0.018,+1.046) | 0.058 |
| 500 Broad-spectrum DDDs per 1000 admissions | **+0.330 (+0.085,+0.575)** | **0.009** |
| 500 Access DDDs per 1000 bed-days | **+0.592 (+0.164,+1.021)** | **0.007** |
| 500 Access DDDs per 1000 admissions | **+0.315 (+0.137,+0.493)** | **0.001** |
| 500 Watch DDDs per 1000 bed-days | **+0.719 (+0.173,+1.266)** | **0.010** |
| 500 Watch DDDs per 1000 admissions | **+0.437 (+0.181,+0.692)** | **0.001** |
| 500 Access or Watch (depending on indication) DDDs per 1000 bed-days | +0.697 (-0.161,+1.554) | 0.110 |
| 500 Access or Watch (depending on indication) DDDs per 1000 admissions | **+0.436 (+0.067,+0.806)** | **0.021** |
| 100 Reserve DDDs per 1000 bed-days | -0.162 (-1.175,+0.851) | 0.753 |
| 100 Reserve DDDs per 100 per 1000 admissions | +0.028 (-0.385,+0.440) | 0.894 |
| 100 Piperacillin/tazobactam and meropenem DDDs per 1000 bed-days | -0.109 (-0.800,+0.581) | 0.755 |
| 100 Piperacillin/tazobactam and meropenem DDDs per 1000 admissions | +0.073 (-0.222,+0.368) | 0.624 |

[1] Each model adjusted for all the factors in Table 1 (main text). Charlson comorbidity score was modelled as a linear predictors, rather than using natural cubic splines, as the use of splines led to poorly fitting models or the failure of models to converge.

**APPENDIX FIGURES**

Figure 1: Order of Data Cleaning Steps Followed to Derive Analytic Cohort

| | |
|---|---|
| n = 88,718,419 spells | Dropped 24,560 spells with unfinished consultant episodes[i] |
| n = 88,693,859 spells | Dropped 274 spells with missing discharge date |
| n = 88,693,585 spells | Dropped 1,577,400 spells from patients <16 years of age in all spells |
| n = 87,116,185 spells | Dropped 367,918 spells from patients missing age in all spells |
| n = 86,748,267 spells | Dropped 14,496 spells from patients with ≥10 ages recorded across all spells[ii] |
| n =86,733,771 spells | Dropped 25,866 spells from patients with >1 death date[iii] |
| n = 86,707,905 spells | Dropped 3,444,789 spells from patients not admitted to general medicine (ever)[iv] |
| n = 83,263,116 spells[v] | Dropped 40,825,371 spells not in general medicine[iv] |
| n = 42,437,745 spells | Dropped 56,229 spells <16 years of age at admission[vi] |
| n = 42,381,516 spells | Dropped 4,540,135 admissions before 01 April 2010[vii] |
| n = 37,841,381 spells | Dropped 510,509 spells not in acute care hospital Trusts |
| n = 37,330,872 spells | Dropped 485,843 spells from hospital Trusts with <50,000 spells during study |
| n = 36,845,029 spells | Dropped 266,969 spells missing IMD score |
| n = 36,578,060 spells | Dropped 453,688 spells in specialist hospital Trusts[viii] |
| n = 36,124,372 spells | |

[i] These spells are duplicated across adjacent HES data-years and thus the duplicate was dropped(1)

[ii] Raw data contained admissions between 2009-2017 (also see [vii] below). Patients with ≥10 ages recorded were dropped, as age was judged to be invalid.

[iii] If a patient was recorded to have died in >1 spell, the earliest date of death was retained and subsequent spells were dropped

[iv] Acute/general medicine defined as in Figure 1 (main text)

[v] At this point, previous hospital exposure and re-admission after discharge were estimated, as all patients had at least 1 admission that met our inclusion criteria in Figure 1 (main text)

[vi] Among patients ever admitted ≥16 years of age

[vii] Admissions between 1 April 2009 to 31 March 2010 were used only to estimate previous hospital exposure for spells beginning from 01 April 2010

[viii] Includes one hospital Trust where most (84.9%) admissions were to clinical haematology (The Royal Marsden NHS Foundation Trust) and three hospital Trusts where all admissions were to either cardiology or respiratory medicine (Royal Brompton and Harefield NHS Foundation Trust, Papworth Hospital NHS Foundation Trust, and Liverpool Heart and Chest NHS Foundation Trust) – i.e. no admissions had a HES main specialty code (mainspef) or treatment specialty code (tretspef) equal to 300 in the first or second consultant episode

Figure 2: Non-linear Relative Risks of Death Within 30 Days of Admission (In/Out of Hospital) Among Model Covariates Fit As Natural Cubic Splines

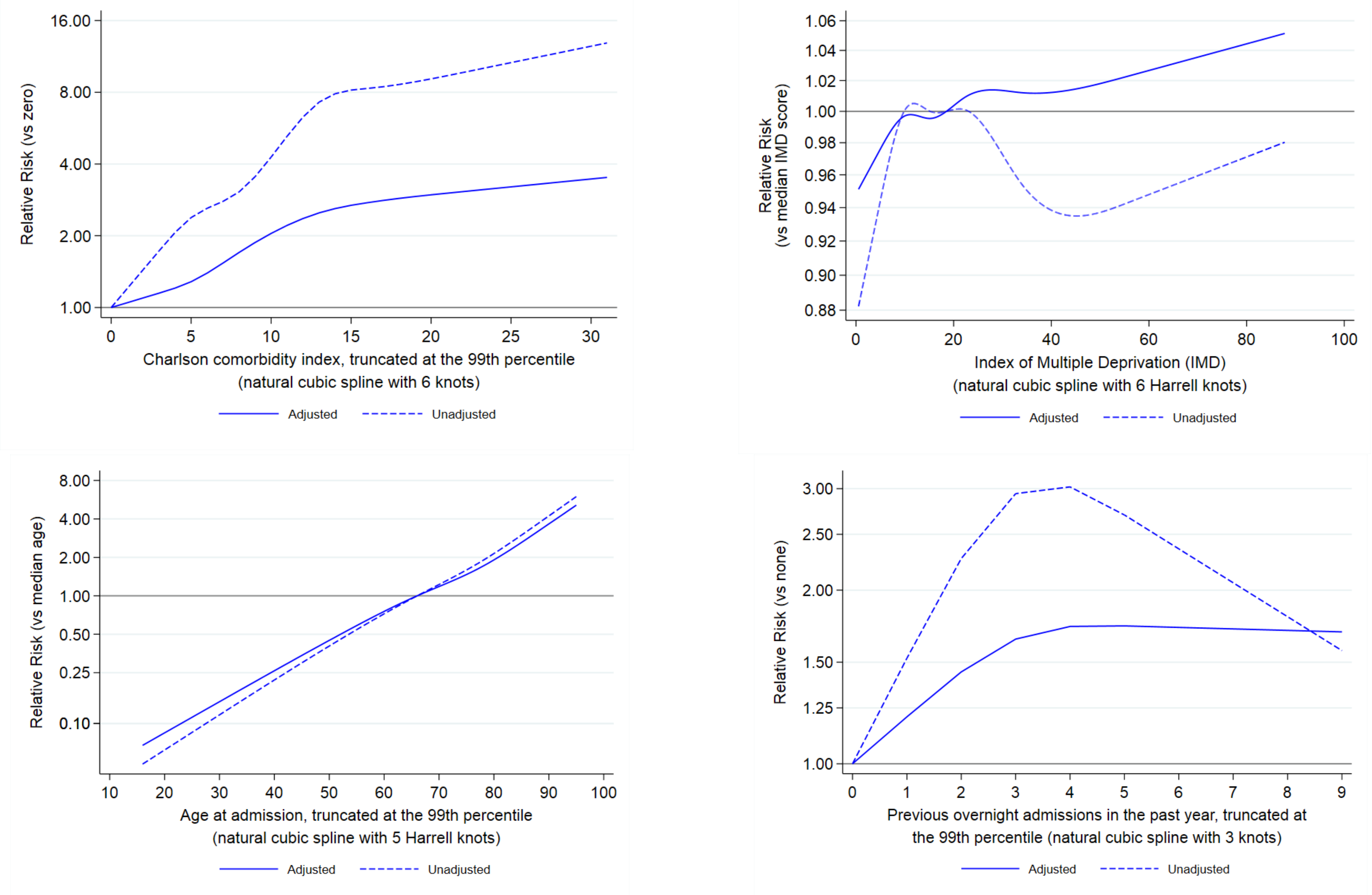

Adjusted risk estimates derived from a single Poisson model containing NHS hospital Trust as a factor and all admission characteristics in Table 1 (main text).

Figure 3: Top 10 Most Commonly Used Antibiotics as a Fraction of Total DDDs Within Each of 135 NHS Acute Care Hospital Trusts (2016), Ranked by Amoxicillin/Clavulanic Acid

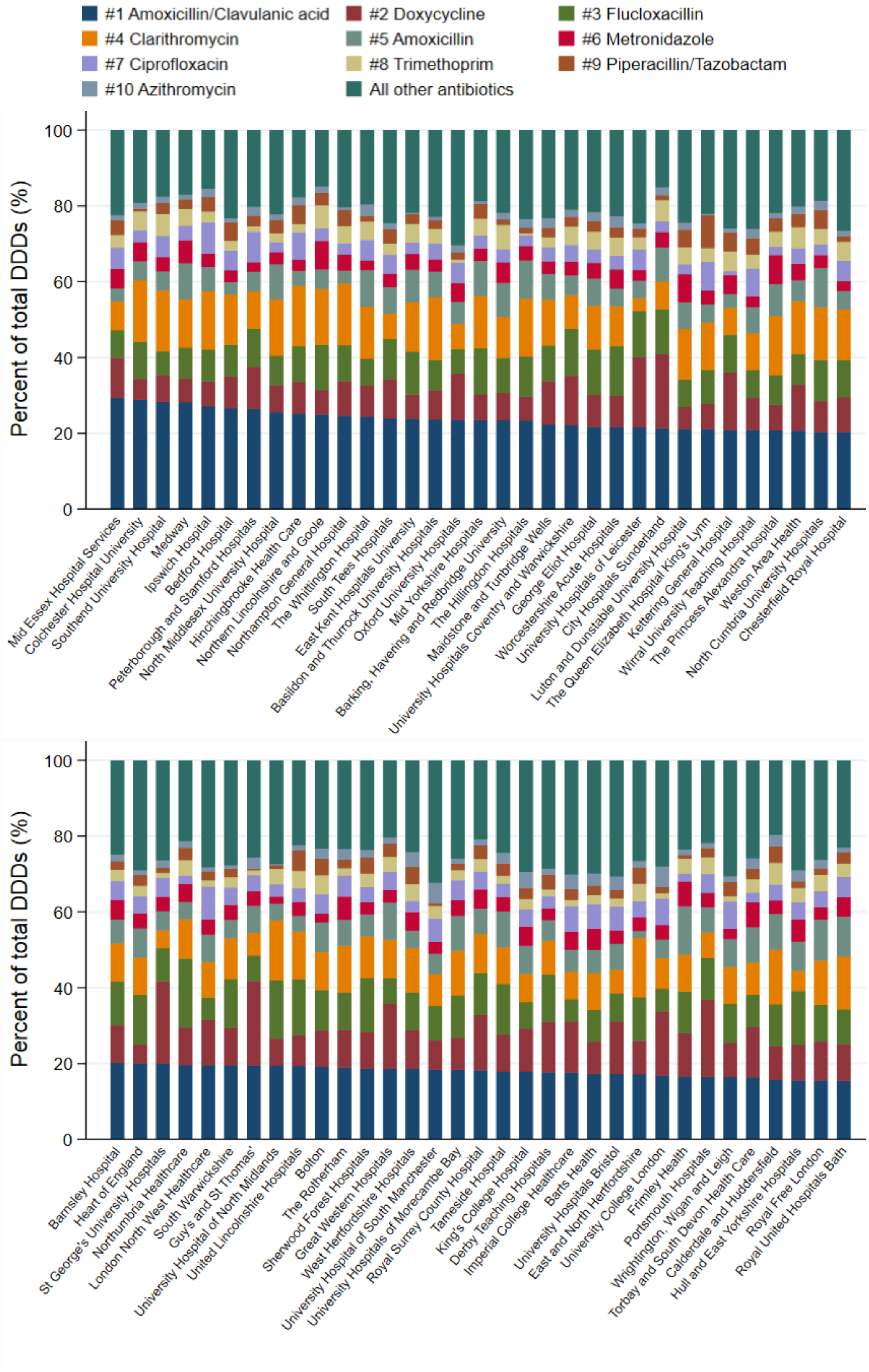

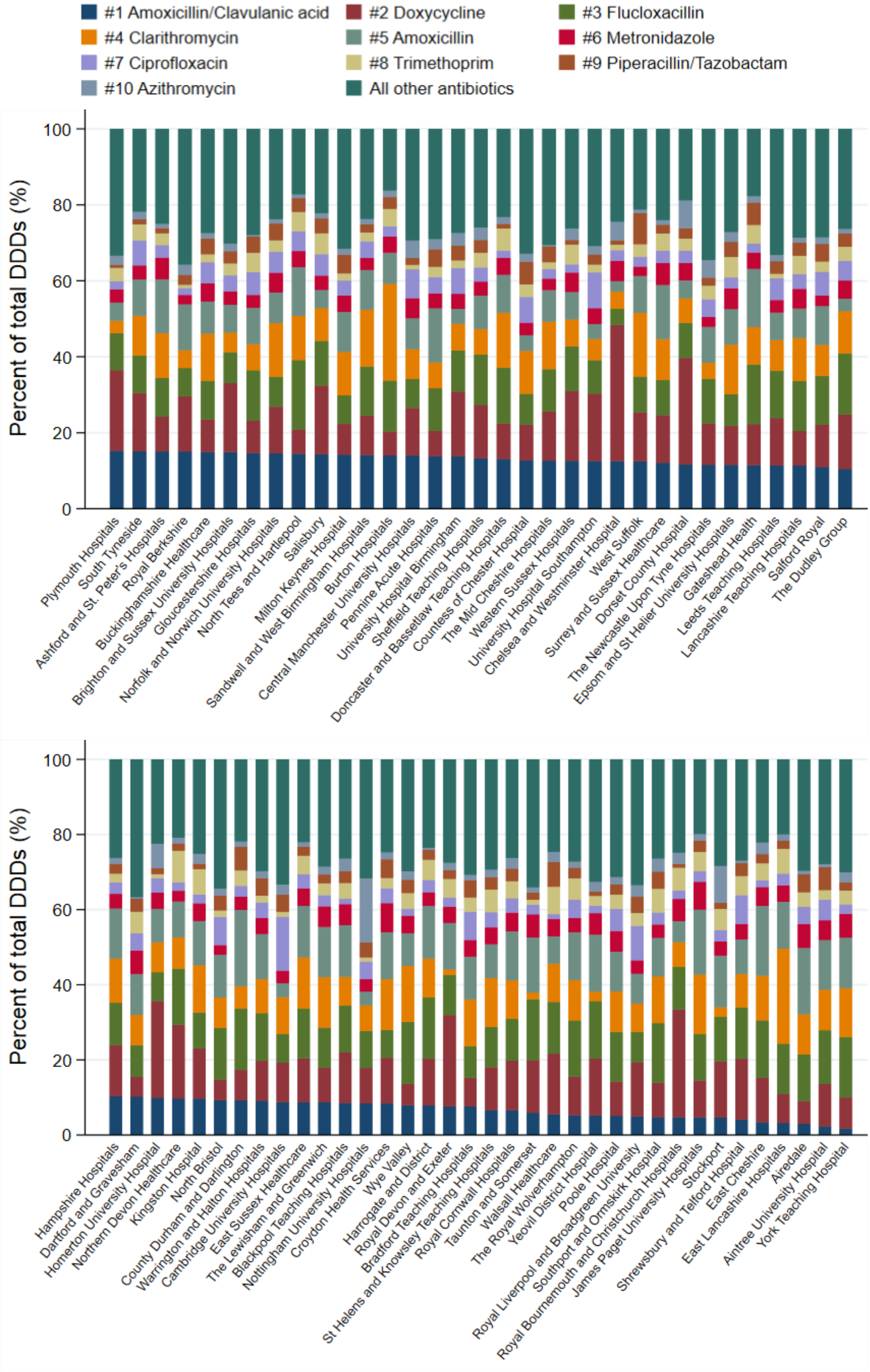
#1 Amoxicillin/Clavulanic acid
#2 Doxycycline
#3 Flucloxacillin
#4 Clarithromycin
#5 Amoxicillin
#6 Metronidazole
#7 Ciprofloxacin
#8 Trimethoprim
#9 Piperacillin/Tazobactam
#10 Azithromycin
All other antibiotics
Percent of total DDDs (%)
0
20
40
60
80
100
Plymouth Hospitals
South Tyneside
Ashford and St. Peter's Hospitals
Royal Berkshire
Buckinghamshire Healthcare
Brighton and Sussex University Hospitals
Gloucestershire Hospitals
Norfolk and Norwich University Hospitals
North Tees and Hartlepool
Salisbury
Milton Keynes Hospital
Sandwell and West Birmingham Hospitals
Burton Hospitals
Central Manchester University Hospitals
Pennine Acute Hospitals
University Hospital Birmingham
Sheffield Teaching Hospitals
Doncaster and Bassetlaw Teaching Hospitals
Countess of Chester Hospital
The Mid Cheshire Hospitals
Western Sussex Hospitals
University Hospital Southampton
Chelsea and Westminster Hospital
West Suffolk
Surrey and Sussex Healthcare
Dorset County Hospital
The Newcastle Upon Tyne Hospitals
Epsom and St Helier University Hospitals
Gateshead Health
Leeds Teaching Hospitals
Lancashire Teaching Hospitals
Salford Royal
The Dudley Group
Percent of total DDDs (%)
0
20
40
60
80
100
Hampshire Hospitals
Dartford and Gravesham
Homerton University Hospital
Northern Devon Healthcare
Kingston Hospital
North Bristol
County Durham and Darlington
Warrington and Halton Hospitals
Cambridge University Hospitals
East Sussex Healthcare
The Lewisham and Greenwich
Blackpool Teaching Hospitals
Nottingham University Hospitals
Croydon Health Services
Wye Valley
Harrogate and District
Royal Devon and Exeter
Bradford Teaching Hospitals
St Helens and Knowsley Teaching Hospitals
Royal Cornwall Hospitals
Taunton and Somerset
Walsall Healthcare
The Royal Wolverhampton
Yeovil District Hospital
Poole Hospital
Royal Liverpool and Broadgreen University
Southport and Ormskirk Hospital
Royal Bournemouth and Christchurch Hospitals
James Paget University Hospitals
Stockport
Shrewsbury and Telford Hospital
East Cheshire
East Lancashire Hospitals
Airedale
Aintree University Hospital
York Teaching Hospital

Figure 4: Random Effects Meta-Regression of the Association Between Adjusted Probability of Death Within 30 Days of Admission (In/Out of Hospital) and Hospital-level Antibiotic Use

4.1a) Total DDDs/1,000 bed-days*

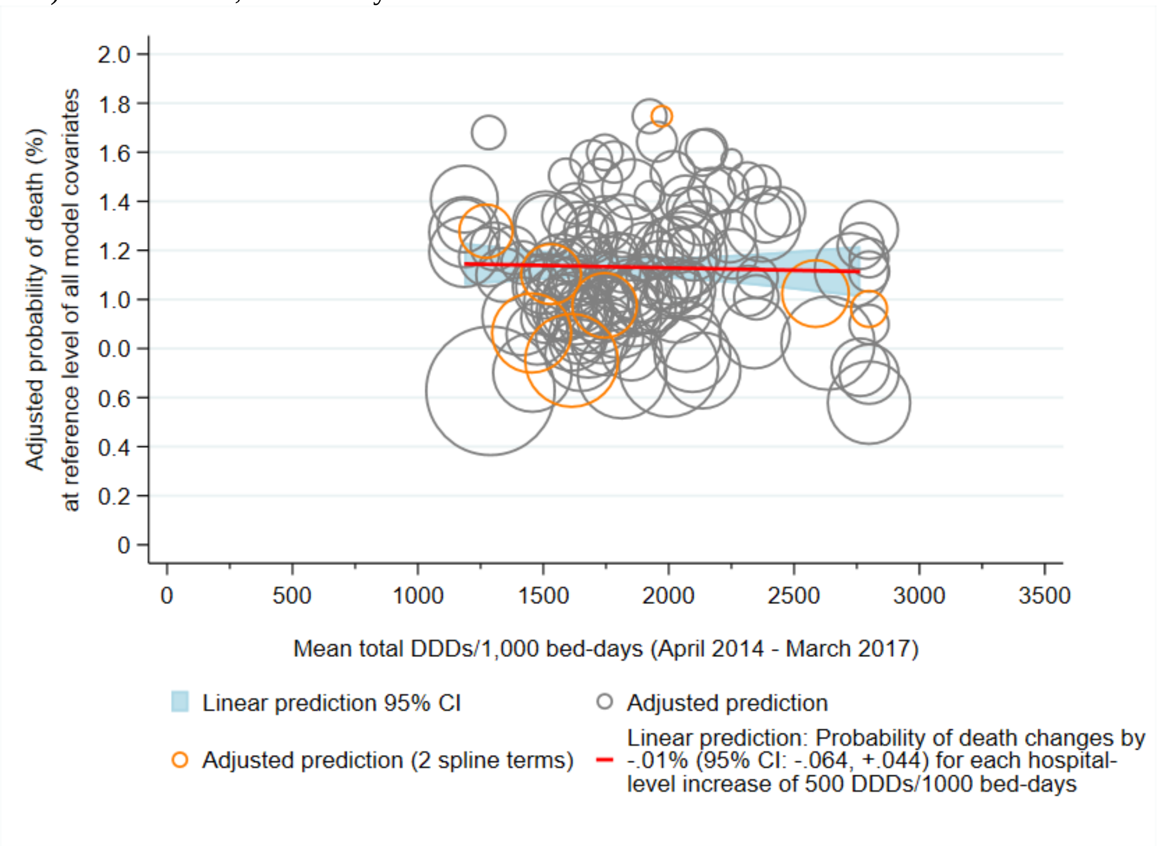

4.1b) Total DDDs/1,000 admissions*

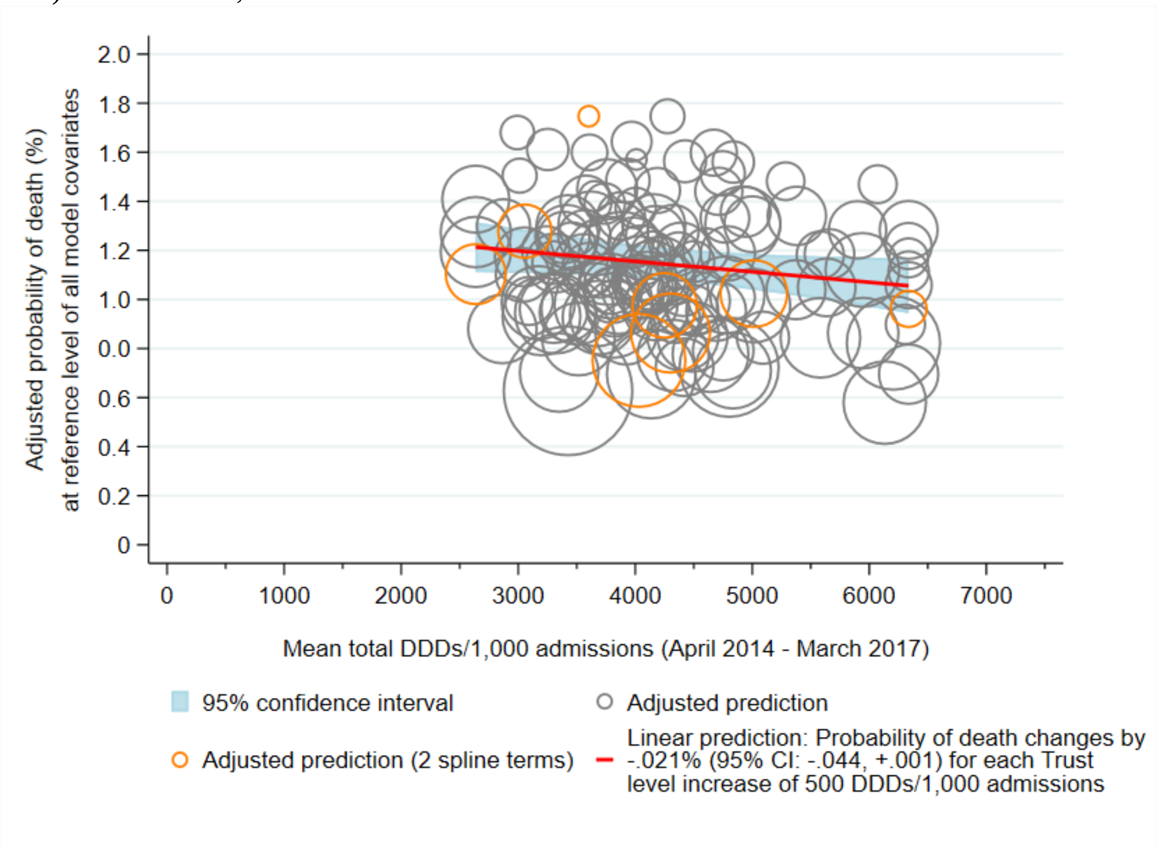

4.2a) Inpatient DDDs/1,000 bed-days*

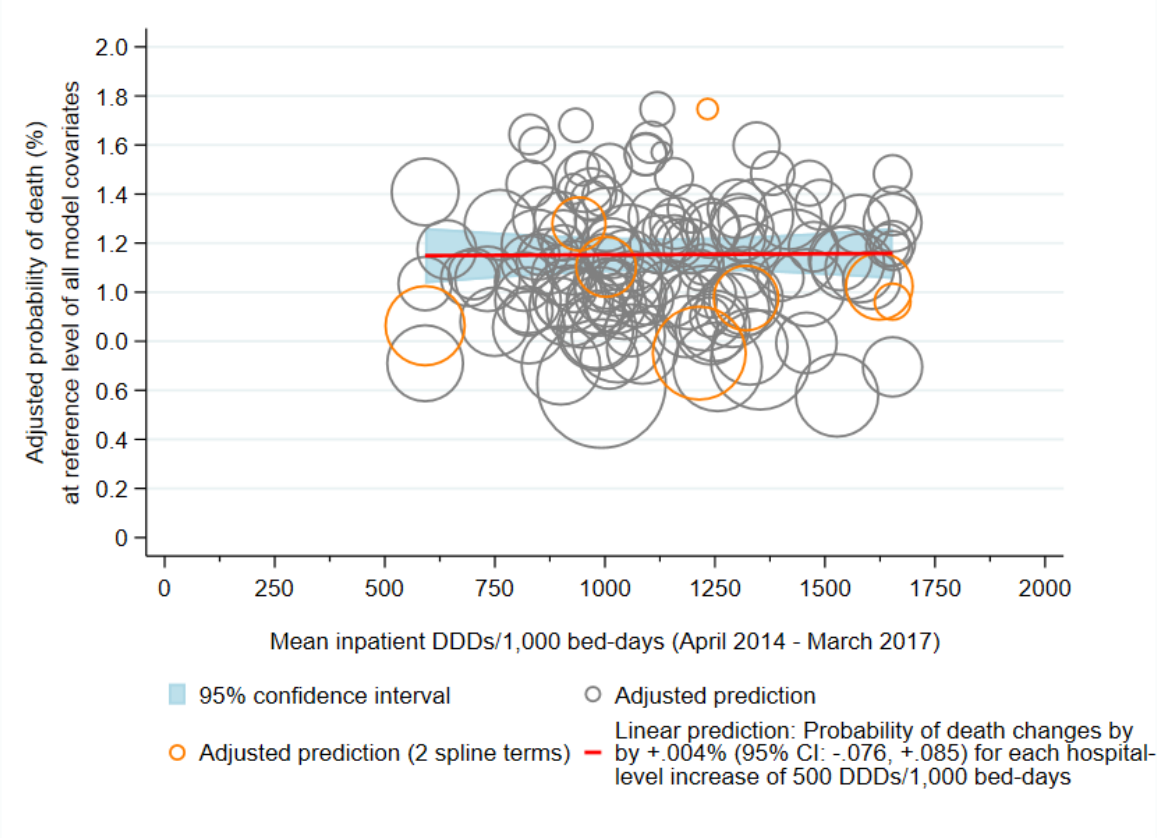

4.2b) Inpatient DDDs/1,000 admissions*

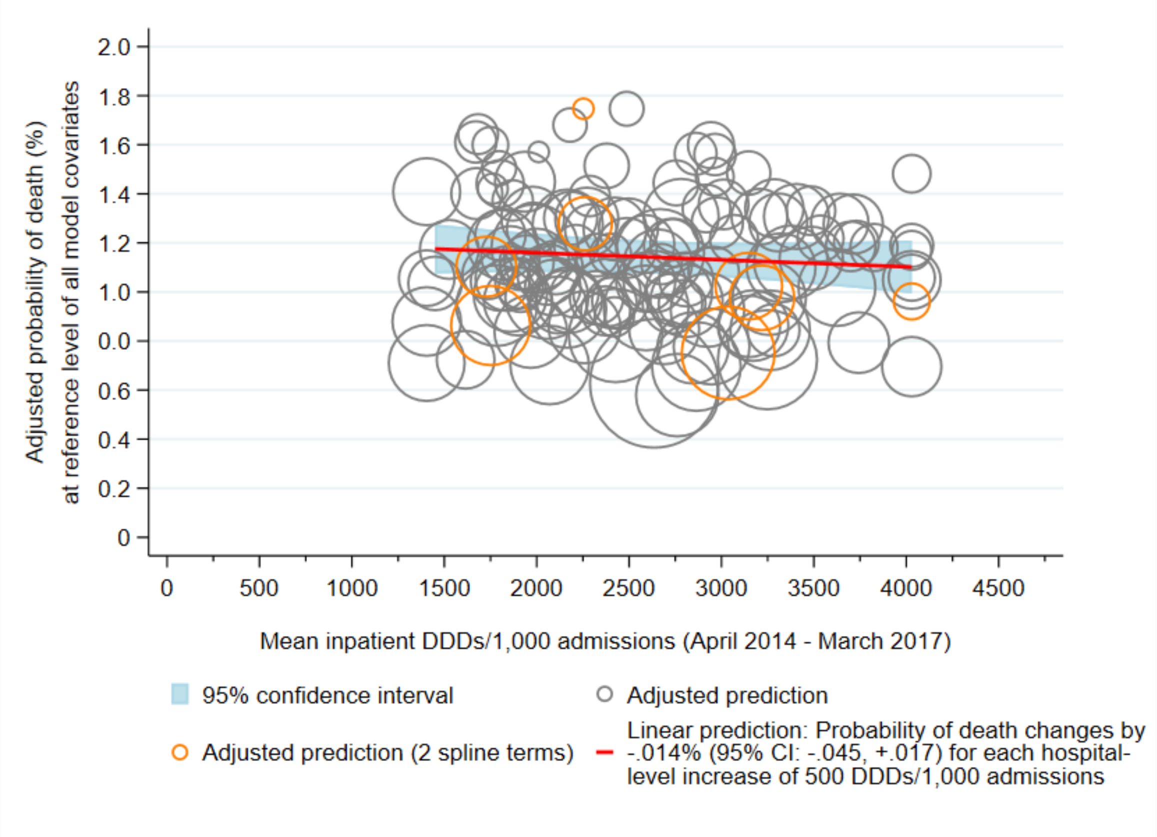

4.3a) Outpatient DDDs/1,000 bed-days*

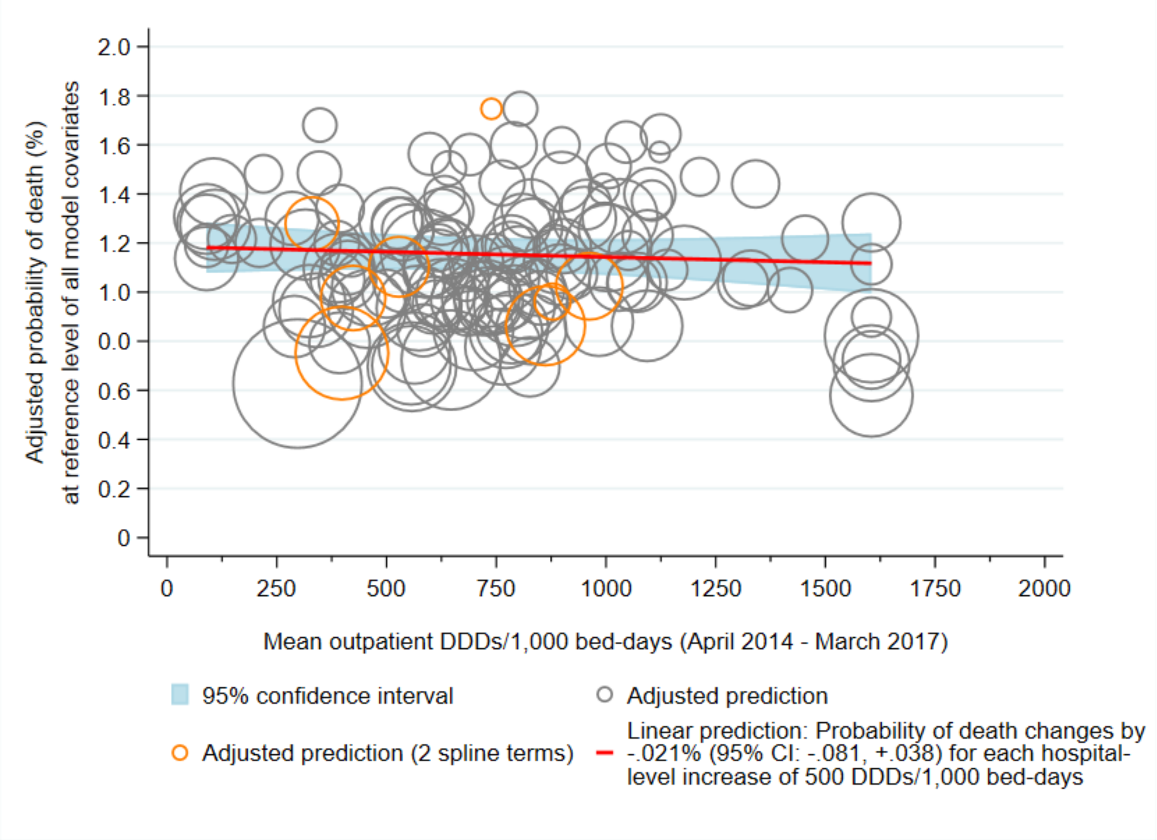

4.3b) Outpatient DDDs/1,000 admissions*

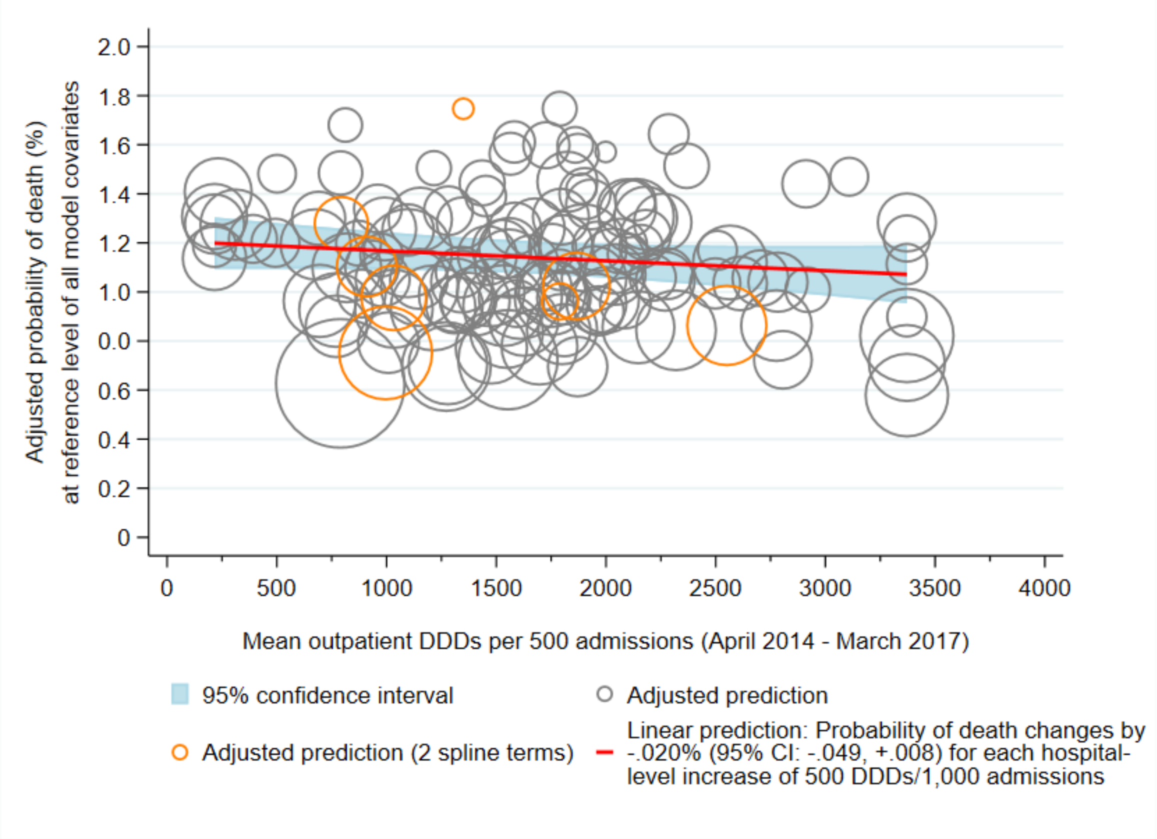

4.4a) Oral DDDs/1,000 bed-days*

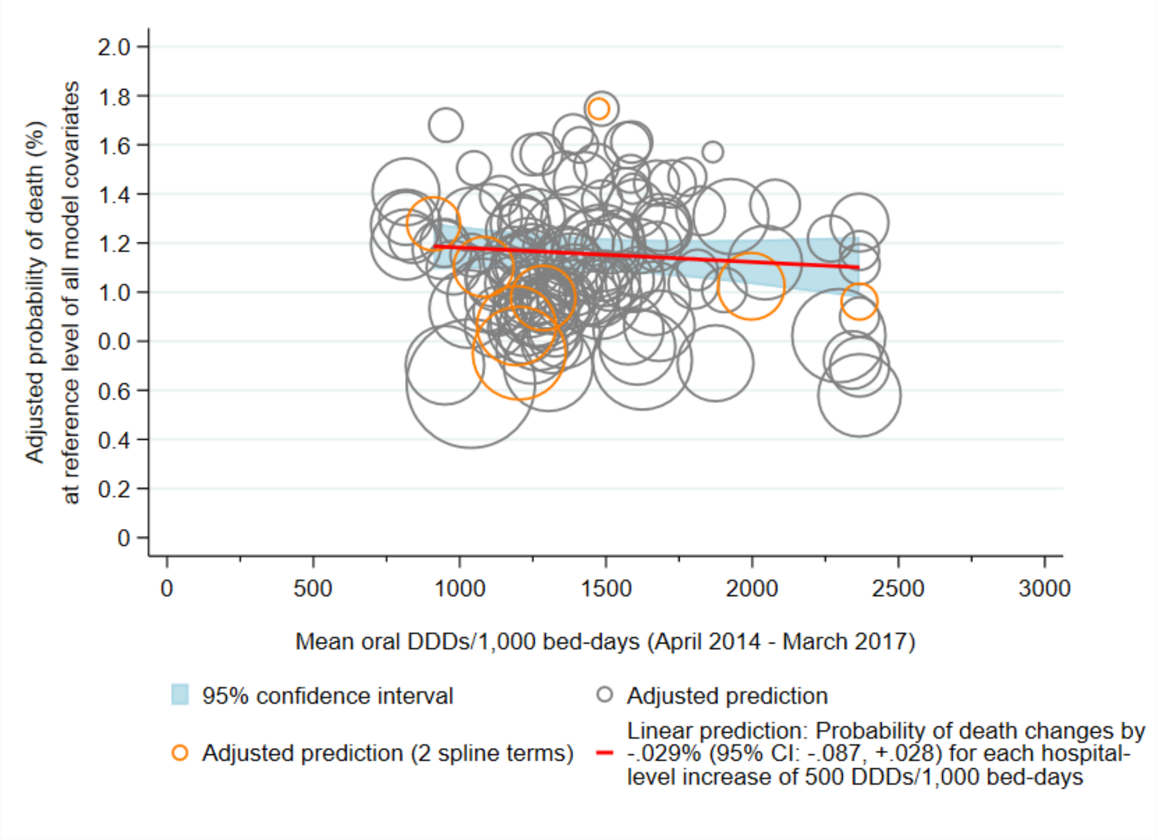

4.4b) Oral DDDs/1,000 admissions*

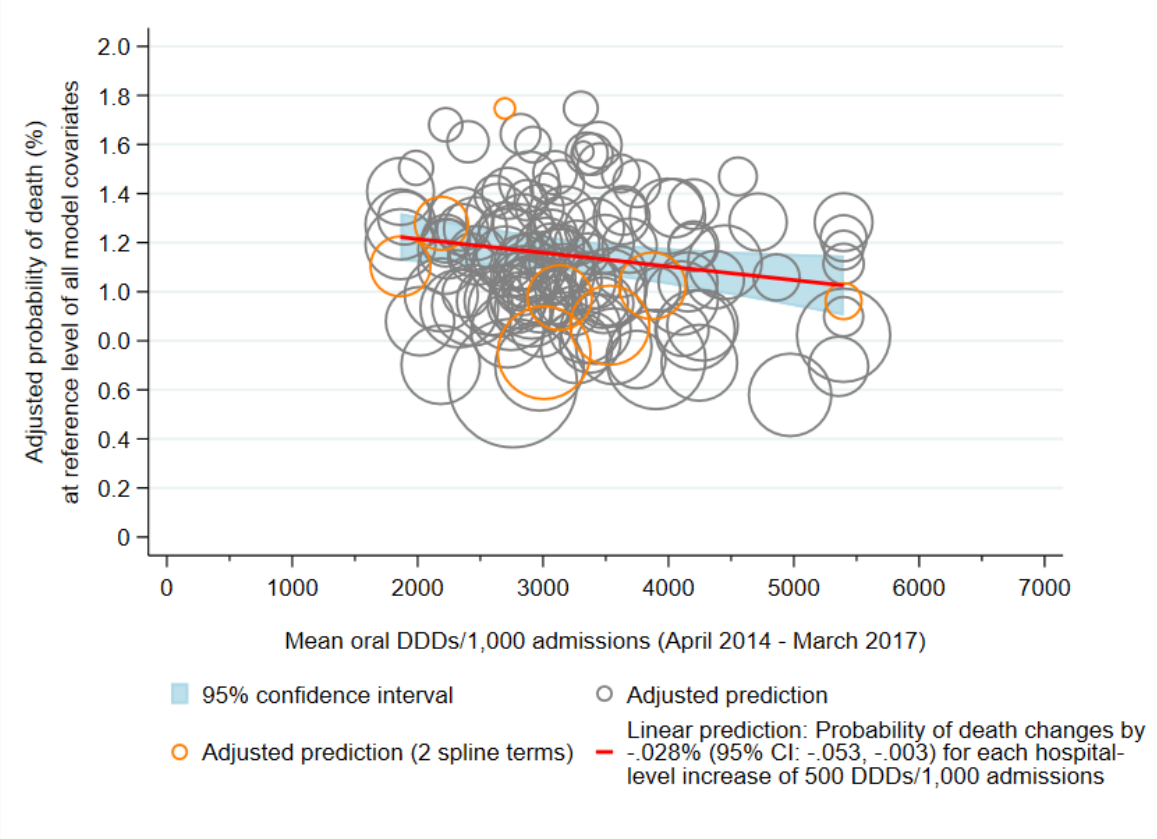

4.5a) Parenteral DDDs/1,000 bed-days*

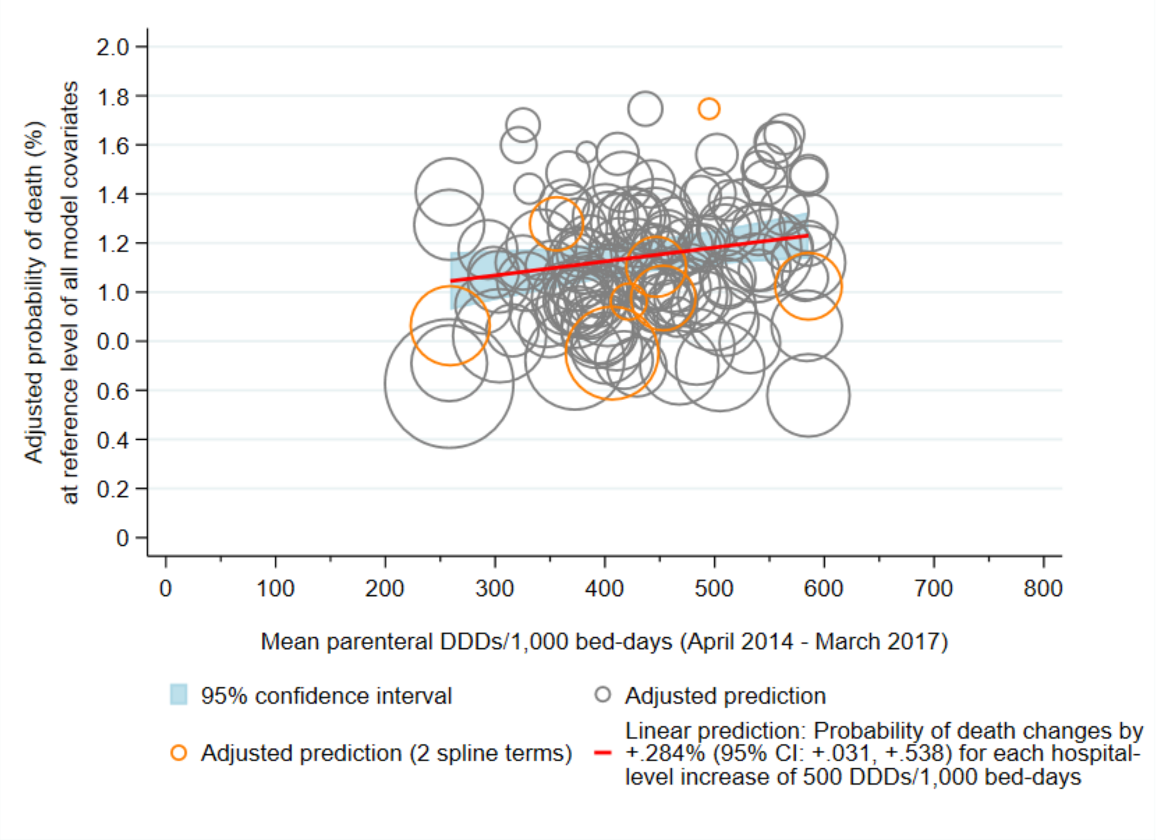

4.5b) Parenteral DDDs/1,000 admissions*

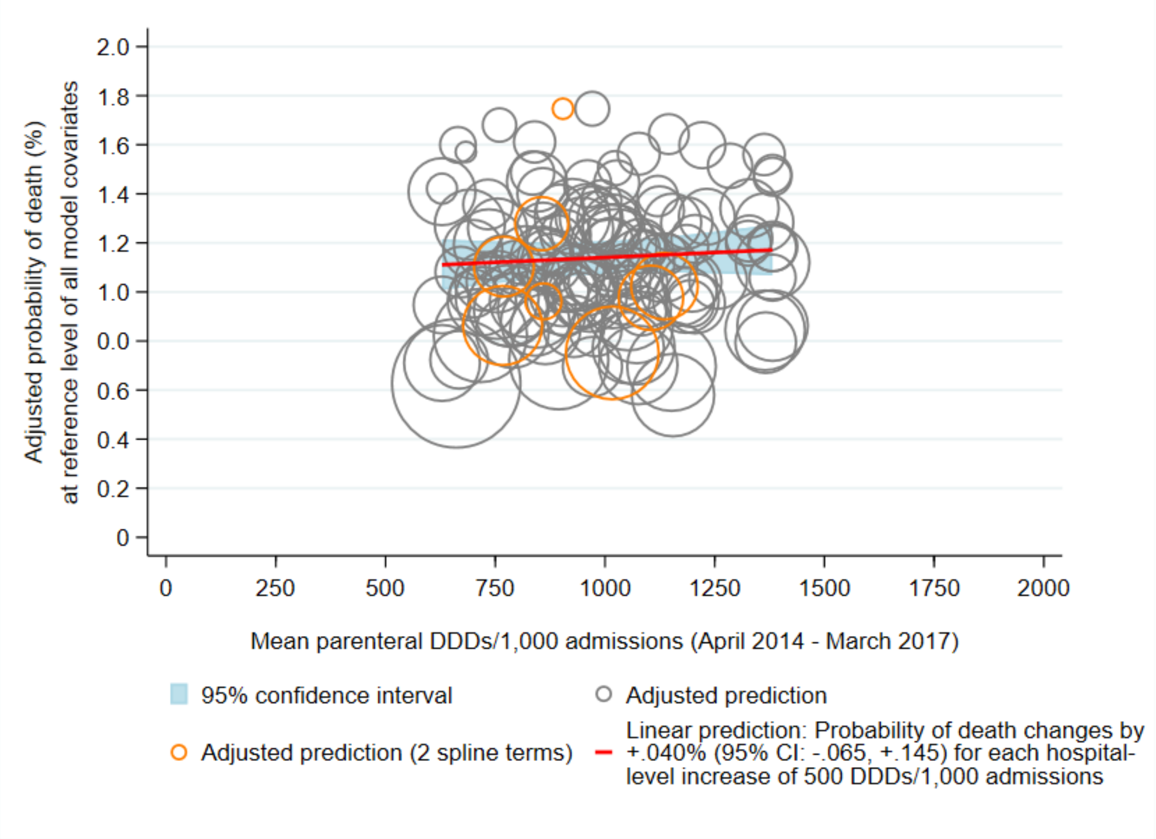

4.6a) Narrow-spectrum DDDs/1,000 bed-days*

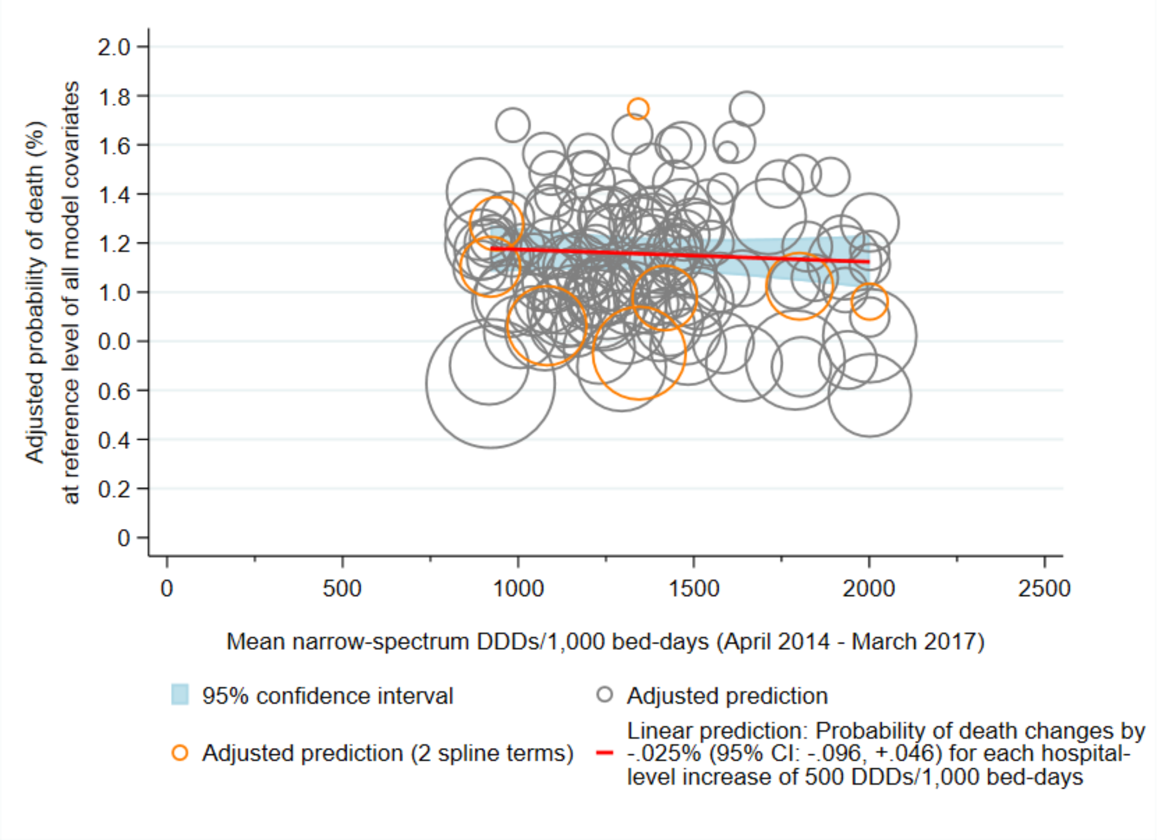

4.6b) Narrow-spectrum DDDs/1,000 admissions*

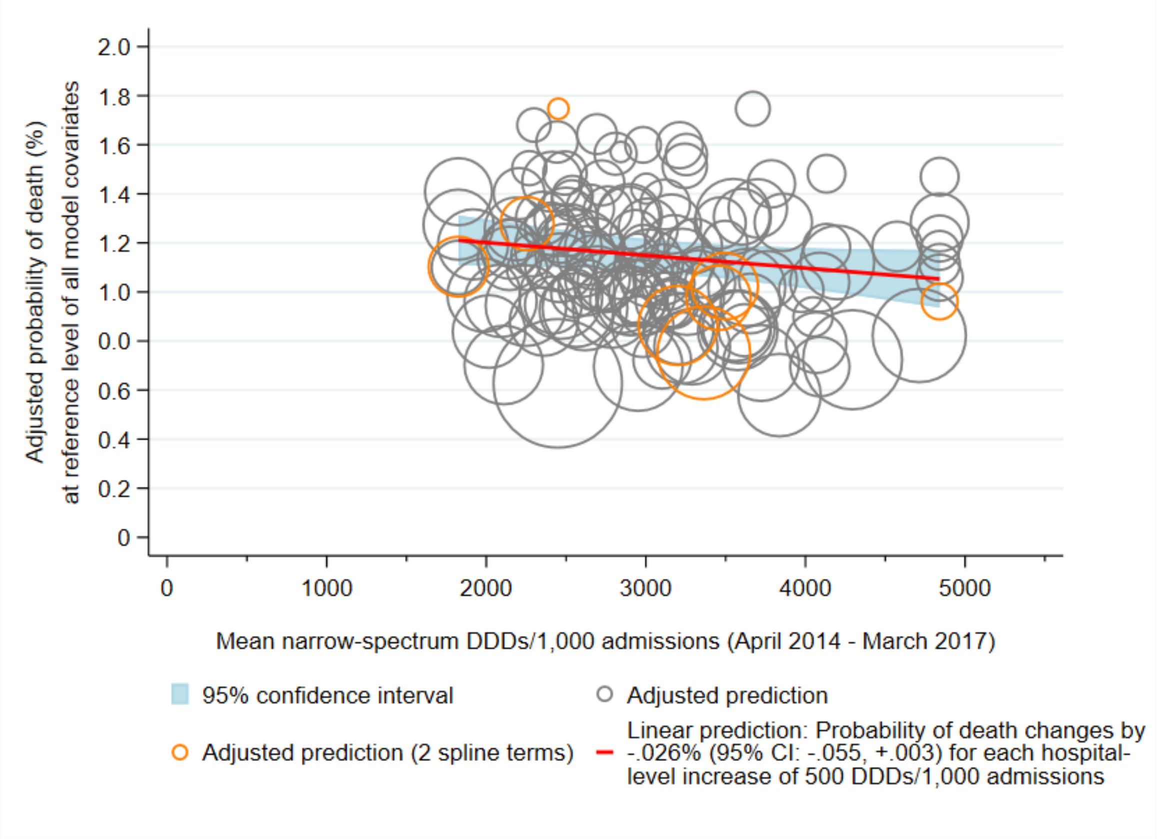

4.7a) Broad-spectrum DDDs/1,000 bed-days*

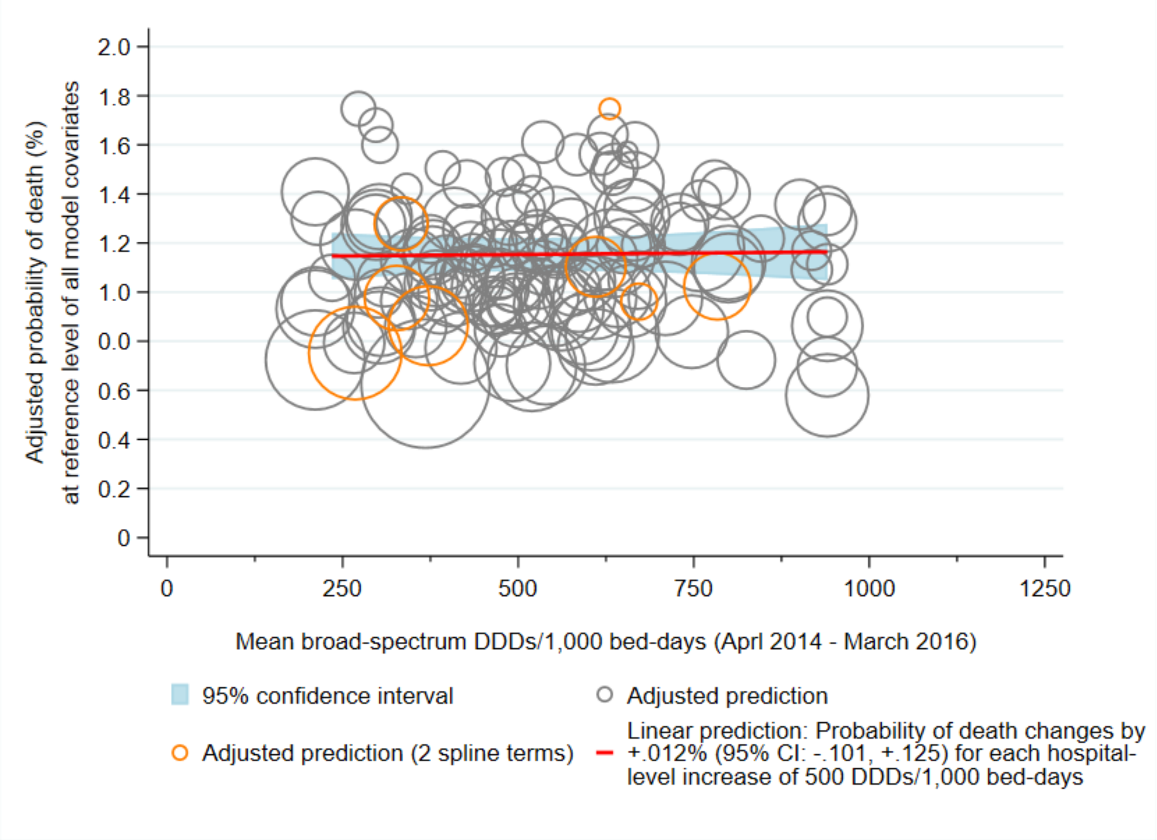

4.7b) Broad-spectrum DDDs/1,000 admissions*

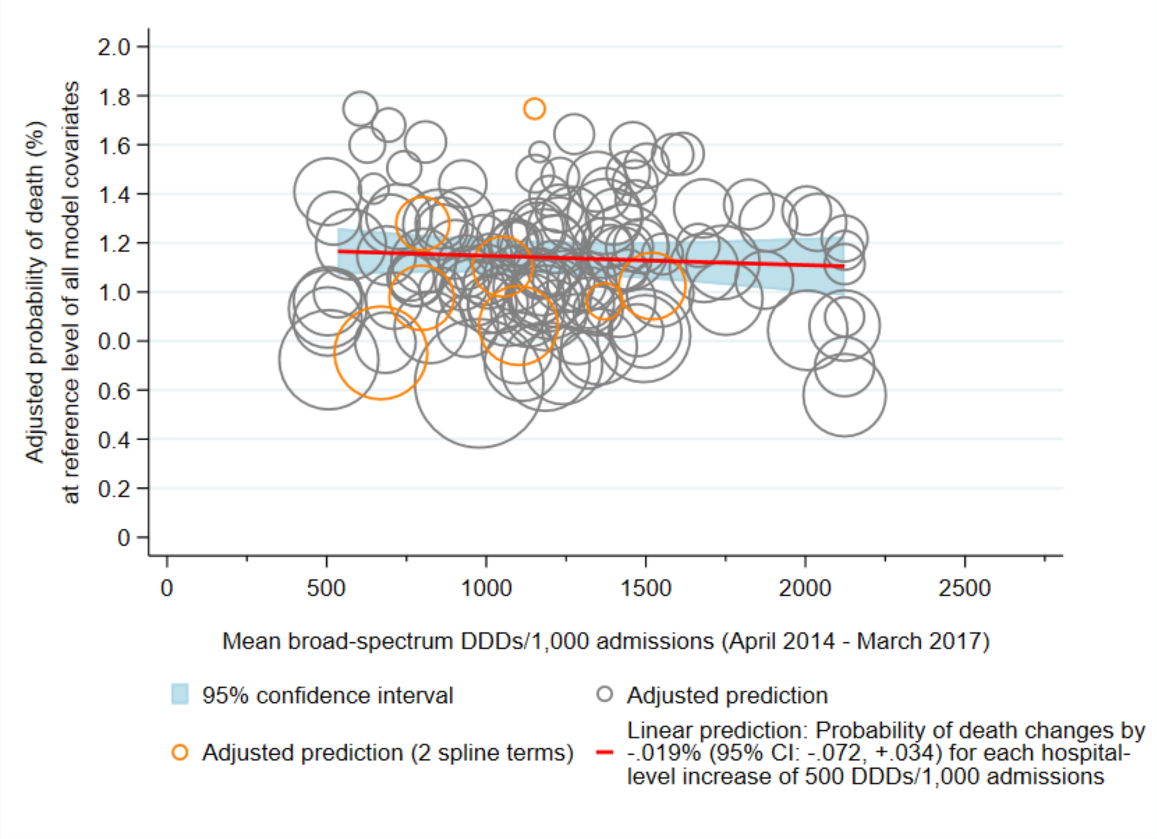

4.8a) Access DDDs/1,000 bed-days*

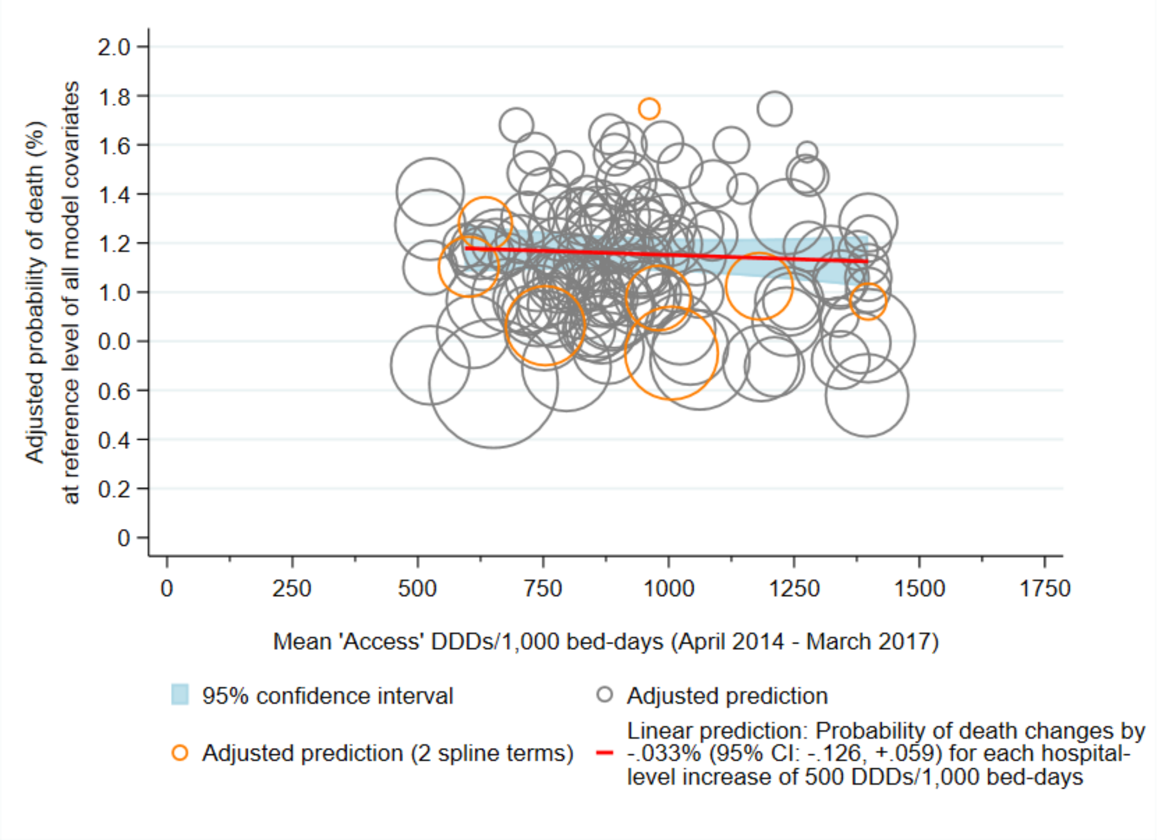

4.8b) Access DDDs/1,000 admissions*

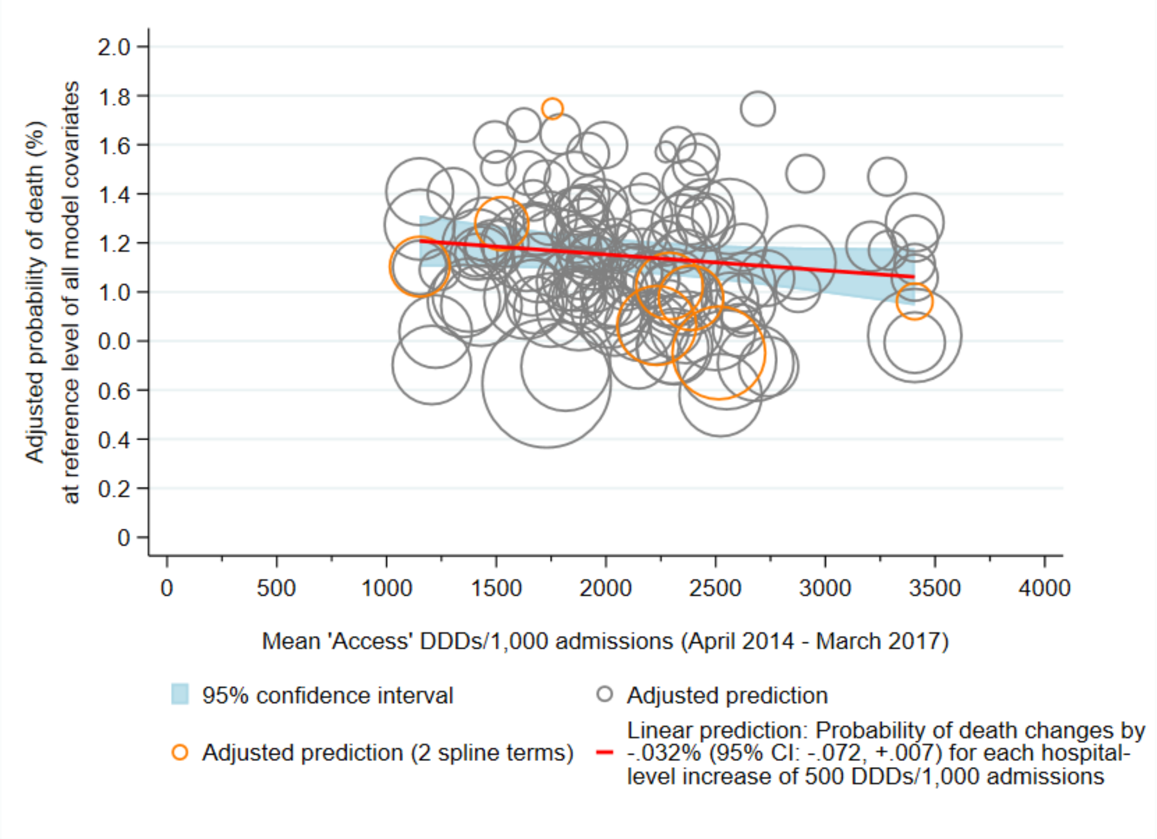

4.9a) Watch DDDs/1,000 bed-days*

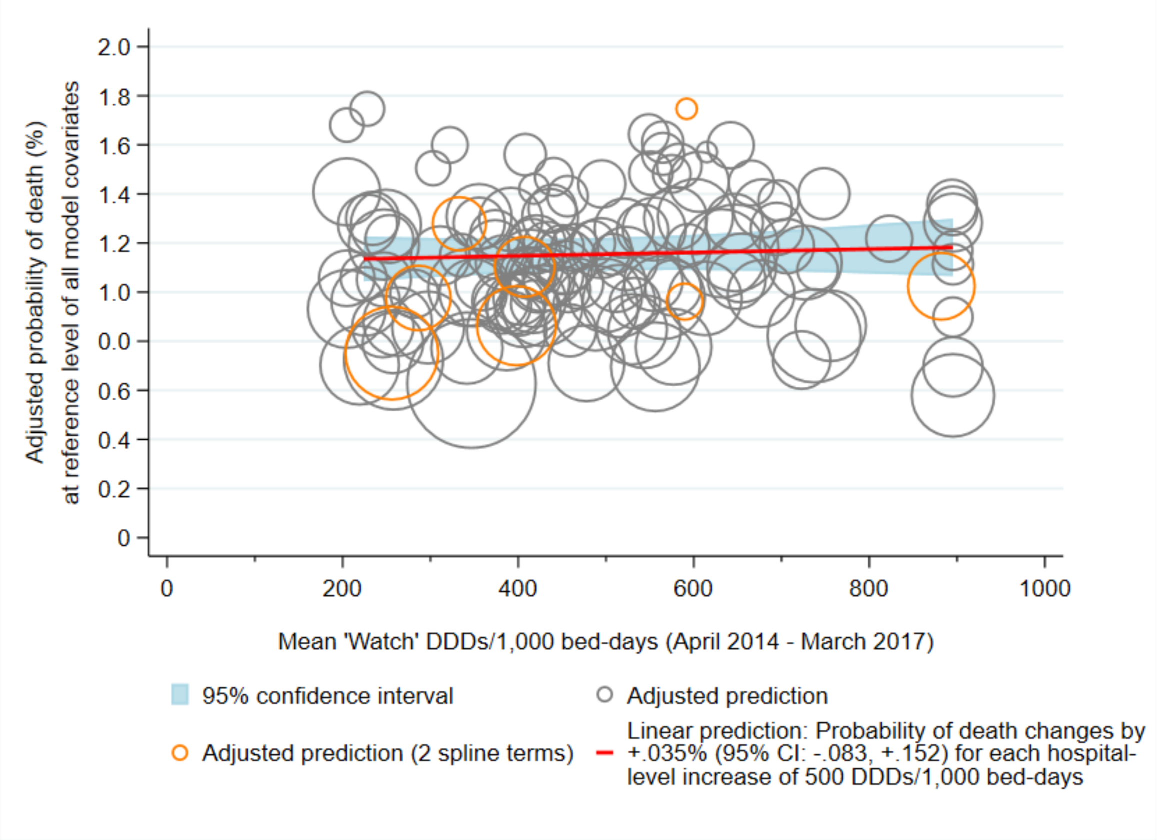

4.9b) Watch DDDs/1,000 admissions*

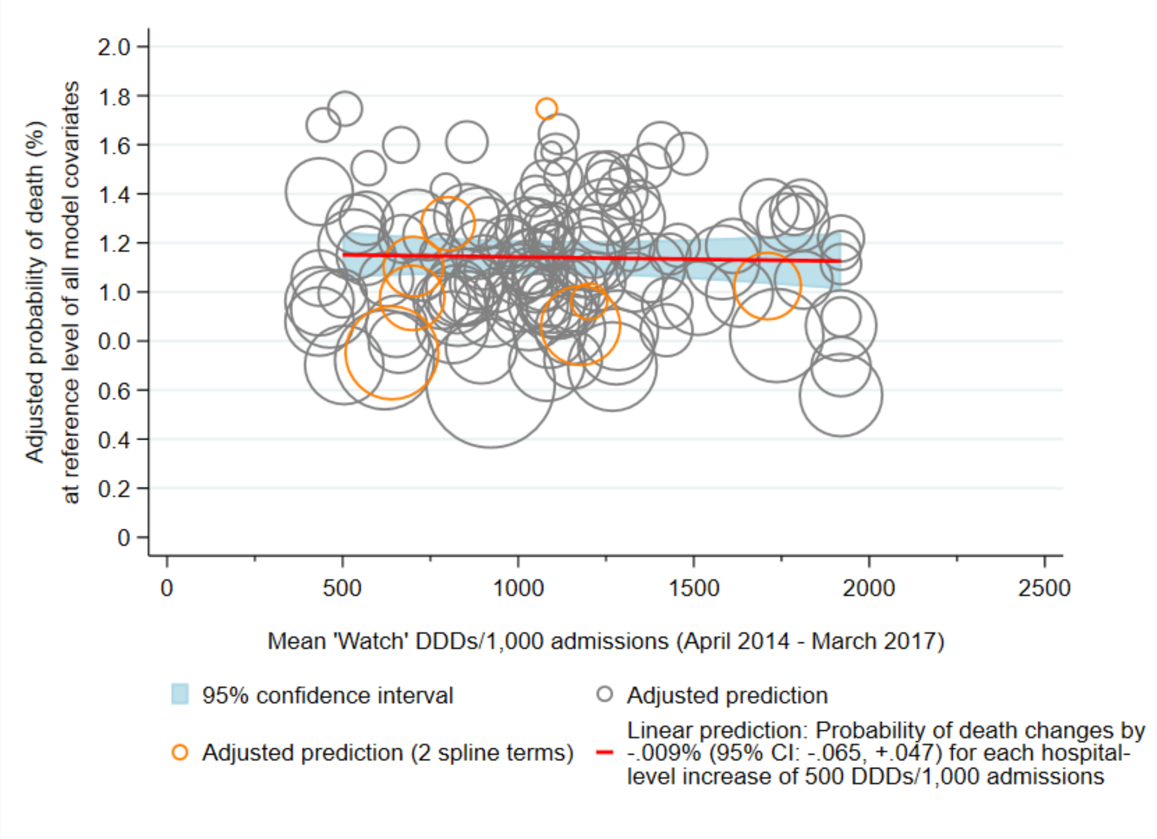

4.10a) Access or Watch (depending on indication) DDDs/1,000 bed-days*

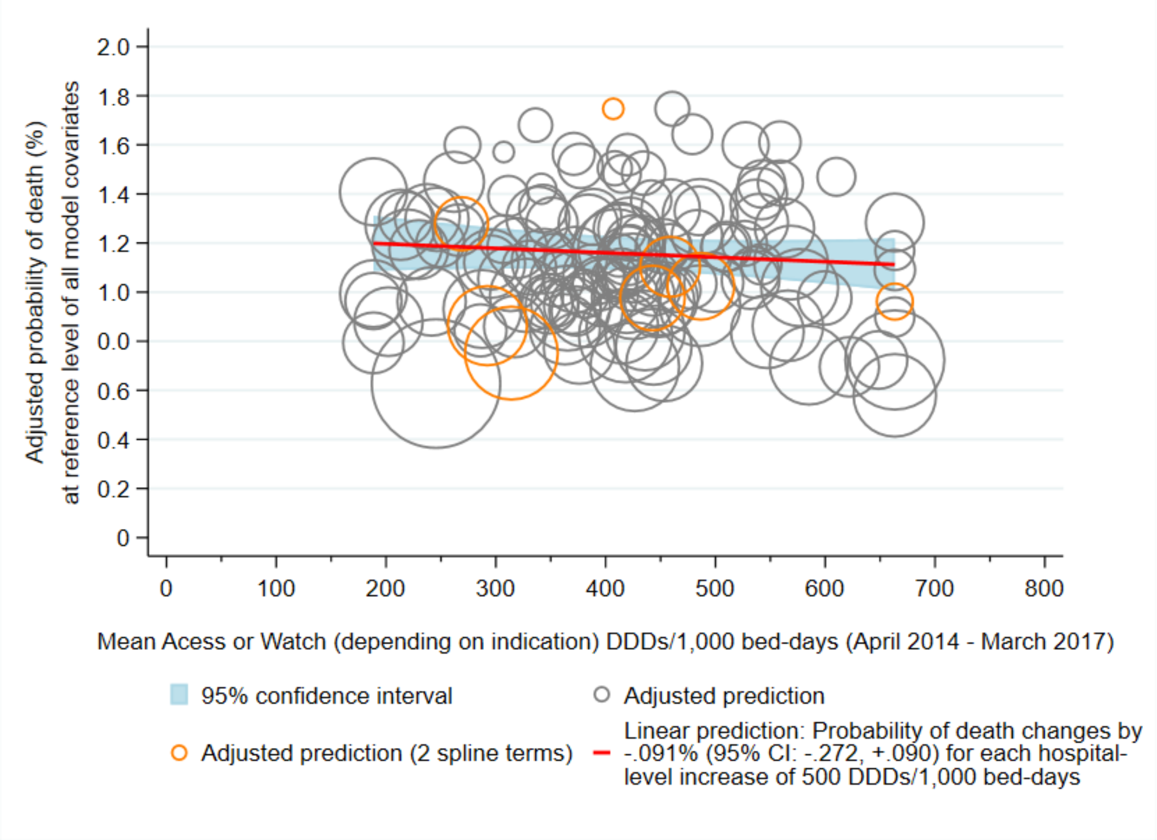

4.10b) Access or Watch (depending on indication) DDDs/1,000 admissions*

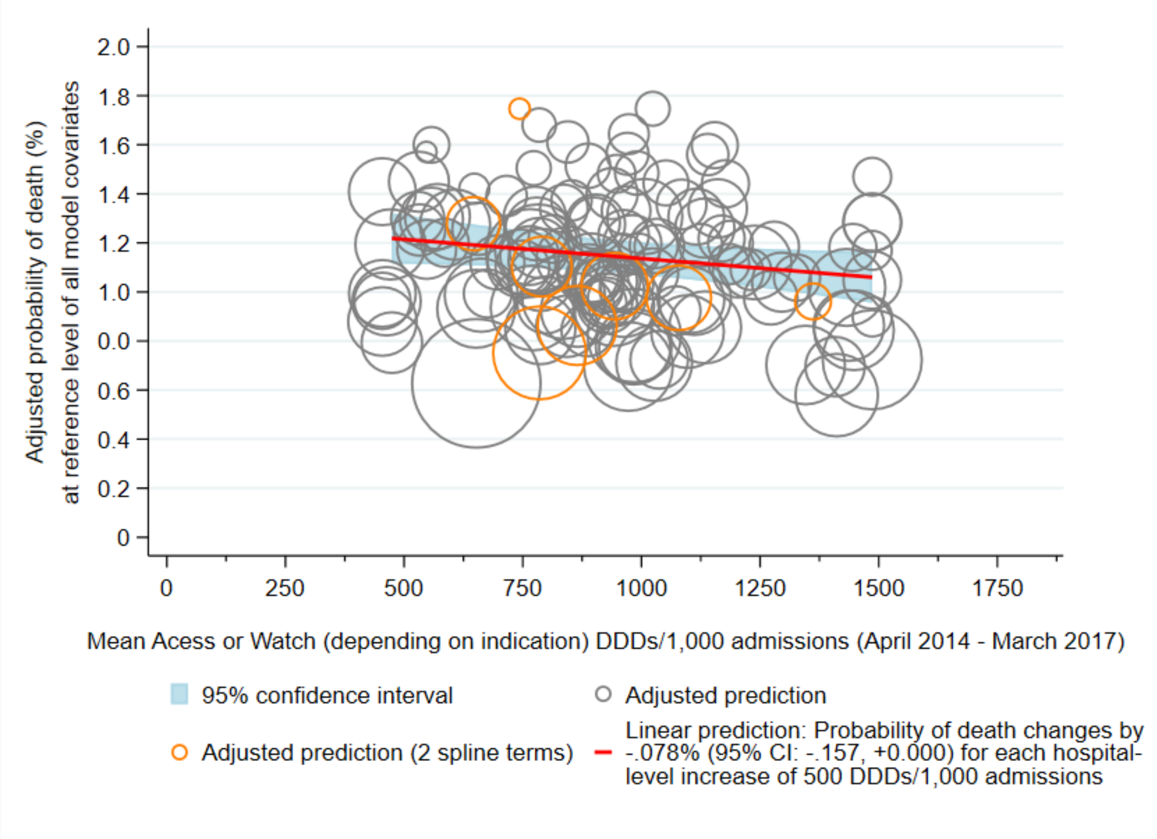

4.11a) Reserve DDDs/1,000 bed-days*

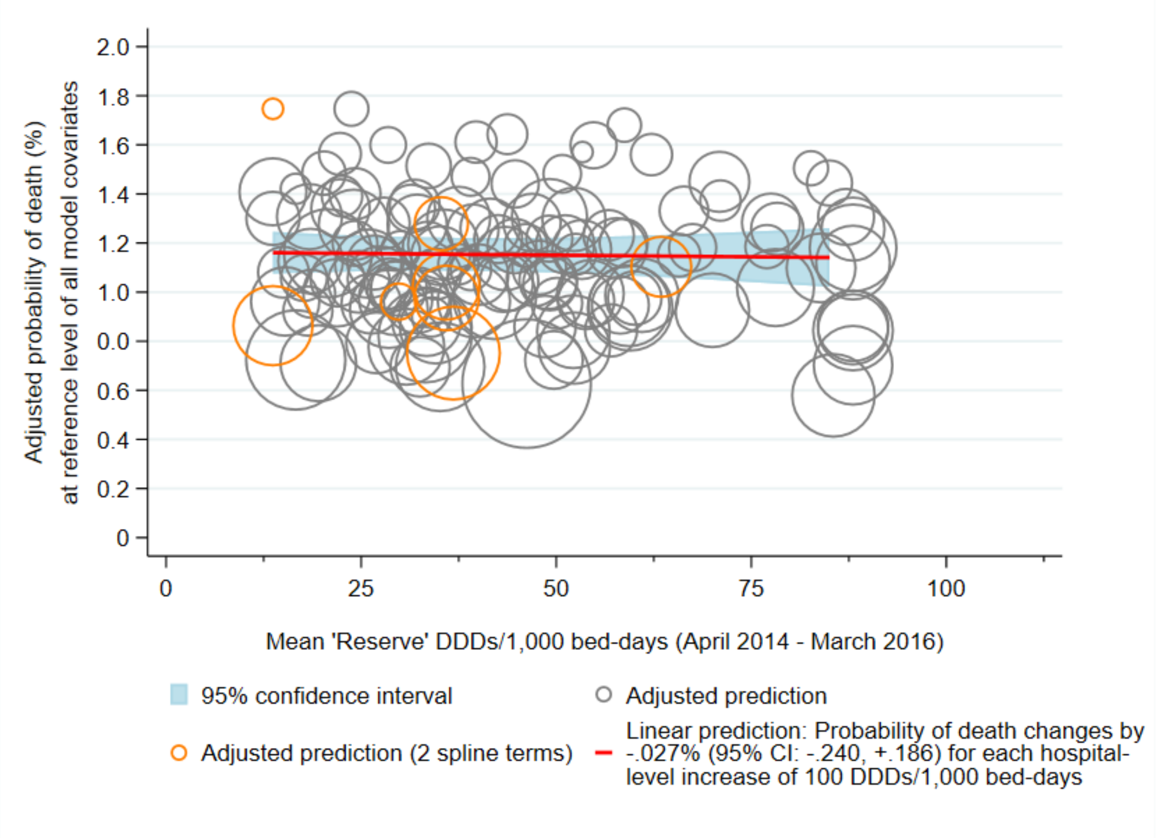

4.11b) Reserve DDDs/1,000 admissions*

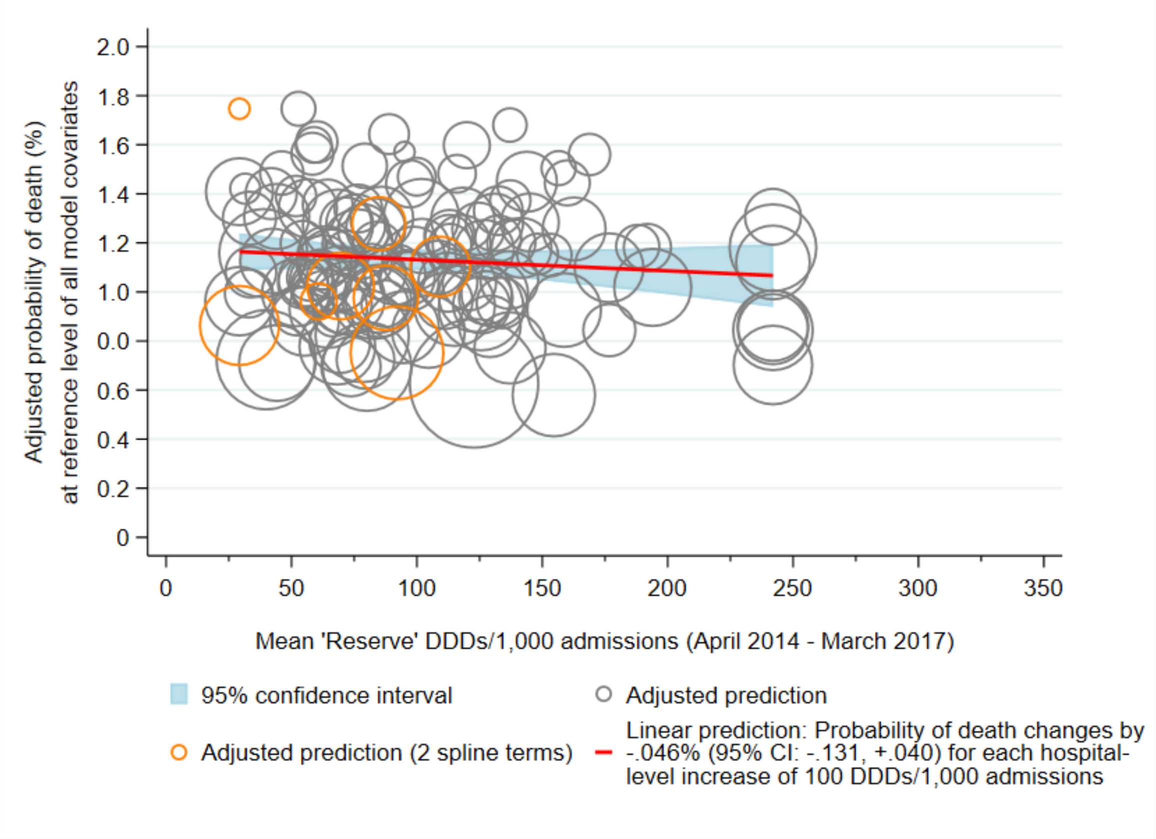

4.12a) Piperacillin/tazobactam and meropenem DDDs/1,000 bed-days*

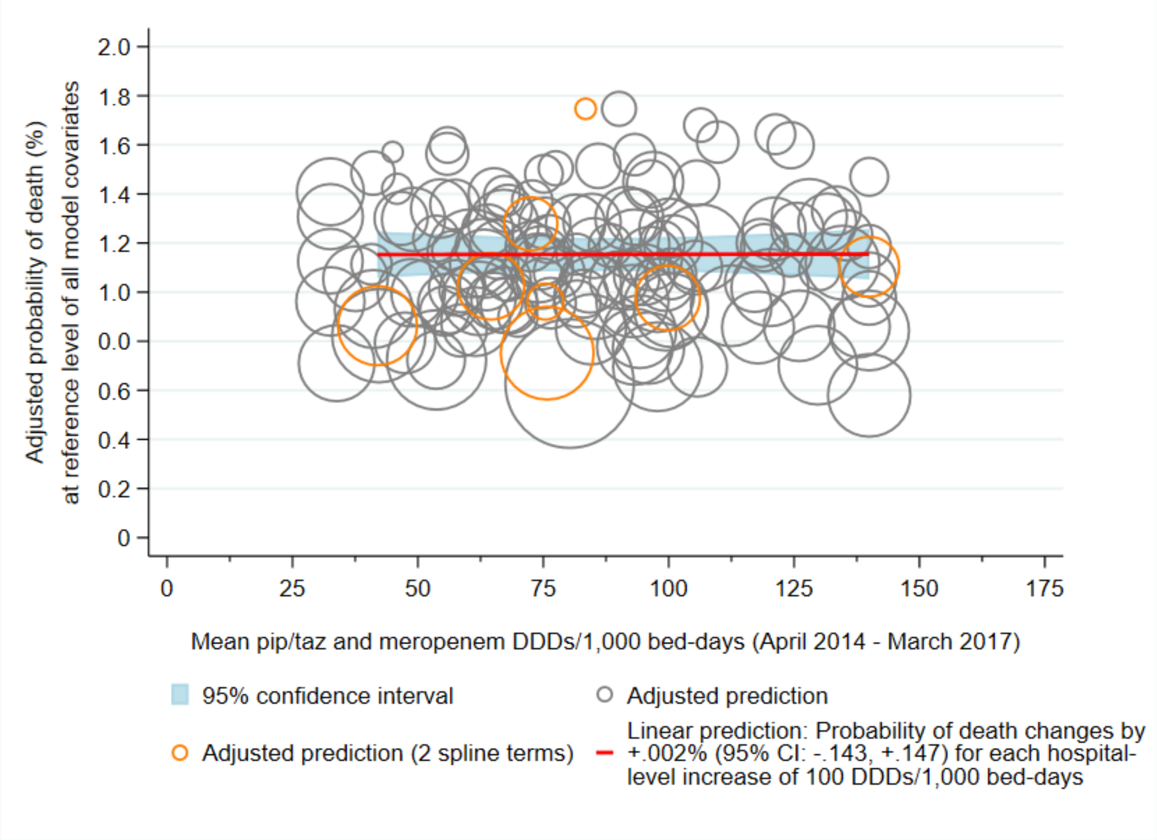

4.12b) Piperacillin/tazobactam and meropenem DDDs/1,000 admissions*

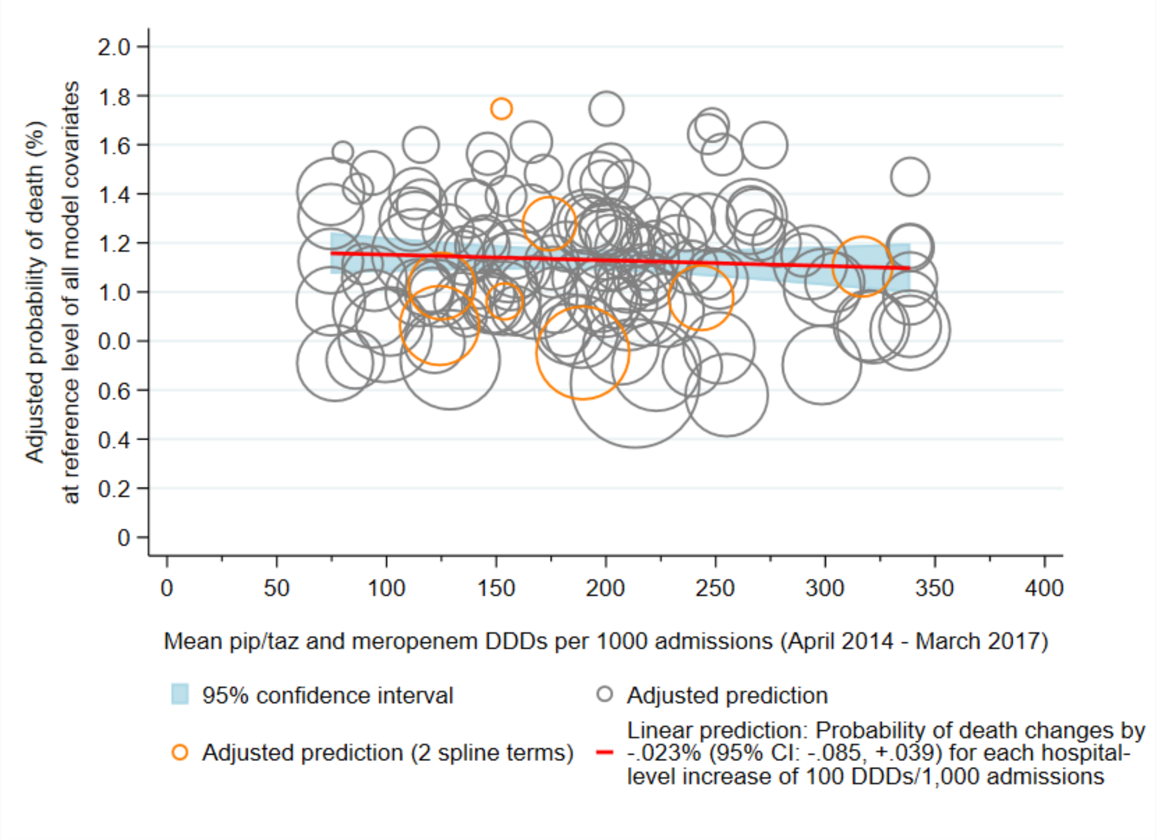

* Marginal effects were derived from 135 separate (hospital-specific) models, represented here as circles sized according to the precision of the estimate (inverse of the within-hospital variance). Most (127/135) hospital-specific multivariate models (grey circles) included four spline terms (for age, Charlson score, IMD score, and overnight admissions in the past year), however models for eight hospital Trusts (orange

circles) would only converge with two spline terms (age and overnight admissions in the past year). Antibiotic use was truncated below the 2.5 percentile and above the 95th percentile. Probability estimates were truncated above the 99th percentile.

**REFERENCES**

1. Herbert A, Wijlaars L, Zylbersztejn A, Cromwell D, Hardelid P. Data Resource Profile: Hospital Episode Statistics Admitted Patient Care (HES APC). *Int J Epidemiol*. 2017;46(4):1093-1093i. Available from: http://www.ncbi.nlm.nih.gov/pubmed/28338941

2. Health and Social Care Information Centre. Methodology to create provider and CIP spells from HES APC data. 2014. Available from: https://digital.nhs.uk/binaries/content/documents/corporate-website/publication-system/ci-hub/compendium-indicators/compendium-indicators/publicationsystem:cilandingasset%5B3%5D/publicationsystem:Attachments%5B12%5D/publicationsystem:attachmentResource

3. Public Health England. MRSA bacteraemia: annual data. Annual counts and rates of meticillin resistant Staphylococcus aureus (MRSA) bacteraemia by acute trust and clinical commissioning group (CCG). 2018. Available from: https://www.gov.uk/government/statistics/mrsa-bacteraemia-annual-data

4. NHS England. KH03 Guidance: Data Definitions. 2010. Available from: https://www.england.nhs.uk/statistics/statistical-work-areas/bed-availability-and-occupancy/

5. NHS Digital. Summary Hospital-level Mortality Indicator (SHMI) Indicator specification. Version 1.32. 2020. Available from: http://digital.nhs.uk/SHMI

6. Walker AS, Mason A, Quan TP, et al. Mortality risks associated with emergency admissions during weekends and public holidays: an analysis of electronic health records. *Lancet*. 2017 Jul 1;390(10089):62–72. Available from: http://linkinghub.elsevier.com/retrieve/pii/S0140673617307821

7. NHS Digital. HES Data Dictionary Admitted Patient Care. 2018. Available from: https://digital.nhs.uk/data-and-information/data-tools-and-services/data-services/hospital-episode-statistics/hospital-episode-statistics-data-dictionary

8. Harrell F. Regression modeling strategies: with applications to linear models, logistic and ordinal regression, and survival analysis. Springer; 2015.

9. Freemantle N, Richardson M, Wood J, et al. Weekend hospitalization and additional risk of death: an analysis of inpatient data. *J R Soc Med*. 2012 Feb;105(2):74–84. Available from: http://www.ncbi.nlm.nih.gov/pubmed/22307037

10. Harbord R, Higgins J. Meta-regression in Stata. *The Stata Journal*. 2008;8(4):493–519.

11. Greenland S, O’Rourke K. Meta-Analysis. In: Rothman K, Greenland S, Lash T, editors. Modern Epidemiology. 3rd ed. Philadelphia, USA: Lippincott Williams & Wilkins; 2008. p. 652–82.

12. Thompson S, Higgins J. How should meta-regression analyses be undertaken and interpreted? *Stat Med*. 2002;21(11):1559–73.

13. Berkey CS, Hoaglin DC, Mosteller F, Colditz GA. A random-effects regression model for meta-analysis. *Stat Med*. 1995 Feb 28;14(4):395–411. Available from: http://www.ncbi.nlm.nih.gov/pubmed/7746979

14. Jackson D, White IR, Thompson SG. Extending DerSimonian and Laird’s methodology to perform multivariate random effects meta-analyses. *Stat Med*. 2009 Apr 30;29(12):1282–97. Available from: http://www.ncbi.nlm.nih.gov/pubmed/19408255

15. Norton EC, Dowd BE, Maciejewski ML. Marginal Effects—Quantifying the Effect of Changes in Risk Factors in Logistic Regression Models. *JAMA*. 2019 Apr 2;321(13):1304. Available from: http://www.ncbi.nlm.nih.gov/pubmed/30848814

16. NHS Digital. Provider spells methodology: provider mapping files October 2019. 2019. Available from: https://digital.nhs.uk/data-and-information/publications/ci-hub/summary-hospital-level-mortality-indicator-shmi

17. NHS Digital. Hospital admitted patient care activity: provider level analysis. 2017. Available from: https://digital.nhs.uk/data-and-information/publications/statistical/hospital-admitted-patient-care-activity
